# A Systematic Performance Evaluation of Three Large Language Models in Answering Questions on moderate Hyperthermia

**DOI:** 10.64898/2026.03.25.26349254

**Authors:** Fabio Dennstädt, Nikola Cihoric, Nicolas Bachmann, Irina Filchenko, Luc M. Berclaz, Hans Crezee, Sergio Curto, Pirus Ghadjar, Boris Hübenthal, Mark D. Hurwitz, H. Petra Kok, Lars H. Lindner, Dietmar Marder, Jason Molitoris, Markus Notter, Sultan Rahman, Oliver Riesterer, Mateusz Spalek, Hana Trefna, Thomas Zilli, Dario Rodrigues, Markus Fürstner, Emanuel Stutz

**Author notes:** Corresponding author Emanuel Stutz, MD, Department of Radiation Oncology, Inselspital, Bern University Hospital, University of Bern, Bern, Switzerland.

## Abstract

**Background:** Large Language Models (LLMs) have demonstrated expert-level performance across many medical domains, suggesting potential utility in clinical practice. However, their reliability in the highly specialized domain of moderate hyperthermia (HT) remains unknown. We therefore evaluated the performance of three modern LLMs in answering HT-related questions.

**Methods:** We conducted an evaluation study by posing 40 open-ended questions—22 clinical and 18 physics-related—to three modern LLMs (DeepSeek-V3, Llama-3.3-70B-Instruct, and GPT-4o). Responses were blinded, randomized, and evaluated by 19 international experts with either a clinical or physics background for quality (5-point Likert scale: 1=very bad, 2=bad, 3=acceptable, 4=good to 5=very good) and for potential harmfulness in clinical decision-making.

**Results:** A total of 1144 quality evaluation responses were collected. Overall reported mean quality scores were similar across models, with DeepSeek scoring 3.26, Llama 3.18, and GPT-4o 3.07, corresponding to an “acceptable” rating. Across expert evaluations, responses were considered potentially harmful in 17.8% of cases for DeepSeek, 19.3% for Llama, and 15.3% for GPT-4o. Notably, despite “acceptable” mean scores, approximately 25% of responses were rated “bad” to “very bad,” and potentially harmful answers occurred in ∼15–19% of evaluations, indicating a non-trivial risk if used without domain expertise.

**Conclusion:** Our findings indicate that the performance of LLMs in HT in versions available at the time of investigation is only partially satisfactory. The proportion of poor-quality responses is too high and may lead non-domain experts to misinterpret the available clinical evidence and draw inappropriate clinical conclusions.

## INTRODUCTION

### Large Language Models

Large Language Models (LLMs), a form of generative Artificial Intelligence (AI), have remarkable capabilities in answering questions across various medical domains[1]. For example, the medically fine-tuned LLM Med-PaLM 2 correctly answered 86.5% of questions in the style of the United States Medical Licensing Exam [2]. With models now reportedly performing “at the level of an expert doctor” on such benchmarks, generative AI is expected to transform the clinical environment[3], having already demonstrated success in applications like medical writing, education, and diagnosis[4]. Other research has also noted impressive results from various LLMs like Llama 1, PaLM 2, Claude-v1, GPT-3.5, and GPT-4 in answering questions across different oncological fields[5]. These findings support the view that LLMs can serve as powerful assistant systems to augment clinical expertise and promote patient-centered care in oncology[6].

Given that LLMs can integrate extensive domain-specific knowledge, their use as assistant systems for answering clinical questions is a frequent topic of discussion[7].

While LLMs are successful in broad medical fields, the performance of LLMs in highly specialized subspecialties remains unexplored. Despite their increasing clinical visibility, these systems have generally not undergone medicine-specific quality assurance processes, nor have they received formal regulatory approval. The evaluation of LLMs in clinical practice is therefore not only important but also timely and necessary.

### Hyperthermia in oncology

The overarching term “thermal medicine” refers to the application of heat in oncology and comprises two main approaches: thermoablation and moderate hyperthermia (HT). Thermoablation is used as a standalone treatment[8], whereas moderate HT refers to the controlled therapeutic heating of tumors, commonly using electromagnetic waves, to achieve temperatures of 40–43 °C for approximately one hour. It is an adjuvant therapy typically administered shortly before or after radiotherapy (RT), or concurrently with chemotherapy (ChT), where it acts as a radio- and chemosensitizer[9–11]. The greatest benefit of HT has been demonstrated with shorter intervals between RT and HT and when sufficiently high intratumoral temperatures are achieved[12–15]. This places specific requirements on the heating capabilities of HT devices and on the quality of treatment delivery. Therefore, evidence-based quality assurance guidelines have been issued by the European Society of Hyperthermic Oncology (ESHO), Society for Thermal Medicine (STM), and the German Atzelsberg Circle[16–19]. However, the application of heat is also common in alternative medicine among cancer patients, where a substantial amount of non-scientific or not evidence-based information is available on the internet.

The above-mentioned background is important for understanding the challenges faced by LLMs in HT. This a niche discipline that is gaining increasing attention in the field of evidence-based oncology. Owing to their accessibility and ease of use, LLMs may be consulted for decision support by clinicians seeking an overview of the HT field, those newly practicing in this area, and patients exploring treatment options in situations of uncertainty[20]. However, available HT literature and data remain relatively limited compared with more established cancer therapies and may be interspersed with non-scientific information, which may negatively affect LLM training and increase the risk of factual errors, “hallucinations” and inaccurate statements[21, 22]. To date, no formal evaluations of LLM performance have been reported in the field of moderate HT, representing a critical knowledge gap that this study aimed to systematically address.

## METHODS

### Study design

We assessed the quality, correctness and potential harmfulness of responses from three state-of-the-art LLMs, as evaluated by international domain experts, in collaboration with the International Society for Radiation Oncology Informatics (ISROI) and ESHO. Based on methodologies used in previous clinical evaluation studies of LLMs [23, 24], we posed open-ended questions from the field of HT to the LLMs and had the responses independently evaluated by domain experts. Before model querying, the study coordinators predefined the question set to cover major domains of moderate HT, including indications, treatment integration with RT/ChT, technical delivery, QA, devices, and areas of current controversy. A schematic overview of the study design is presented in **Figure 1**.

**Figure 1:**
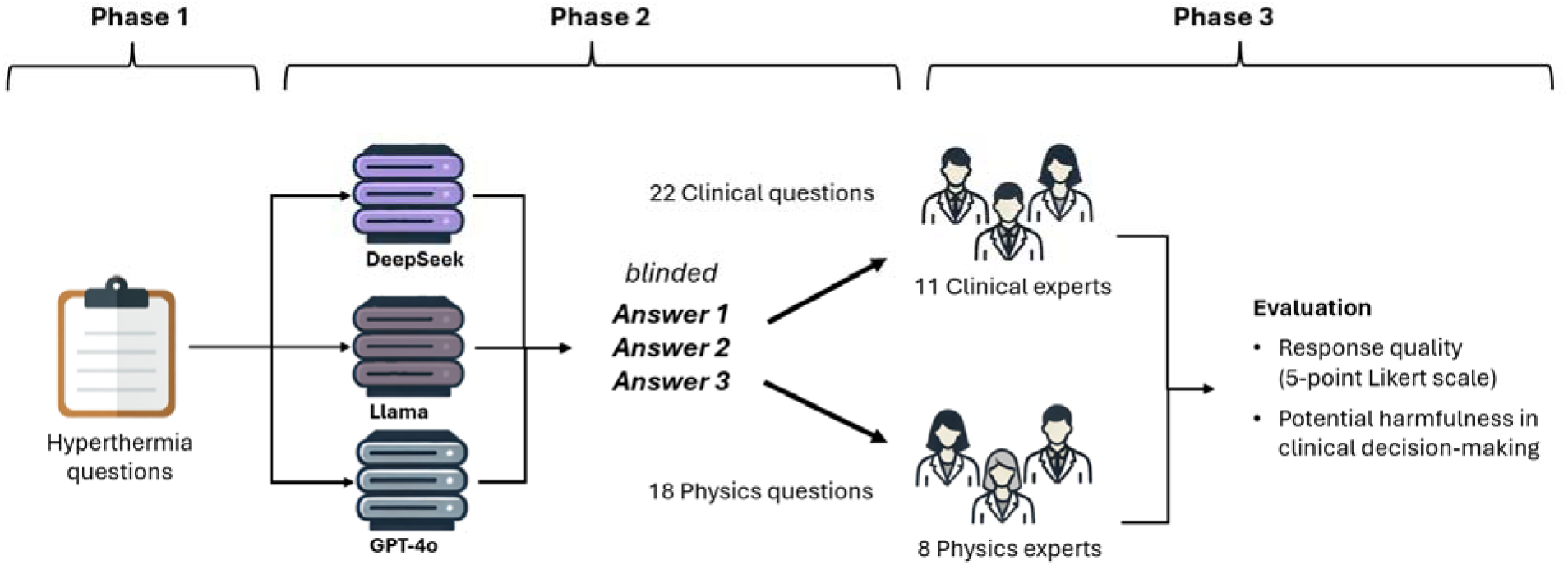
Schematic illustration of the study design. Relevant questions from the field of HT were developed by the study coordinators (authors: ES, FD, NC) in Phase 1 and answered by three LLMs in Phase 2. In Phase 3, these blinded question-answer pairs were evaluated by HT domain experts with clinical and physics backgrounds.

### Phase 1: Development of moderate hyperthermia-related study questions

In the first phase, the study coordinators (ES, NC, and FD) created a list of open-ended questions (**Supplementary Table S1 and S2**) about HT covering a wide range of topics and current challenges spanning both the clinical and physics aspects of the field. The first draft was reviewed by NB and MF. Each question was labeled as either “clinical” or “physics.” Open-ended questions were intentionally chosen to better mimic real-life medical practice, where complex problems rarely have a single, clearly correct answer, in contrast to typical multiple-choice exam questions. One group of questions was designed to elicit responses based on established knowledge (“ground truths”), whereas the other comprised questions reflecting common controversies in the field, intended to assess the reasoning and argumentative capabilities of the LLMs. A total of 40 HT-related questions were used, comprising 22 questions focusing on clinical aspects and 18 questions focusing on physics-related aspects.

### Phase 2: Response generation by three LLMs

To answer the questions, three modern LLMs were selected to represent different types of generative AI models available at the time of the study (April 2025). We focused on evaluating general, non-medical LLMs. In contrast to medical LLMs, general LLMs are widely accessible, cost-effective, and are used by healthcare professionals as well as by patients. The selected LLMs were:

- DeepSeek-V3 (DeepSeek)[25]: Developed by DeepSeek AI, this is a mixture-of-experts (MoE) model with 236 billion parameters. For any given token, it activates only 21 billion parameters, which makes it highly efficient. It is designed for natural language processing and code generation. DeepSeek-V3 is open-source and designed to be a cost-effective yet powerful option.
- Llama-3.3-70B-Instruct (Llama)[26]: This is a 70-billion parameter, open-weight model developed by Meta, optimized for following complex instructions in multiple languages). It is part of the Llama 3 series and is designed for large-scale AI applications. A key feature is its ability to follow complex instructions accurately.
- GPT-4o[27]: Developed by OpenAI, GPT-4o (“o” for omni) is a multimodal model that can process and generate text, audio, and image inputs and outputs. Compared with earlier versions, GPT-4o integrates these modalities within a single neural network, enabling more natural and real-time interactions.

A Python script was created to query the LLMs and collect their responses to the questions. The DeepSeek model and the Llama model were accessed through the cloud service DeepInfra[28], whereas GPT-4o, a proprietary model developed by OpenAI, was accessed through OpenAI API. Further details on the technical execution as well as the Python script are provided on GitHub[29].

All the study questions were answered by all three LLMs. No restrictions on text length were imposed. For the evaluation, the responses were collected exactly as generated by the models without modification.

### Phase 3: Expert evaluation of LLM responses

International HT domain experts with clinical or physics expertise, as well as clinical users, were contacted for participation in this evaluation study. Following a pilot run that included HT users from five Swiss centers within the Swiss HT Network[30], the expert panel was expanded to include international experts who are active members of ESHO. Of the 22 invited experts, 19 consented to participate, while 3 declined due to time constraints. These participants represented 13 distinct clinical departments across six countries in Europe and the United States.

To prepare the evaluation materials, the three answers for each question were blinded to mask their LLM origin and their presentation order was randomized using a pseudo-random number generator[31]. The 11 experts with a clinical background evaluated responses to the clinical questions, whereas the eight experts with a physics background evaluated responses to the physics-related questions.

The quality of each individual response was rated using a 5-point Likert scale. Scores ranged from 1 – very bad, 2 – bad, 3 – acceptable, 4 – good, to 5 – very good. To standardize the assessment of open-ended questions that lack a single definitive answer, evaluators were instructed to ground their quality ratings on widely accepted medical knowledge. In addition, the experts indicated whether they considered a response potentially harmful if used in clinical decision-making (binary variable: yes/no). The open-source online platform SmartOncology[32] was used for collecting the evaluations of the domain experts.

### Data and statistical analysis

The statistical analyses were performed in R (version 4.3.1). All statistical tests were two-sided and conducted at a significance level of 5%. Unless stated otherwise, continuous variables (Likert scores) were presented as median and interquartile range, while binary variables (harmfulness ratings) were presented as counts and percentages. Missing values were not imputed.

First, interrater agreement (IRA) was assessed. For quality ratings, which were measured on an ordered Likert scale, we used the intraclass correlation coefficient (ICCk2) for average random raters, as it quantifies the consistency of ratings across multiple raters while accounting for both within- and between-rater variability [33]. The values of ICC were interpreted as poor (<0.5), moderate (0.5–0.75), good (0.75–0.9), and excellent (>0.90). We also quantified within-group interrater agreement using the r_wg_ statistic for individual questions, which ranges from 0 (no agreement) to 1.0 (perfect agreement) [34]. For harmfulness ratings, agreement was quantified using the observed agreement rate [35], defined as the proportion of raters agreeing on the most common response.

We compared model performance across three LLMs. Quality was expressed as the mean score per question per rater, whereas harmfulness was expressed as the percentage of raters who classified the response as potentially harmful. Comparisons between LLMs were performed using Wilcoxon signed-rank test, with false discovery rate (FDR) correction applied to account for multiple comparisons. Analyses were conducted for all questions combined and separately for clinical and physics questions.

The question-response sets with expert evaluations were additionally reviewed by the study coordinators. Specifically, the question–response pairs with the highest and lowest ratings were examined in detail. In addition, one representative example was selected to illustrate the challenges associated with evaluating free-text responses and the potential sources of disagreement among reviewers.

### Ethical considerations

The study did not involve any patient- or person-related healthcare data. Therefore, approval from an ethics committee was not required.

## RESULTS

### Hyperthermia questions and corresponding answers

The median length of the questions was 14.5 (12–17) words, with 16.0 (13.3–22.8) words for the clinical and 13.5 (11.25–14.75) words for the physics questions.

For clinical and physics-related questions, as well as overall, the median answer length was 287.0 (216.8–394.3), 364.5 (343.3–394.3), and 323 (237–376) words for DeepSeek; 362.0 (307.8–390.0), 418.0 (387.8–467.8), and 387.0 (343.8–440.5) words for Llama; and 197.5 (159.5–250.5), 336.5 (297.0–375.5), and 265.0 (190.5–321.8) words for GPT-4o, respectively.

All questions, the corresponding responses of the LLMs and the individual ratings of the domain experts are provided in **Supplementary Table S1 and S2**.

### Interrater Agreement

For the clinical questions, the IRA was moderate, with an ICC of 0.64 for quality ratings and an observed agreement rate of 45.5% (36.4–48.9%) for the assessment of harmfulness. For the physics-related questions, agreement was also moderate, with an ICC of 0.64 for quality ratings and an observed agreement rate of 62.5% (62.5–87.5%) for the assessment of harmfulness.

### Quality of the answers

The three LLMs generated 66 responses to the 22 clinical questions, which were evaluated by 11 clinical domain experts, yielding 726 potential ratings, of which 723 were available for analysis. For the 18 physics-related questions, 54 responses were assessed by 8 physics domain experts, resulting in 432 potential ratings, of which 421 were available. Overall, 1144 of 1158 possible evaluations were completed, with missing ratings attributable to saving errors or missed questions. The mean quality scores across all responses (clinical and physics combined) were 3.26 for DeepSeek, 3.18 for Llama and 3.07 for GPT-4o, with a median score of 3 for all models.

For the clinical questions, the mean quality scores were 3.40 (median 4) for DeepSeek, 3.41 (median 4) for Llama, and 3.24 (median 3) for GPT-4o. For the physics questions, the mean scores were 3.02 (median 3) for DeepSeek, 2.80 (median 3) for Llama, and 2.77 (median 3) for GPT-4o. An overview of the distributions is provided in **Figure 2**, and box-plot diagrams illustrating the performance of the three LLMs are shown in **Figure 3**.

**Figure 2:**
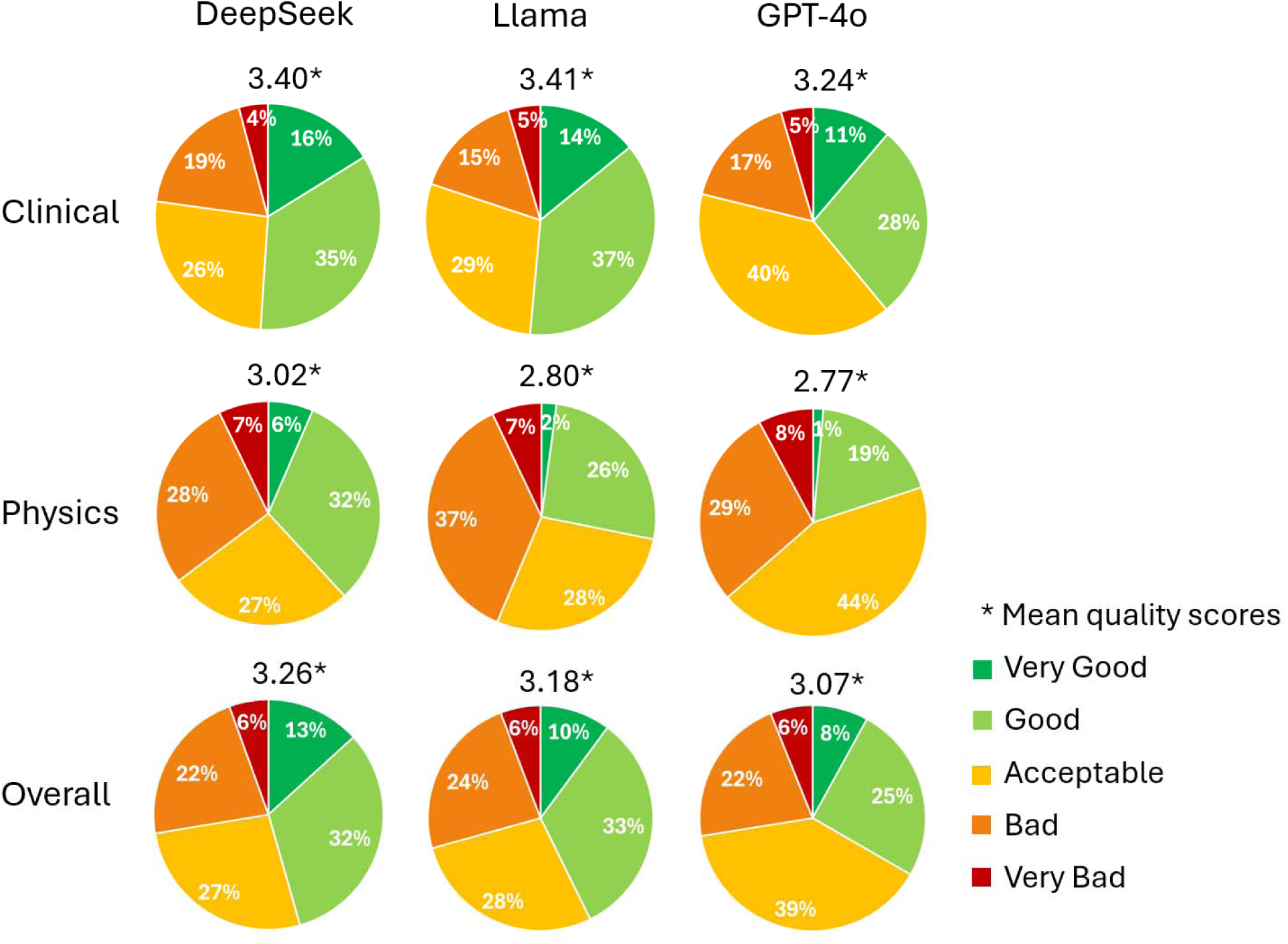
Mean quality scores with their distributions across the Likert Scale from 1 (Very Bad) to 5 (Very Good) for the answers given by the three LLMs: DeepSeek, Llama and GPT-4o.

**Figure 3:**
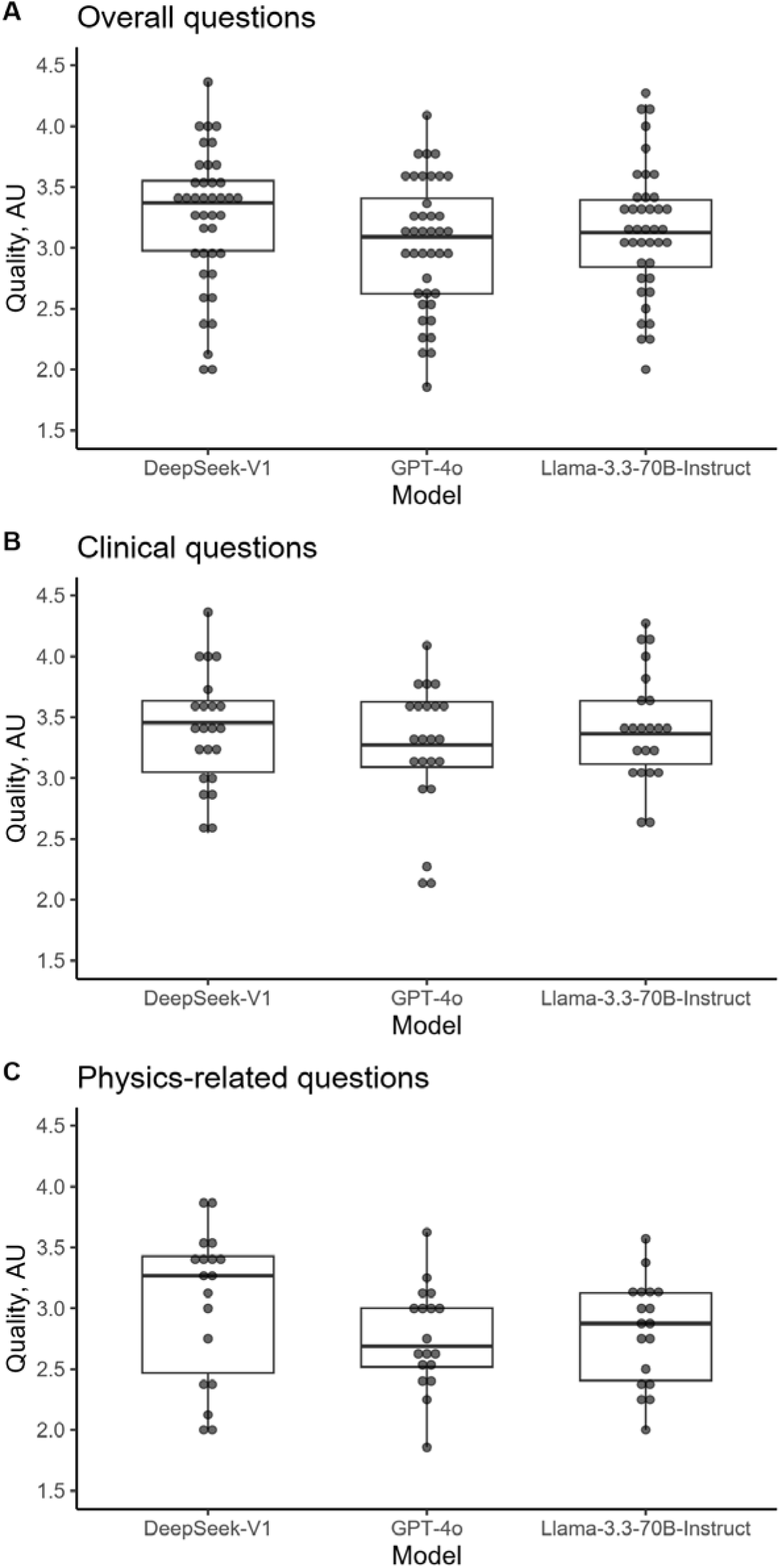
Box plots of quality ratings. (A) Overall questions. (B) Clinical questions. (C) Physics-related questions. All p > 0.05 (Wilcoxon signed-rank test with false discovery rate correction for multiple comparisons). Each point represents the mean of quality rating of a single question.

DeepSeek showed significantly higher answer quality than Llama for the physics-related questions (p = 0.046). However, this difference was not significant after adjustment for multiple comparisons. No other statistically significant differences in quality were observed between LLMs.

### Potential harmfulness of responses

Potential harmfulness was assessed in 1136 of 1158 ratings (722/726 for clinical questions and 414/432 for physics-related questions) with missing ratings attributable to saving errors or missed questions. With respect to the overall frequency of responses flagged as potentially harmful, DeepSeek responses were classified as potentially harmful in 67 of 377 evaluations (17.8%), compared with 73 of 379 evaluations (19.3%) for Llama, and 58 of 380 evaluations (15.3%) for GPT-4o. When analyzed at the question level, 14 of the 22 clinical responses (63.6%) generated by DeepSeek were rated as potentially harmful by at least one expert. Similar proportions were observed for Llama (14/22; 63.6%) and GPT-4o (13/22; 59.1%).

Performance regarding harmfulness was even poorer for physics-related question-responses pairs. At least one expert identified potential harmfulness in responses to 14 of 18 questions (77.8%) for DeepSeek, all 18 responses (100%) for Llama, and 15 of 18 responses (83.3%) for GPT-4o. However, there were no significant differences in harmfulness between LLMs. An overview of the proportion of experts indicating a response as potentially harmful is provided in the heatmap in **Figure 4** for all individual responses. Box plots illustrating the performance of the three LLMs are shown in **Figure 5**.

**Figure 4:**
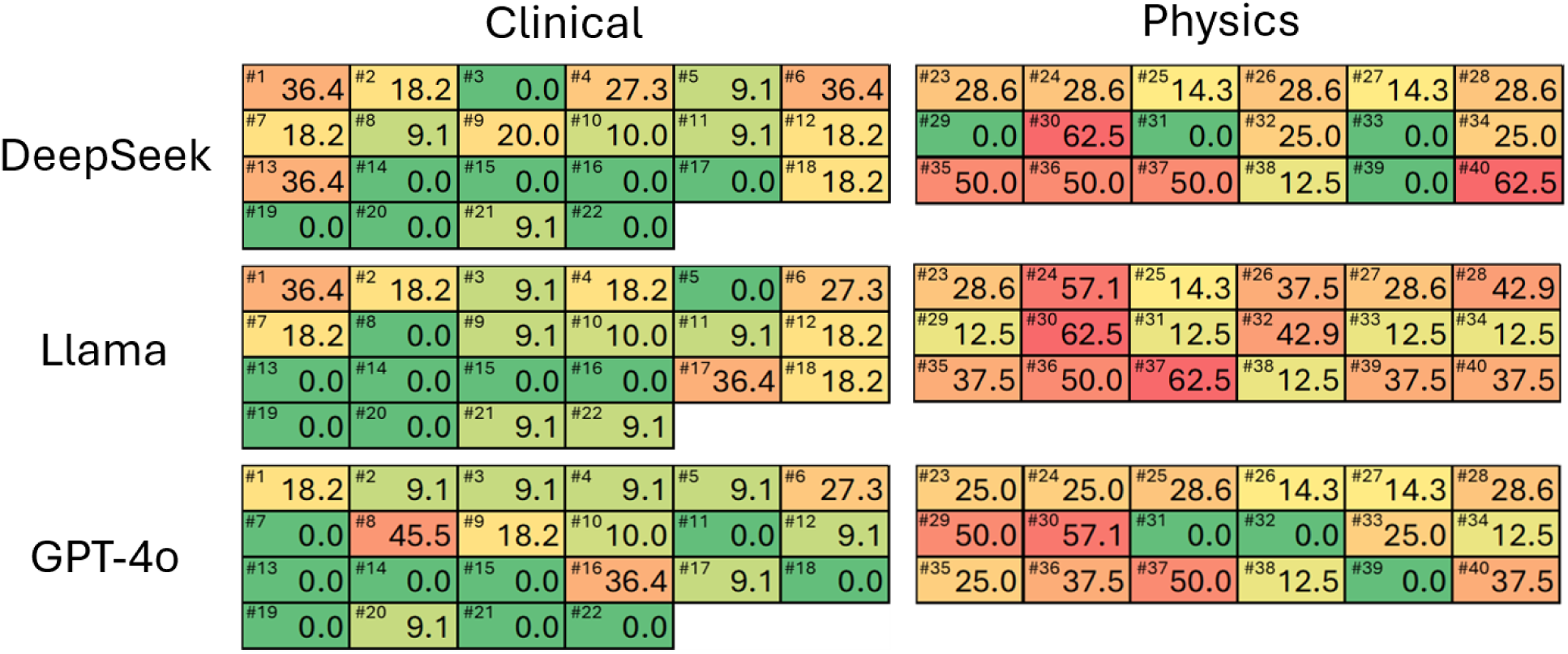
Proportion of experts indicating that an answer given by an LLM may be potentially harmful if used for clinical decision-making. Color-coding reflects the percentage of indications, ranging from green (lower percentage) to red (higher percentage).

**Figure 5:**
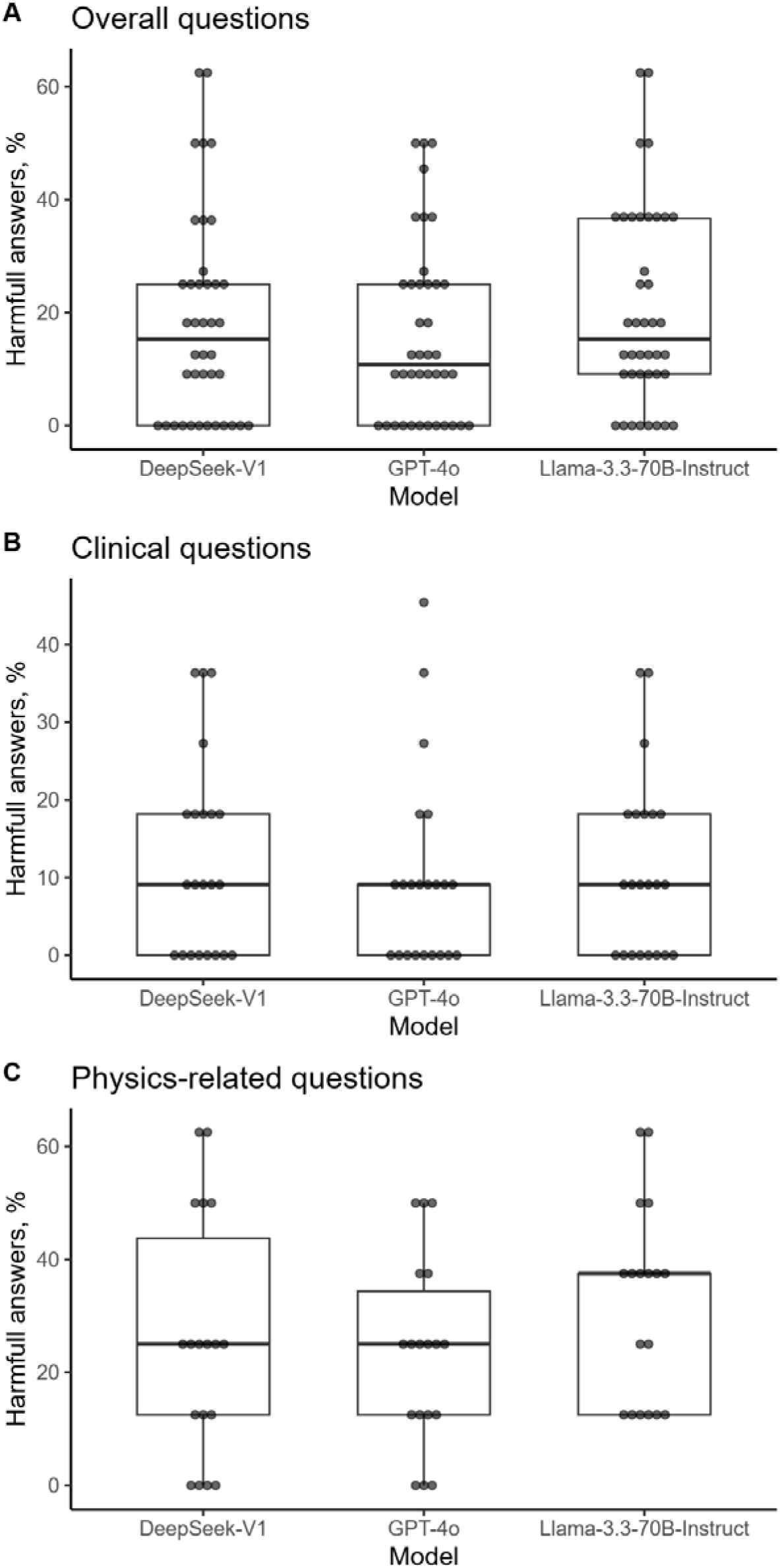
Box plots of the proportion of experts indicating potential harmfulness of responses. (A) Overall questions. (B) Clinical questions. (C) Physics-related questions. All p > 0.05 (Wilcoxon signed-rank test with false discovery rate correction for multiple comparisons). Each point represents the proportion of experts rating a response as potentially harmful for a given question.

### Examples of Questions

#### Example of «good» answers

One exemplary question with two good answers is the clinical Question 16: «A patient presents with a 10 cm solitary plasmacytoma located in the iliac bone. Is HT a beneficial adjunct to definitive RT in this setting?”

The answers from DeepSeek and Llama can be summarized as follows (full answers provided in the **Supplementary Table S1**):

*Definitive RT is the primary treatment, and plasmacytomas are generally considered radiosensitive, meaning they respond well to RT, with doses typically ranging from 40–50 Gy and are achieving high local control rates (approximately 90% for solitary bone plasmacytoma). Larger lesions may have hypoxic regions that could theoretically benefit from HT, but clinical evidence is lacking. Higher RT doses (e.g., up to 50 Gy) may be considered for large lesions as an alternative strategy*.

The responses to Question 16 from DeepSeek and Llama achieved the highest ratings among all 40 questions in the study, with a median score of 4 for both models and mean scores of 4.36 and 4.27, respectively (**Supplementary Table S1**). None of the reviewers classified these answers as “potentially harmful.” In contrast, GPT-4o’s response was rated as “bad,” with the worst mean score (2.18) among all clinical responses, while simultaneously showing one of the highest levels of interrater agreement. In summary, it described HT and its mechanism of action as a radiosensitizer and then stated that “the use of HT as an adjunct therapy should be considered on a case-by-case basis”. This response was deemed “potentially harmful” by 4 of 11 reviewers (36.4%; **Figure 4**), because the LLM’s response could be interpreted as endorsing the addition of HT to RT as a viable therapeutic strategy.

DeepSeek and Llama seemed to correctly recognize that there are no available data supporting the use of HT in hematologic malignancies or plasmacytomas[36]. However, a phase III randomized trial has investigated the combination of RT and HT in large bone metastases and showed superiority in the primary endpoint of complete pain remission in the HT arm[37]. Importantly, neither DeepSeek nor Llama fell into this conceptual trap by misclassifying solitary plasmacytoma as a bone metastasis. Instead, for a tumor known to be radiosensitive, the LLMs appropriately recommended higher RT doses[38] rather than the addition of HT without supporting evidence, which is clinically appropriate.

#### Example of «bad» answers

One exemplary question with three bad responses and the worst rating of all responses is physics Question 30: “Provide a list of commercially available devices for superficial and deep HT therapy?”

For this question, a clearly correct and objective “ground truth” exists, although the relevant information is dispersed across the internet. The overall performance of all three LLMs was “bad” with ratings for DeepSeek (mean 2.38, median 2; classified from 5/8 reviewer [62.5%] as potentially harmful), Llama (mean 2.38, median 2; 5/8 [62.5%] potentially harmful), and GPT-4o (mean 1.86, median 2; 4/7 [57.1%] potentially harmful) (**Figure 4, Supplementary Table S2**). The evaluation of the GPT-4o response yielded the lowest score in the study, while simultaneously demonstrating the highest interrater reliability for this specific question-response pair (DeepSeek r_WG_=0.58, Llama r_WG_=0.29, GPT-4o r_WG_=0.93).

#### Example of “challenging” responses

An example of a question highlighting the current challenges in evaluating responses generated by LLMs in the field of HT is clinical Question 8: “A patient with locally advanced cervical carcinoma (LACC) is unable to receive concomitant radiosensitizing ChT. She is being treated with HT as a radiosensitizer. Should HT be administered once or twice weekly during definitive RT?”

DeepSeek cited a landmark study by the Dutch Deep HT Group[39, 40] and arrived at the correct recommendation of once-weekly HT. However, this conclusion was supported by several inaccuracies: (i) it referenced an incorrect, hallucinated study name (“HYPO” study); (ii) it cautioned against increased toxicity associated with twice-weekly HT, an assertion that contradicts evidence from randomized trials using twice-weekly HT[41, 42]; and (iii) it ultimately justified the once-weekly recommendation by citing a non-specific and non-existent guideline from ESHO. The response received a median rating of 3 and a mean rating of 3.36, with one reviewer classifying it as “potentially harmful.” Interrater reliability was low (r_WG_=0.57).

LIama appeared to rely primarily on rational argumentation without referencing any supporting sources. Despite a very lengthy response, it ultimately failed to provide a concrete answer to the question. Interestingly, the experts nevertheless assigned a favorable rating to this response. The overall rating was a median score of 4 and a mean score of 3.82, but with a low interrater reliability (r_WG_=0.52).

GPT-4o provided an incorrect recommendation by suggesting twice-weekly HT, despite the fact that the majority of relevant studies employed a once-weekly treatment schedule. This response received a median rating of 2 and a mean of 2.27 and was classified as “potentially harmful” by 5 of 11 reviewers (45%) with strong within-group interrater agreement (r_WG_=0.79).

The standard of care for LACC is definitive RT with concomitant ChT. If a patient is not eligible for ChT as radiosensitizer, HT may be used as an alternative; this approach is endorsed by Dutch[43, 44], German[45] and Swiss clinical guidelines[30]. However, these guidelines do not provide detailed recommendations regarding the practical aspects of HT administration, such as treatment frequency.

This represents a clinically relevant scenario in which a physician must determine the weekly frequency of HT treatments administered concurrently with RT without being familiar with the existing evidence. To avoid conducting a comprehensive literature review, the physician might consult an LLM. The detailed responses generated by the three LLMs are provided in **Supplementary Table S1.**

**Supplementary Table S3.** summarizes selected data from eight published randomized controlled trials serving as “ground truth”. Four of these randomized trials compared an experimental RT+HT arm with standard RT alone, whereas the other four investigated different treatment schedules combining HT, ChT, and RT. In addition, several systematic reviews, meta-analyses, and network meta-analyses have been published[46–48] and could have been incorporated into LLM training.

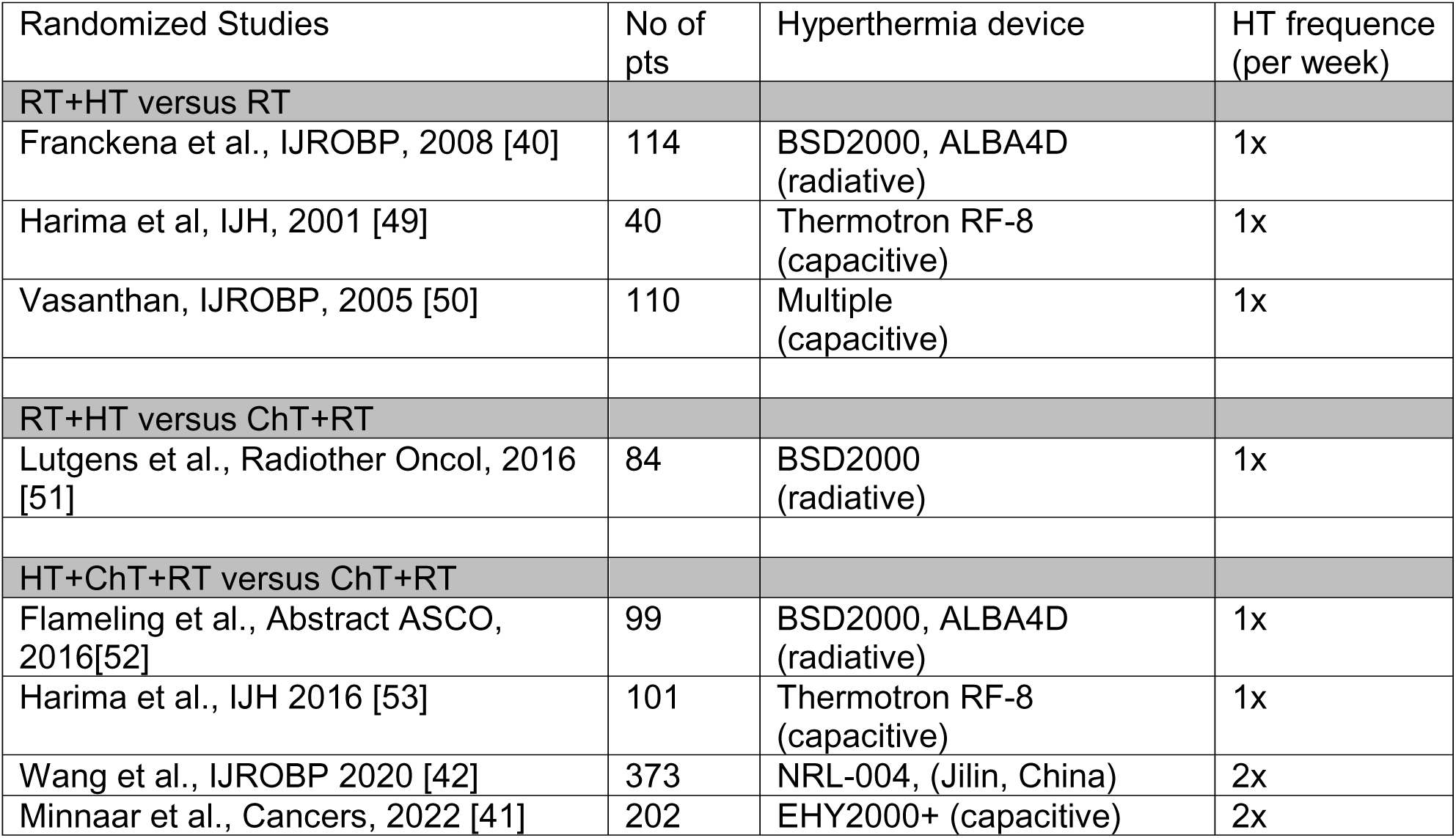

**Supplementary Table S3.** summarizes data from eight published randomized controlled trials using either concurrent radiation and hyperthermia (RT+HT) or concurrent chemoradiation and hyperthermia (ChT+RT+HT) serving as the “ground truth”. In Question 8, the frequency of hyperthermia administered concurrently with radiotherapy in patients unable to tolerate chemotherapy is explicitly addressed. The table shows that, in studies investigating this combination, hyperthermia was administered once per week.

Abbreviation: IJH, International Journal of Hyperthermia; IJROBP, International Journal of Radiation Oncology, Biology, Physics; ASCO, American Society of Clinical Oncology

## DISCUSSION

To the best of our knowledge, this is the first study investigating the capabilities of LLMs in answering questions in the field of HT. In contrast to comparable oncology studies that typically evaluate a single LLM, we assessed three models simultaneously to enable direct comparison[23, 24]. Our study found that the three LLMs—DeepSeek, Llama, and GPT-4o—in their most recent versions at the time of the study, performed at a similar, merely “acceptable” level. The mean quality scores were 3.26 for DeepSeek, 3.18 for Llama, and 3.07 for GPT-4o. The LLM DeepSeek achieved the highest score, although the difference was not statistically significant. While an overall rating of “acceptable” may not appear alarming at first glance, a disconcerting finding was that all three models generated responses rated as “bad” or “very bad” in approximately one quarter of cases (**Figure 2**). Furthermore, a substantial proportion of responses—ranging from 15.3% (GPT-4o) to 19.3% (Llama)—were flagged as potentially harmful if used in clinical decision-making. From the perspective of a user without expertise in HT, this performance is not satisfactory. In approximately one out of four questions, an LLM user would receive a “bad” or “very bad” response that could be harmful if applied in clinical practice. Without access to domain experts, users would not be able to reliably identify flawed or unsafe recommendations on their own.

In this context, it is relevant to consider how LLMs perform in comparable evaluations in other oncological subspecialties. A recent multicenter study by some of the authors of the present work evaluated 50 radiation oncology questions using the medically fine-tuned Llama3-OpenBioLLM-70B model, with responses assessed by 20 clinical reviewers. The model achieved a mean quality score of 3.38 on a 5-point Likert scale (slightly higher than DeepSeek in our study) while 16% of responses were flagged as potentially harmful in clinical decision-making [23]. These findings are broadly comparable to our results. Although a direct comparison is methodologically limited due to differences in the evaluated LLMs, the focus of the questions, oncologic subspecialties, and reviewer backgrounds, such a comparison nevertheless helps to contextualize our findings.

After the initial descriptive analysis of all questions and ratings, a detailed review of selected representative questions provided deeper insight into potential strengths and weaknesses of the LLMs.

Question 16 served as an example of a high-quality response: two LLMs generated detailed and nuanced answers that received the best expert ratings. Interestingly, the third model produced the lowest-rated response within the clinical section to the same question, accompanied by one of the highest levels of interrater reliability. This striking contrast within a single question highlights the potential for substantial differences in performance between LLMs and suggests that outputs may vary considerably across models. However, comparing responses from multiple LLMs does not by itself resolve the problem, because non-expert users may still be unable to determine which response is more accurate or clinically appropriate.

Regarding Question 30, none of the LLMs was able to generate a comprehensive list of commercially available devices for superficial and deep HT, despite the fact that these devices are described in published studies and on manufacturer websites. This is notable because screening and summarizing large amounts of information would typically be considered a strength of LLMs. In contrast, a domain expert completed this task within minutes.

One explanation may be that LLM performance is limited in highly specialized fields such as moderate HT, where the availability of structured, high-quality training data is comparatively low[6]. Compared with HT, there are substantially more publications on systemic therapies and RT. Moreover, HT is either not addressed in major international oncology guidelines or included only for selected indications, reducing the availability of standardized, guideline-based information for LLM training. In data-sparse domains such as HT, LLMs may be more prone to generating plausible but incorrect statements (“hallucinations”)[21, 22], which may be difficult for non-experts to detect. An example is the DeepSeek response to Question 8, where incorrect information was generated instead of citing available randomized trials, systematic reviews, and meta-analyses documented by the authors (**Supplementary Table S3**).

We hypothesize that the apparent “scarcity” of HT data may reflect limited visibility or accessibility rather than a true absence of evidence or data.

It is currently unresolved how to best evaluate the performance of LLMs in general, but particularly for their use in medicine[54]. Several approaches have been proposed, including classical examinations, Elo-based systems, or logical benchmarks[20, 55]. However, methodological challenges are inherent to studies of this nature. Our core group had already practical experience in investigating LLMs in medical domains and was able to rely on a well-established methodology[23, 24]. The foundation of such a study lies in the selection of the questions. Open-ended questions are certainly important for conducting clinical evaluations as they allow for a more realistic and comprehensive assessment of a model’s clinical reasoning and communication abilities. Questions from clinical practice are also open-ended and not exam-style multiple choice questions used in many medical benchmarks. However, this focus on real-world relevance introduces challenges for objective analysis.

The study itself was based on a limited number of 40 questions, divided into two subgroups: “clinical” and “physics”. Although we aimed to cover a range of topics, this sample may not represent the entire spectrum of clinical and physics-related challenges in moderate HT, which theoretically could limit the generalizability of our findings.

The level of agreement between experts was “moderate” for clinical responses and “moderate” to “good” for physics-related assessments. The moderate interrater agreement (IRA) in the clinical domain was surprising and could be considered a limitation. However, we do not assume that this was primarily due to a lack of consensus among the experts. The reviewer panel was deliberately selected from the Swiss HT Network and from active members of ESHO to maximize domain-specific consensus. In particular, most physics experts were co-authors of the ESHO clinical guidelines responsible for standardizing the technical application of HT which might be the reason for the moderate to good agreement. This approach was intended to minimize the possibility that heterogeneity in reviewer backgrounds and understanding of HT would bias the evaluation toward poorer perceived LLM performance. Instead, the moderate agreement observed in the clinical group likely reflects the inherently subjective nature of evaluating clinical responses and the complex and sometimes convoluted structure of LLM-generated answers. The DeepSeek response to Question 8 illustrates this difficulty. Although the question had a clear literature-based ground truth (**Supplementary Table S3**), the response—while professionally written and ultimately arriving at the correct recommendation—contained flawed reasoning, hallucinated references, and factual inaccuracies, including the incorrect claim that twice-weekly HT increases toxicity. Consequently, expert ratings varied widely, ranging from “bad” and “potentially harmful” to “good” and “very good.” Such combinations of correct conclusions with embedded errors complicate consistent quality assessment, as reviewers may weigh different aspects of the response differently.

A further relevant example is the evaluation of the Llama response to clinical Question 12, where the within-group interrater reliability was 0, indicating essentially no agreement (**Supplementary Table S1**).

Ratings span the full Likert scale from 1 to 5. While the response contains several strong sub-answers, it is striking that re-irradiation combined with HT is described as a “relative contraindication.” This is surprising, given that HT has been specifically used and evaluated in studies in the context of re-irradiation[56–58]. It is plausible that this particular sub-answer led some reviewers to assign a Likert score of 1 and classify the response as “potentially harmful,” despite the overall high quality of the other sub-answers (**Supplementary Table S1**).

We believe that lengthy responses containing multiple sub-arguments likely contributed to reduced interrater agreement and may have influenced response ratings in some cases. In practice, this limitation might be mitigated through iterative interaction with the LLM, where an initially non-specific response could be clarified through follow-up questions, as would occur in real-world use.

In retrospect, we acknowledge that the capabilities of LLMs may have been overestimated at the outset of the study, and that some real-world questions—although scientifically and methodologically appropriate—may have exceeded the current capabilities of these systems. This limitation became apparent only during the in-depth analysis. The use of more closed-ended question formats might have facilitated clearer and more consistent responses that were easier to evaluate. Furthermore, LLM response length was not constrained, resulting in lengthy and sometimes convoluted outputs containing multiple sub-responses. This complicated the evaluation process, as reviewers had to decide which components of a response should determine the overall rating. We assume that this factor contributed substantially to interrater variability.

A major limitation is that overall response quality was captured using a single Likert item, although quality in this context is multidimensional and encompasses factual accuracy, completeness, clarity, clinical usefulness, and safety. As a result, raters may have weighed these attributes differently, which may have contributed to the only moderate interrater agreement observed. However, Likert scale–based assessment is well established, and even this review proved highly time-consuming. A multidimensional assessment across five dimensions would have resulted in 330 and 270 individual ratings, respectively, and would therefore not have been feasible.

Despite these limitations, our study provides a valuable and timely snapshot of current LLM performance in addressing real-world questions in a highly specialized medical field.

## CONCLUSIONS

Overall, the quality of responses generated by modern LLMs to HT questions was only “acceptable,” with approximately 25% of answers rated as “bad” or “very bad” and some considered potentially harmful by international domain experts. Although two of the evaluated LLMs received ratings of “good” to “very good” for about 45% of their responses, this level of performance remains insufficient for reliable use in a clinical setting. A contributing factor appears to be the perceived scarcity of HT data. We suggest, however, that the problem is less a true lack of available HT evidence than a lack of sufficiently structured data for effective LLM training and retrieval.

Based on the present findings, the use of LLMs to answer HT-related questions cannot currently be recommended, particularly for users without domain-specific expertise. At present, general-purpose LLMs may be useful for broad orientation within the field, but they cannot be relied upon for clinically consequential or technically specific HT questions without expert oversight. This is particularly relevant in HT, where the situations in which clinicians might benefit most from AI support are often those in which current models are least reliable.

Ultimately, increased evidence generation and the development of standardized treatment guidelines in HT are likely to improve the data available for LLM training and retrieval. Improved structuring of HT knowledge, stronger guideline representation, and ongoing advances in LLM development may further enhance the reliability and future clinical utility of LLMs in moderate HT.

## Data Availability

All the question-answer pairs with the results from the evaluation are provided in the Supplementary Material (Supplementary Table S1 and S2). The source code used to answer the questions by the three LLMs is publicly available on GitHub.

https://github.com/f-dennstaedt/systematic-evaluation-of-three-llms-in-oncological-hyperthermia

## List of abbreviations

AI: Artificial Intelligence
ChT: Chemotherapy
ESHO: European Society for Hyperthermic Oncology
Gy: Gray
HT: Hyperthermia
ICC: Intraclass Correlation Coefficient
IRA: Interrater Agreement
ISROI: International Society for Radiation Oncology Informatics
LACC: Locally Advanced Cervical Carcinoma
LLM: Large Language Model
NCCN: National Comprehensive Cancer Network
RT: Radiotherapy
STM: Society for Thermal Medicine

## Funding details

No funding was received for this study.

## Disclosure statement

Nikola Cihoric serves as Technical Lead for the SmartOncology project - the open-source online platform used to collect the evaluations of the domain experts in this study - and as a medical advisor for Wemedoo AG (Steinhausen, Switzerland). The other co-authors declare no disclosures directly related to the content of this study.

## Declaration of use of Generative Artificial Intelligence (AI)

In this project, the performance of three LLMs in the field of HT was investigated. The use of LLMs constituted an integral part of the study; details regarding the specific models and their application are described in the Methods section under “Phase 2: Response Generation by Three LLMs.” In addition, GPT-5.3 was used exclusively for language improvement during the preparation of this manuscript. The authors take full accountability and responsibility for the content of this article.

## Data availability statement and data deposition

All the question-answer pairs with the results from the evaluation are provided in the Supplementary Material (**Supplementary Table S1 and S2**). The source code used to answer the questions by the three LLMs is publicly available on GitHub[29].

## Supplemental online material

- Supplementary Table S1
- Supplementary Table S2
- Supplementary Table S3

