## Supplementary Files for "A Systematic Performance Evaluation of Three Large Language Models in Answering Questions on moderate Hyperthermia"

**Supplementary Tables**

Supplementary Table S1.

**Clinical questions, responses and ratings:** For each of the 22 clinical questions posed to the three large Language Models (LLMs), the three LLM responses are presented side by side for comparison. The left column shows DeepSeek-V3, the middle column Llama-3.3-70B-Instruct, and the right column GPT-4o. The question number is shown in the upper left corner in each case. Subsequently, the ratings of the clinical reviewers are listed for each question and for each LLM. The reviewers are anonymized as C1 – C11. For the Likert scale rating, the quality of each individual response was assessed using a 5-point Likert scale (1 = very bad, 2 = bad, 3 = acceptable, 4 = good, 5 = very good). The Intraclass Correlation Coefficient (ICC) measures interrater agreement and reliability, with values interpreted as poor (<0.5), moderate (0.5-0.75), good (0.75-0.9), and excellent (>0.90). Furthermore, the experts indicated whether they believed an answer could be potentially harmful if used in clinical decision-making. If yes, this is marked with a ‘y = yes’ and highlighted in red.

| 1 | Hyperthermia has various temperature-dependent targets. Which is the most relevant for the clinical effect of hyperthermia? | | |
| --- | --- | --- | --- |
|  | DeepSeek-V3 | Llama-3.3-70B-Instruct | GPT-4o |
|  | The most clinically relevant temperature-dependent target of hyperthermia is **protein denaturation and cellular membrane disruption**, particularly in the range of **41-47°C**. This range is critical because: 1. **Direct Cytotoxicity (42-45°C)**:   - Hyperthermia induces protein unfolding, misfolding, and aggregation, leading to **loss of enzyme function** and **disruption of cellular homeostasis**.   - **Membrane fluidity increases**, compromising integrity and leading to ion imbalance and cell death.  2. **Synergy with Radiation and Chemotherapy**:   - At **41-43°C**, hyperthermia enhances tumor oxygenation and blood flow, improving radiation sensitivity.   - It also inhibits DNA repair mechanisms (e.g., homologous recombination), making cancer cells more vulnerable to chemo/radiotherapy.  3. **Immunogenic Effects**:   - Heat shock proteins (HSPs, e.g., HSP70) released at **42-45°C** trigger **anti-tumor immune responses** by promoting antigen presentation.  4. **Vascular Damage (43-45°C)**:   - Prolonged heating causes **tumor vasculature collapse**, leading to ischemia and necrosis.  ### Clinical Relevance:  - **Moderate hyperthermia (40-44°C)** is commonly used in oncology due to its balance between efficacy and safety.  - **Higher temperatures (>50°C)**, used in ablation (e.g., radiofrequency ablation), cause immediate coagulative necrosis but are less selective.  Thus, **protein denaturation and membrane disruption in the 41-47°C range** are central to hyperthermia is clinical effects, especially when combined with other therapies. | Hyperthermia, or elevated body temperature, can have various effects on the body, depending on the temperature and duration of exposure. The most relevant temperature-dependent target for the clinical effect of hyperthermia is likely to be the proteins and cellular membranes. At temperatures between 40°C to 45°C (104°F to 113°F), hyperthermia can cause: 1. **Protein denaturation**: Proteins can unfold and lose their native conformation, leading to changes in their function and activity. This can affect various cellular processes, including enzyme activity, protein-protein interactions, and cell signaling. 2. **Cell membrane fluidity**: Hyperthermia can alter the fluidity of cell membranes, affecting the movement of molecules and ions across the membrane. This can lead to changes in cellular transport, signaling, and communication. 3. **DNA damage**: Elevated temperatures can cause DNA damage, including strand breaks and mutations, which can lead to cell death or altered cellular behavior. In the context of cancer treatment, hyperthermia is often used in combination with radiation or chemotherapy to enhance the effectiveness of these therapies. The temperature-dependent targets in this context are likely to be: 1. **Tumor cells**: Hyperthermia can directly kill tumor cells or make them more sensitive to radiation and chemotherapy. 2. **Tumor vasculature**: Hyperthermia can damage the blood vessels within tumors, reducing blood flow and oxygen delivery, which can enhance the effectiveness of radiation and chemotherapy. 3. **Immune system**: Hyperthermia can stimulate the immune system, enhancing the body's natural anti-tumor response. The optimal temperature for hyperthermia in cancer treatment is typically between 40°C to 43°C (104°F to 109.4°F), as this range can induce significant tumor cell killing while minimizing damage to normal tissues. In summary, the most relevant temperature-dependent targets for the clinical effect of hyperthermia are proteins, cellular membranes, and DNA, which can lead to changes in cellular function, communication, and behavior. In the context of cancer treatment, the targets are tumor cells, tumor vasculature, and the immune system. | Hyperthermia, which involves raising the temperature of body tissues, has several temperature-dependent targets that contribute to its clinical effects. The most relevant target for the clinical effect of hyperthermia is typically the enhancement of tumor cell sensitivity to radiation and certain chemotherapeutic agents. This is primarily achieved through: 1. **Protein Denaturation and Aggregation**: At elevated temperatures, proteins can denature and aggregate, disrupting cellular functions and leading to cell death. This effect is more pronounced in cancer cells due to their altered protein homeostasis. 2. **Increased Blood Flow and Oxygenation**: Hyperthermia can increase blood flow to the tumor, improving oxygenation. This is particularly important in radiotherapy, as oxygenated cells are more susceptible to radiation damage. 3. **Inhibition of DNA Repair**: Hyperthermia can impair the ability of cancer cells to repair DNA damage, which is particularly beneficial when combined with treatments like radiation that induce DNA damage. 4. **Immune Modulation**: Hyperthermia can stimulate the immune system, enhancing the recognition and destruction of cancer cells. Among these, the enhancement of tumor cell sensitivity to radiation and chemotherapy through mechanisms like impaired DNA repair and increased oxygenation is often considered the most clinically relevant effect of hyperthermia. This synergistic effect can lead to improved treatment outcomes in cancer therapy. |
| 2 | What is the target intratumoral temperature that should be achieved during treatment? | | |
|  | DeepSeek-V3 | Llama-3.3-70B-Instruct | GPT-4o |
|  | The target **intratumoral temperature** during **hyperthermia treatment** typically falls within the range of **40-45°C (104-113°F)**. This range is chosen because: 1. **Cytotoxic Effects (42°C)**:   - Temperatures above **42°C** induce direct tumor cell death via protein denaturation, membrane disruption, and inhibition of DNA repair mechanisms.   - Prolonged exposure (30-60 minutes) enhances thermal damage. 2. **Synergy with Radiation/Chemotherapy**:   - Mild hyperthermia (40-43°C) improves blood flow and oxygen supply, sensitizing tumors to **radiation therapy**.   - It also enhances drug delivery and efficacy of **chemotherapy** (e.g., cisplatin, doxorubicin).  3. **Avoiding Normal Tissue Damage**:   - Temperatures **>45°C** risk excessive damage to healthy tissues, while **<40°C** may be insufficient for therapeutic effects.  ### Clinical Applications:  - **Local Hyperthermia**: Focused heating (e.g., via RF, microwave, or ultrasound) targets tumors directly.  - **Whole-Body Hyperthermia**: Rare, used for metastatic disease (typically ‰¤42°C).  ### Key Considerations:  - **Temperature uniformity** is critical to avoid under-treated regions.  - **Real-time monitoring** (e.g., thermocouples, MRI thermometry) ensures precision.  For **thermal ablation** (e.g., radiofrequency ablation), much higher temperatures (**50-100°C**) are used to induce immediate coagulative necrosis.  Would you like details on a specific cancer type or heating technique? | The target intratumoral temperature that should be achieved during treatment, such as hyperthermia or thermal ablation, depends on the specific treatment modality and the type of cancer being treated. However, here are some general guidelines: **Hyperthermia:** * Mild hyperthermia: 39°C to 40°C (102°F to 104°F) for 30-60 minutes * Moderate hyperthermia: 40°C to 42°C (104°F to 108°F) for 30-60 minutes * Severe hyperthermia: 42°C to 45°C (108°F to 113°F) for shorter durations (e.g., 15-30 minutes) **Thermal Ablation:** * Radiofrequency ablation (RFA): 50°C to 100°C (122°F to 212°F) for 10-30 minutes * Microwave ablation (MWA): 50°C to 100°C (122°F to 212°F) for 10-30 minutes * Laser-induced thermal therapy (LITT): 50°C to 100°C (122°F to 212°F) for 10-30 minutes * Cryoablation: -20°C to -40°C (-4°F to -40°F) for 10-30 minutes **Note:** * The target temperature may vary depending on the specific tumor type, size, and location. * The treatment duration and temperature may be adjusted based on the patient's response and the tumor's thermal sensitivity. * It's essential to monitor the temperature in real-time during treatment to avoid overheating or underheating, which can lead to adverse effects or reduced treatment efficacy. It's crucial to consult with a qualified healthcare professional or a medical physicist to determine the optimal treatment parameters for a specific patient and tumor type. | Hyperthermia, or elevated body temperature, can have various effects on the body, depending on the temperature and duration of exposure. The most relevant temperature-dependent target for the clinical effect of hyperthermia is likely to be the proteins and cellular membranes. At temperatures between 40°C to 45°C (104°F to 113°F), hyperthermia can cause: 1. **Protein denaturation**: Proteins can unfold and lose their native conformation, leading to changes in their function and activity. This can affect various cellular processes, including enzyme activity, protein-protein interactions, and cell signaling. 2. **Cell membrane fluidity**: Hyperthermia can alter the fluidity of cell membranes, affecting the movement of molecules and ions across the membrane. This can lead to changes in cellular transport, signaling, and communication. 3. **DNA damage**: Elevated temperatures can cause DNA damage, including strand breaks and mutations, which can lead to cell death or altered cellular behavior. In the context of cancer treatment, hyperthermia is often used in combination with radiation or chemotherapy to enhance the effectiveness of these therapies. The temperature-dependent targets in this context are likely to be: 1. **Tumor cells**: Hyperthermia can directly kill tumor cells or make them more sensitive to radiation and chemotherapy. 2. **Tumor vasculature**: Hyperthermia can damage the blood vessels within tumors, reducing blood flow and oxygen delivery, which can enhance the effectiveness of radiation and chemotherapy. 3. **Immune system**: Hyperthermia can stimulate the immune system, enhancing the body's natural anti-tumor response. The optimal temperature for hyperthermia in cancer treatment is typically between 40°C to 43°C (104°F to 109.4°F), as this range can induce significant tumor cell killing while minimizing damage to normal tissues. In summary, the most relevant temperature-dependent targets for the clinical effect of hyperthermia are proteins, cellular membranes, and DNA, which can lead to changes in cellular function, communication, and behavior. In the context of cancer treatment, the targets are tumor cells, tumor vasculature, and the immune system. |
| 3 | Hyperthermia acts as a radiosensitizer in malignant cells. Does it also exert a radioprotective effect on normal tissues? | | |
|  | DeepSeek-V3 | Llama-3.3-70B-Instruct | GPT-4o |
|  | Hyperthermia (typically 40-45°C) is well-established as a **radiosensitizer** in malignant cells, enhancing the effects of radiation therapy by increasing oxygen radical production, impairing DNA repair, and disrupting tumor vasculature. However, its impact on **normal tissues** is more nuanced and context-dependent: ### **Does Hyperthermia Protect Normal Tissues from Radiation?** 1. **Possible Protective Mechanisms**   - **Thermotolerance**: Normal tissues can develop transient thermotolerance (via heat shock proteins like HSP70), potentially reducing heat-induced damage.   - **Differential Sensitivity**: Some normal tissues (e.g., muscle, fat) are less heat-sensitive than tumors, allowing selective radiosensitization of malignant cells.   - **Vascular Effects**: While hyperthermia disrupts tumor vasculature, normal tissue vasculature may recover better, reducing radiation-induced damage.  2. **Evidence of Radioprotection**   - Some preclinical studies suggest that **mild hyperthermia (‰¤ 41°C)** before radiation may upregulate protective heat shock proteins in normal tissues, reducing apoptosis and inflammation.   - However, **higher temperatures (> 42°C) can exacerbate radiation damage** in normal tissues if not carefully controlled.  3. **Clinical Observations**   - In clinical settings, hyperthermia is often **spatially targeted** (e.g., using focused ultrasound or RF heating) to minimize normal tissue exposure.   - Some studies report **reduced late radiation toxicity** (e.g., fibrosis) when hyperthermia is applied with precise temperature control.  ### **Conclusion**  Hyperthermia **does not universally protect normal tissues** from radiation, but under controlled conditions (lower temperatures, proper timing, and selective application), it may **reduce radiation-induced damage** in some normal tissues while enhancing tumor radiosensitization. The effect depends on:  - **Temperature and duration** (mild hyperthermia may be protective, while higher heat can be harmful).  - **Tissue type** (well-vascularized normal tissues may recover better).  - **Timing** (preconditioning with heat before radiation may induce protective responses).  Further research is needed to optimize hyperthermia protocols for **differential radioprotection** of normal tissues alongside tumor radiosensitization. | Hyperthermia, or elevated temperature, is known to act as a radiosensitizer, meaning it can enhance the effectiveness of radiation therapy in killing malignant cells. This is because hyperthermia can increase the damage caused by radiation to the DNA of cancer cells, making them more susceptible to death. However, the effect of hyperthermia on normal tissues is more complex. While hyperthermia can indeed exert a radioprotective effect on some normal tissues, the extent and mechanisms of this effect are still being researched and debated. Some studies suggest that hyperthermia can induce a protective response in normal tissues, such as: 1. **Heat shock protein (HSP) induction**: Hyperthermia can induce the expression of HSPs, which can help protect normal cells against radiation-induced damage. 2. **Increased DNA repair**: Hyperthermia can stimulate DNA repair mechanisms in normal cells, making them more resistant to radiation-induced DNA damage. 3. **Improved tissue perfusion**: Hyperthermia can increase blood flow and oxygen delivery to normal tissues, which may help mitigate radiation-induced damage. However, other studies have reported that hyperthermia can also exacerbate radiation damage in normal tissues, particularly at higher temperatures (>42°C) or with prolonged exposure. This is because hyperthermia can: 1. **Increase oxidative stress**: Hyperthermia can lead to the production of reactive oxygen species (ROS), which can damage normal cells and exacerbate radiation-induced damage. 2. **Disrupt tissue architecture**: Hyperthermia can cause changes in tissue structure and function, making normal cells more vulnerable to radiation damage. The radioprotective effect of hyperthermia on normal tissues is likely to depend on various factors, including: 1. **Temperature and duration**: The temperature and duration of hyperthermia exposure can influence the balance between radioprotection and radiosensitization. 2. **Tissue type**: Different normal tissues may respond differently to hyperthermia, with some being more resistant to radiation damage than others. 3. **Dose and schedule of radiation**: The dose and schedule of radiation therapy can impact the effectiveness of hyperthermia as a radioprotector or radiosensitizer. In summary, while hyperthermia can exert a radioprotective effect on normal tissues under certain conditions, the relationship between hyperthermia, radiation, and normal tissue damage is complex and requires further research to fully understand its mechanisms and optimize its therapeutic applications. | Hyperthermia, or elevated body temperature, can have various effects on the body, depending on the temperature and duration of exposure. The most relevant temperature-dependent target for the clinical effect of hyperthermia is likely to be the proteins and cellular membranes. At temperatures between 40°C to 45°C (104°F to 113°F), hyperthermia can cause: 1. **Protein denaturation**: Proteins can unfold and lose their native conformation, leading to changes in their function and activity. This can affect various cellular processes, including enzyme activity, protein-protein interactions, and cell signaling. 2. **Cell membrane fluidity**: Hyperthermia can alter the fluidity of cell membranes, affecting the movement of molecules and ions across the membrane. This can lead to changes in cellular transport, signaling, and communication. 3. **DNA damage**: Elevated temperatures can cause DNA damage, including strand breaks and mutations, which can lead to cell death or altered cellular behavior. In the context of cancer treatment, hyperthermia is often used in combination with radiation or chemotherapy to enhance the effectiveness of these therapies. The temperature-dependent targets in this context are likely to be: 1. **Tumor cells**: Hyperthermia can directly kill tumor cells or make them more sensitive to radiation and chemotherapy 2. **Tumor vasculature**: Hyperthermia can damage the blood vessels within tumors, reducing blood flow and oxygen delivery, which can enhance the effectiveness of radiation and chemotherapy. 3. **Immune system**: Hyperthermia can stimulate the immune system, enhancing the body's natural anti-tumor response. The optimal temperature for hyperthermia in cancer treatment is typically between 40°C to 43°C (104°F to 109.4°F), as this range can induce significant tumor cell killing while minimizing damage to normal tissues. In summary, the most relevant temperature-dependent targets for the clinical effect of hyperthermia are proteins, cellular membranes, and DNA, which can lead to changes in cellular function, communication, and behavior. In the context of cancer treatment, the targets are tumor cells, tumor vasculature, and the immune system. |
| 4 | What is the optimal sequence for hyperthermia in relation to radiotherapy”should hyperthermia be administered before or after irradiation? | | |
|  | DeepSeek-V3 | Llama-3.3-70B-Instruct | GPT-4o |
|  | The optimal sequence for combining hyperthermia with radiotherapy depends on the therapeutic goals, biological mechanisms, and practical considerations. Generally, **administering hyperthermia before radiotherapy (pre-irradiation hyperthermia) is considered more effective** for several reasons: ### **1. Biological Rationale for Pre-Irradiation Hyperthermia**  - **Radiosensitization**: Hyperthermia (typically 40-44°C) enhances tumor oxygenation and disrupts DNA repair mechanisms, making cancer cells more susceptible to subsequent radiation damage.  - **Inhibition of DNA Repair**: Heating cells before radiation impairs their ability to repair sublethal and potentially lethal damage induced by radiotherapy.  - **Vascular Effects**: Pre-heating can improve tumor perfusion, reducing hypoxia (a major cause of radioresistance) before irradiation.  ### **2. Post-Irradiation Hyperthermia Considerations**  - While hyperthermia after radiation can still enhance cytotoxicity, it may be less effective because:   - Radiation-induced vascular damage can reduce heat distribution in the tumor.   - The primary radiosensitizing effects (e.g., oxygen enhancement) are maximized when hyperthermia precedes radiation.  ### **3. Clinical and Preclinical Evidence**  - **Preclinical studies** consistently show superior tumor control when hyperthermia is given before radiation.  - **Clinical trials** (e.g., in cervical cancer, soft tissue sarcomas, and recurrent breast cancer) often use hyperthermia **1-4 hours before radiotherapy** for optimal synergy.  ### **4. Practical Considerations**  - **Timing**: Hyperthermia is typically administered **30 minutes to 4 hours before radiotherapy** to maximize radiosensitization.  - **Temperature and Duration**: Mild hyperthermia (41-43°C) for 30-60 minutes is commonly used to avoid normal tissue damage.  ### **Conclusion**  **Hyperthermia should ideally be administered before radiotherapy** to exploit its radiosensitizing effects. However, in some cases (e.g., deep-seated tumors with heating challenges), concurrent or post-radiation hyperthermia may still provide benefits. The exact protocol should be tailored based on tumor type, location, and available hyperthermia technology.  Would you like specific references or clinical trial data on this topic? | The optimal sequence for hyperthermia in relation to radiotherapy is a topic of ongoing research and debate. The sequence of hyperthermia and radiotherapy can affect the outcome of treatment, and the ideal sequence may depend on various factors, including the type of cancer, the stage of disease, and the specific treatment goals. **Pre-irradiation hyperthermia:** Administering hyperthermia before radiotherapy (pre-irradiation hyperthermia) has been shown to enhance the radiosensitivity of tumor cells. Hyperthermia can increase blood flow and oxygenation to the tumor, making it more susceptible to radiation damage. Additionally, hyperthermia can induce changes in the tumor microenvironment, such as increased apoptosis and decreased tumor cell proliferation, which can enhance the effectiveness of subsequent radiotherapy. **Post-irradiation hyperthermia:** On the other hand, administering hyperthermia after radiotherapy (post-irradiation hyperthermia) can also be effective. Hyperthermia can enhance the repair of radiation-induced damage in normal tissues, while simultaneously increasing the cytotoxic effects of radiation on tumor cells. Post-irradiation hyperthermia can also induce a phenomenon called "thermal radiosensitization," where the heat shock proteins induced by hyperthermia can enhance the radiosensitivity of tumor cells. **Simultaneous hyperthermia and radiotherapy:** Some studies have investigated the use of simultaneous hyperthermia and radiotherapy, where hyperthermia is administered during radiotherapy. This approach can take advantage of the synergistic effects of heat and radiation on tumor cells, potentially leading to improved treatment outcomes. **Current recommendations:** Based on the available evidence, the following general recommendations can be made: 1. For superficial tumors, pre-irradiation hyperthermia may be preferred, as it can enhance the radiosensitivity of tumor cells and improve treatment outcomes. 2. For deeper-seated tumors, post-irradiation hyperthermia may be more effective, as it can enhance the repair of radiation-induced damage in normal tissues while maintaining the cytotoxic effects of radiation on tumor cells. 3. Simultaneous hyperthermia and radiotherapy may be considered for tumors that are resistant to radiation or have a high likelihood of recurrence. **Future directions:** Further research is needed to determine the optimal sequence of hyperthermia and radiotherapy for specific types of cancer and treatment scenarios. The development of new hyperthermia technologies, such as magnetic resonance-guided focused ultrasound, may also provide new opportunities for optimizing the sequence of hyperthermia and radiotherapy. In summary, the optimal sequence for hyperthermia in relation to radiotherapy depends on various factors, and both pre-irradiation and post-irradiation hyperthermia can be effective. Further research is needed to determine the best approach for specific clinical scenarios. | The optimal sequence of hyperthermia in relation to radiotherapy can depend on several factors, including the type of cancer being treated, the specific protocols being used, and individual patient considerations. However, research generally suggests that administering hyperthermia immediately before or simultaneously with radiotherapy can be beneficial. Hyperthermia, which involves raising the temperature of tumor tissues, can enhance the effects of radiotherapy by making cancer cells more sensitive to radiation. This is because hyperthermia can inhibit the repair of DNA damage caused by radiation, increase blood flow to the tumor (improving oxygenation), and potentially disrupt the tumor microenvironment. Administering hyperthermia before or during radiotherapy can maximize these effects. When hyperthermia is applied before irradiation, it can help to increase the oxygenation of the tumor, which is crucial because oxygenated cells are more susceptible to radiation damage. Simultaneous application can also be effective, as it ensures that the radiosensitizing effects of hyperthermia are present during radiation exposure. Ultimately, the decision on sequencing should be made based on clinical evidence, the specific characteristics of the tumor, and the treatment protocol being followed. It is important for clinicians to tailor the approach to the individual needs of the patient and the specifics of their condition. |
| 5 | What is the optimal sequence for hyperthermia in relation to radiotherapy” should hyperthermia be administered before or after irradiation? | | |
|  | DeepSeek-V3 | Llama-3.3-70B-Instruct | GPT-4o |
|  | The ideal interval between hyperthermia (HT) and radiotherapy (RT) depends on several factors, including the type of hyperthermia (local, regional, or whole-body), tumor characteristics, and the goal of treatment (radiosensitization vs. immune modulation). However, based on preclinical and clinical evidence, the following general principles apply ### **Optimal Timing for Radiosensitization:** 1. **Simultaneous or Immediate (<1 hour):**   - Hyperthermia is most effective as a radiosensitizer when administered **within 1 hour before or after RT**.   - Heat increases tumor oxygenation, inhibits DNA repair, and enhances radiation-induced DNA damage.   - Preclinical studies suggest the highest thermal enhancement ratio (TER) when RT is given **during or immediately after heating** (41-43°C for 30-60 min).  2. **Up to 4 Hours After Hyperthermia:**   - Some studies show persistent radiosensitization effects for **up to 3-4 hours post-HT**, but efficacy diminishes with longer delays.   - Beyond 4 hours, the thermal enhancement effect decreases due to recovery of tumor cells.  ### **For Immune Modulation (if applicable):**  - If hyperthermia is used to stimulate immune responses (e.g., via heat shock protein release), a longer interval (e.g., **24-72 hours before RT**) may be beneficial to allow immune cell activation.  ### **Clinical Evidence:** - **Head and neck, breast, and pelvic tumors:** Clinical trials often use hyperthermia **within 30-60 minutes before RT** for optimal radiosensitization.  - **Superficial tumors (e.g., melanoma, recurrent breast cancer):** Studies like the Dutch Hyperthermia Group trials used HT **immediately before RT** with improved outcomes.  - **Deep-seated tumors (e.g., cervical cancer):** Some protocols apply HT **after RT** (within 1 hour) to avoid heat-induced hypoxia.  ### **Conclusion:**  For **maximum radiosensitization**, the best interval is **<1 hour before or after RT**. If immune effects are desired, a longer delay (24-72 hours) may be considered. Clinical protocols should be tailored based on tumor type, heating technique, and treatment goals.  Would you like specific references for particular cancer types? | The ideal interval between hyperthermia and radiotherapy to optimize treatment efficacy and/or therapeutic ratio is a topic of ongoing research and debate. However, based on the current literature, here are some general guidelines: **General principles:* 1. **Simultaneous treatment**: Treating tumors with hyperthermia and radiotherapy simultaneously (i.e., during the same session) can be beneficial, as hyperthermia can enhance the radiosensitivity of tumor cells. 2. **Short interval**: A short interval (e.g., 30 minutes to 2 hours) between hyperthermia and radiotherapy can also be effective, as the tumor cells are still in a state of increased radiosensitivity after hyperthermia. 3. **Optimal interval**: The optimal interval between hyperthermia and radiotherapy is likely to depend on the specific tumor type, size, and location, as well as the hyperthermia treatment parameters (e.g., temperature, duration, and frequency). **Specific intervals:** 1. **0-30 minutes**: Simultaneous treatment or a short interval (0-30 minutes) between hyperthermia and radiotherapy has been shown to be effective in enhancing tumor response and reducing toxicity in various studies. 2. **30 minutes to 2 hours**: An interval of 30 minutes to 2 hours between hyperthermia and radiotherapy has been used in several clinical trials, with promising results. 3. **2-4 hours**: Some studies have investigated intervals of 2-4 hours between hyperthermia and radiotherapy, with mixed results. 4. **>4 hours**: Intervals greater than 4 hours between hyperthermia and radiotherapy are generally considered less effective, as the tumor cells may have recovered from the hyperthermic insult. **Tumor-specific considerations:** 1. **Superficial tumors**: For superficial tumors (e.g., skin, breast, or head and neck cancers), simultaneous treatment or a short interval (0-30 minutes) between hyperthermia and radiotherapy may be more effective. 2. **Deep-seated tumors**: For deep-seated tumors (e.g., prostate, liver, or lung cancers), a longer interval (e.g., 30 minutes to 2 hours) between hyperthermia and radiotherapy may be necessary to allow for adequate heat penetration and tumor heating. **Conclusion:** The ideal interval between hyperthermia and radiotherapy to optimize treatment efficacy and/or therapeutic ratio is likely to depend on the specific tumor type, size, and location, as well as the hyperthermia treatment parameters. While simultaneous treatment or a short interval (0-30 minutes) between hyperthermia and radiotherapy may be effective for some tumors, a longer interval (e.g., 30 minutes to 2 hours) may be necessary for others. Further research is needed to determine the optimal interval for specific tumor types and treatment protocols. | The ideal interval between hyperthermia and radiotherapy to optimize treatment efficacy and the therapeutic ratio is generally considered to be within a few hours, typically around 0 to 4 hours. Hyperthermia is known to enhance the effects of radiotherapy by increasing tumor oxygenation, inhibiting DNA repair mechanisms, and sensitizing cancer cells to radiation. The timing is crucial because the biological effects of hyperthermia, such as increased blood flow and oxygenation, are transient. Studies suggest that administering hyperthermia immediately before or after radiotherapy can maximize these synergistic effects. However, the exact timing can vary depending on the specific type of cancer, the hyperthermia technique used, and individual patient factors. Therefore, treatment protocols should be tailored to the specific clinical scenario, and ongoing research continues to refine these recommendations. It is always best to consult with a multidisciplinary team to determine the most appropriate treatment schedule for each patient. |
| 6 | In a standard hyperthermia session, a warm-up phase precedes the therapeutic phase. What is the recommended duration of the therapeutic (treatment) phase? | | |
|  | DeepSeek-V3 | Llama-3.3-70B-Instruct | GPT-4o |
|  | In a standard hyperthermia session, the **therapeutic (treatment) phase** typically lasts between **30 to 60 minutes** once the target temperature is reached. The exact duration depends on factors such as:  - **Type of hyperthermia** (local, regional, or whole-body)  - **Treatment goals** (e.g., cancer therapy, pain management, immune stimulation)  - **Patient tolerance and response**  For **local or regional hyperthermia** (e.g., for tumors), the therapeutic phase is often around **30-45 minutes** at the target temperature (usually **40-44°C**).  For **whole-body hyperthermia**, the duration may extend up to **60 minutes** at a lower target temperature (typically **39.5-41.5°C**) to avoid excessive systemic stress.  The warm-up phase (heating to target temperature) is not counted as part of the therapeutic phase and may take an additional **15-30 minutes** depending on the method used (e.g., infrared, radiofrequency, or water-filtered infrared A).  Would you like details on specific hyperthermia protocols? | The recommended duration of the therapeutic (treatment) phase in a standard hyperthermia session can vary depending on the specific treatment protocol, the type of hyperthermia being used (e.g., local, regional, or whole-body), and the individual patient's condition and response to treatment. However, based on general guidelines and clinical practices, here are some common recommendations for the duration of the therapeutic phase: * Local hyperthermia: 30-60 minutes * Regional hyperthermia: 60-90 minutes * Whole-body hyperthermia: 60-120 minutes For example, the National Cancer Institute (NCI) recommends the following treatment durations for hyperthermia: * Local hyperthermia: 30-60 minutes, with temperatures between 40°C and 45°C * Regional hyperthermia: 60-90 minutes, with temperatures between 40°C and 43°C * Whole-body hyperthermia: 60-120 minutes, with temperatures between 38.5°C and 40°C It's essential to note that these are general guidelines, and the actual treatment duration may vary depending on the specific clinical protocol, the patient's response to treatment, and the treating physician's professional judgment. The therapeutic phase should be carefully monitored to ensure that the patient's temperature is maintained within a safe and effective range, and that any potential side effects are minimized. | In a standard hyperthermia session, the therapeutic phase typically lasts between 30 to 60 minutes. This duration can vary depending on the specific protocol being used, the condition being treated, and the patient's tolerance. It's important for the session to be closely monitored by healthcare professionals to ensure safety and effectiveness. |
| 7 | To what extent do non-thermal effects of radiative hyperthermia contribute to the overall therapeutic outcome? | | |
|  | DeepSeek-V3 | Llama-3.3-70B-Instruct | GPT-4o |
|  | The non-thermal effects of radiative hyperthermia contribute significantly to the overall therapeutic outcome, though their exact extent depends on the specific modality (e.g., radiofrequency, microwave, ultrasound, or laser therapy) and the biological target. While hyperthermia is primarily known for its thermal effects (direct cell killing via protein denaturation, membrane disruption, and vascular damage), non-thermal mechanisms also play a crucial role, particularly in enhancing radiosensitivity, immunotherapy responses, and cellular stress pathways. Here is a breakdown of key contributions: ### **1. Enhanced Radiosensitization (Thermo-Radiotherapy)**  - **Non-thermal mechanisms**: Hyperthermia (even at sub-lethal temperatures, 39-42°C) can impair DNA repair pathways (e.g., inhibition of homologous recombination) and increase oxidative stress, making tumor cells more susceptible to radiation. This is not solely due to heat but also involves heat shock protein (HSP) activation and altered tumor microenvironment (TME) dynamics.  - **Clinical impact**: Studies show synergistic effects when hyperthermia is combined with radiotherapy, improving local control rates in cancers like breast, cervical, and soft-tissue sarcomas. ### **2. Immunomodulation**  - **Heat shock proteins (HSPs)**: Mild hyperthermia upregulates HSPs (e.g., HSP70), which can act as danger signals, promoting dendritic cell maturation and antigen cross-presentation, thus enhancing anti-tumor immunity.  - **T-cell infiltration**: Non-lethal heating increases blood flow and endothelial permeability, facilitating immune cell infiltration into tumors.  - **Combination with immunotherapy**: Preclinical data suggest hyperthermia potentiates checkpoint inhibitors (e.g., anti-PD-1) by reducing immunosuppressive factors (e.g., Tregs, MDSCs). ### **3. Tumor Microenvironment (TME) Modulation**  - **Vascular effects**: Sub-ablative hyperthermia can normalize tumor vasculature transiently, improving drug delivery (e.g., chemotherapy) while also inducing hypoxia-reoxygenation stress.  - **Metabolic disruption**: Non-thermal RF/microwave fields may alter ion channels and mitochondrial function, increasing reactive oxygen species (ROS) independently of bulk heating. ### **4. Direct Non-Thermal Cellular Effects**  - **Electromagnetic field interactions**: Some studies propose that oscillating fields (e.g., RF) can affect cell membrane potentials or calcium signaling, though evidence is debated.  - **Low-level photothermal effects**: Laser or near-infrared therapies may trigger photobiomodulation, influencing cellular metabolism without significant temperature rise. ### **Clinical Relevance** - **Moderate hyperthermia (40-44°C)**: Dominated by thermal effects but with notable non-thermal contributions (e.g., HSP-mediated immunogenicity). - **Mild hyperthermia (<40°C)**: Non-thermal effects (e.g., immune priming, radiosensitization) may dominate, especially in combinatorial therapies. ### **Challenges & Research Gaps** - Quantifying non-thermal effects is difficult due to overlapping mechanisms. - Some purported non-thermal phenomena (e.g., specific RF effects) lack robust clinical validation. ### **Conclusion** Non-thermal effects likely contribute **20-40%** of the therapeutic benefit in hyperthermia, particularly in immunomodulation and radiosensitization, while thermal effects remain primary for direct cytotoxicity. Their importance grows in combination therapies, where mild heating can "prime" tumors for other treatments. Further research is needed to isolate and harness these mechanisms optimally. | The extent to which non-thermal effects of radiative hyperthermia contribute to the overall therapeutic outcome is a topic of ongoing research and debate. While thermal effects are well-established and widely recognized, non-thermal effects are less well understood and their contribution to the therapeutic outcome is still being investigated. Non-thermal effects of radiative hyperthermia refer to the biological responses that occur at temperatures below the thermal threshold (typically considered to be around 42°C) or in the absence of significant temperature elevations. These effects can be induced by various forms of electromagnetic radiation, including radiofrequency (RF), microwave, and laser radiation. Several non-thermal effects have been reported, including: 1. **Cell membrane permeabilization**: Radiative hyperthermia can alter the permeability of cell membranes, allowing for the uptake of therapeutic agents or ions. 2. **Intracellular signaling**: Non-thermal radiation can activate intracellular signaling pathways, influencing gene expression, protein synthesis, and cell behavior. 3. **Reactive oxygen species (ROS) generation**: Radiative hyperthermia can induce the production of ROS, which can damage cellular components and trigger cell death. 4. **Immune system modulation**: Non-thermal radiation can stimulate the immune system, enhancing the anti-tumor response. 5. **Angiogenesis inhibition**: Radiative hyperthermia can inhibit the formation of new blood vessels, reducing tumor growth and metastasis. The contribution of non-thermal effects to the overall therapeutic outcome of radiative hyperthermia is likely to be significant, particularly in the following contexts: 1. **Low-temperature hyperthermia**: At temperatures below 42°C, non-thermal effects may dominate the therapeutic response. 2. **Pulsed radiation**: Pulsed radiation regimens, which involve brief, high-intensity radiation pulses, may induce non-thermal effects that contribute to the therapeutic outcome. 3. **Combination therapies**: Non-thermal effects may enhance the efficacy of combination therapies, such as chemotherapy, radiation therapy, or immunotherapy, when used in conjunction with radiative hyperthermia. 4. **Tumor microenvironment**: Non-thermal effects may influence the tumor microenvironment, affecting the behavior of cancer cells, immune cells, and other non-cancerous cells within the tumor. However, the extent to which non-thermal effects contribute to the therapeutic outcome is still uncertain and may depend on various factors, including: 1. **Radiation parameters**: The frequency, intensity, and duration of radiation exposure can influence the magnitude and type of non-thermal effects. 2. **Tumor type and biology**: The response to non-thermal effects may vary depending on the tumor type, stage, and genetic profile.  3. **Treatment schedule and duration**: The timing and duration of radiative hyperthermia treatments can impact the development and magnitude of non-thermal effects. In conclusion, while non-thermal effects of radiative hyperthermia are likely to contribute to the overall therapeutic outcome, their extent and significance are still being investigated. Further research is needed to fully understand the mechanisms and clinical implications of non-thermal effects in radiative hyperthermia. | Radiative hyperthermia is a treatment modality that involves raising the temperature of body tissues to enhance the therapeutic effects of other treatments, such as radiation therapy or chemotherapy. The primary mechanism of action is thermal, where increased temperatures can lead to direct cytotoxic effects on cancer cells, improved blood flow, and increased oxygenation, which can enhance the efficacy of concurrent treatments. However, non-thermal effects of hyperthermia are also thought to contribute to the overall therapeutic outcome, although their extent and mechanisms are less well understood and are an area of ongoing research. Some proposed non-thermal effects include: 1. **Cellular Stress Response**: Hyperthermia can induce a stress response in cells, leading to the expression of heat shock proteins (HSPs). These proteins can modulate immune responses and potentially enhance the recognition and destruction of cancer cells by the immune system. 2. **Immune Modulation**: Non-thermal effects may include the activation of the immune system. Hyperthermia can increase the presentation of tumor antigens and enhance the activity of immune cells, such as dendritic cells and T-cells, contributing to an anti-tumor immune response. 3. **Membrane Fluidity and Permeability**: Changes in cell membrane fluidity and permeability can occur at temperatures that do not cause direct thermal damage. This can affect drug uptake and the sensitivity of cancer cells to chemotherapy. 4. **Gene Expression**: Hyperthermia can alter the expression of various genes involved in cell survival, apoptosis, and repair mechanisms, potentially sensitizing cancer cells to other treatments. 5. **Vascular Effects**: While increased blood flow is a thermal effect, non-thermal changes in the vasculature, such as alterations in endothelial cell function and vascular permeability, may also play a role in enhancing treatment efficacy. The extent to which these non-thermal effects contribute to the overall therapeutic outcome of radiative hyperthermia is still being elucidated. They likely act synergistically with thermal effects to enhance treatment efficacy, but quantifying their individual contributions remains challenging. Further research is needed to better understand these mechanisms and to optimize hyperthermia protocols for improved clinical outcomes. |
| 8 | A patient with locally advanced cervical carcinoma is unable to receive concomitant radiosensitizing chemotherapy. She is being treated with hyperthermia as a radiosensitizer. Should hyperthermia be administered once or twice weekly during definitive radiotherapy? | | |
|  | DeepSeek-V3 | Llama-3.3-70B-Instruct | GPT-4o |
|  | For a patient with locally advanced cervical carcinoma receiving **definitive radiotherapy** with **hyperthermia as a radiosensitizer** (due to inability to tolerate concurrent chemotherapy), the optimal frequency of hyperthermia administration is typically **once weekly**.  ### Key Considerations:  1. **Evidence from Clinical Studies**:   - The **HYPRO trial** (Dutch Hyperthermia Group) and other studies have shown that **once-weekly hyperthermia** (in combination with radiotherapy) improves local control and survival in cervical cancer, with manageable toxicity.   - Twice-weekly hyperthermia has been explored but does not consistently show superior outcomes and may increase toxicity (e.g., skin burns, pain).  2. **Practical and Safety Factors**:   - Hyperthermia is logistically complex and requires specialized equipment.   - More frequent sessions (e.g., twice weekly) may lead to **cumulative thermal dose toxicity** without clear additional benefit.  3. **Current Guidelines**:   - The **European Society for Hyperthermic Oncology (ESHO)** and clinical protocols often recommend **once-weekly hyperthermia** alongside radiotherapy (5 fractions per week).  ### Conclusion:  **Once-weekly hyperthermia** is the preferred and evidence-based approach in this setting. If the tumor shows suboptimal response, individualized adjustments may be considered, but twice-weekly hyperthermia is not routinely recommended.  Would you like references to specific studies or protocols? | For a patient with locally advanced cervical carcinoma who cannot receive concomitant radiosensitizing chemotherapy and is instead being treated with hyperthermia as a radiosensitizer, the optimal schedule for hyperthermia administration during definitive radiotherapy can depend on several factors, including the specific protocol being followed, the stage and location of the tumor, and the patient's overall health status. However, based on general principles and clinical studies, hyperthermia is often administered once or twice weekly. The rationale behind administering hyperthermia once or twice weekly is to maximize its radiosensitizing effect while minimizing potential side effects and ensuring that the treatment can be tolerated by the patient. Hyperthermia works by heating the tumor to a higher temperature than the surrounding normal tissues, which can increase the effectiveness of radiation therapy. This is because heated cells are more sensitive to radiation damage. Administering hyperthermia twice weekly may offer a more intense radiosensitizing effect, potentially leading to better tumor control, as it increases the frequency of heat treatment in relation to radiation fractions. However, this approach must be balanced against the potential for increased toxicity and the practical considerations of treatment delivery, including patient tolerance and the logistics of hyperthermia treatment sessions. On the other hand, administering hyperthermia once weekly may be more feasible and better tolerated by some patients, especially if they have significant comorbidities or if the hyperthermia treatment itself causes significant side effects. The once-weekly schedule can still provide a significant radiosensitizing effect, although it might be less intense compared to a twice-weekly schedule. The decision on whether to administer hyperthermia once or twice weekly should ideally be made based on clinical guidelines, the specific characteristics of the patient's tumor, and the patient's ability to tolerate the treatment. It's also important to consider the results of clinical trials and studies that have investigated the efficacy and safety of different hyperthermia schedules in the context of radiotherapy for cervical carcinoma. In summary, while both once and twice weekly hyperthermia administration have been used and studied, the choice between these schedules should be individualized and based on the best available evidence, patient factors, and clinical judgment. | In the context of treating locally advanced cervical carcinoma with hyperthermia as a radiosensitizer, it is generally recommended to administer hyperthermia twice weekly during definitive radiotherapy. This approach is based on evidence suggesting that twice-weekly hyperthermia can enhance the effectiveness of radiotherapy by improving tumor oxygenation and increasing the sensitivity of cancer cells to radiation. However, the specific treatment plan should be tailored to the individual patient's condition and response to therapy, and it is important to follow the protocols and guidelines established by the treating institution or the recommendations of the oncology team. |
| 9 | What additional thermal equivalent dose (in Gy) can be achieved when hyperthermia is combined with radiotherapy? | | |
|  | DeepSeek-V3 | Llama-3.3-70B-Instruct | GPT-4o |
|  | When hyperthermia is combined with radiotherapy, the thermal equivalent dose (TED) enhancement is typically estimated to be **0.5-1.0 Gy per °C** above normal tissue temperature (37°C) for temperatures in the range of **40-45°C**.  ### Key Points: 1. **Thermal Enhancement Ratio (TER):**   - Hyperthermia increases the radiosensitivity of tumor cells, often quantified by the **Thermal Enhancement Ratio (TER)**, which ranges from **1.2 to 1.6** (meaning radiation effectiveness increases by 20-60% when combined with hyperthermia).   - For example, at **41-43°C**, the TER is ~1.5, meaning a radiation dose of 2 Gy with hyperthermia may be equivalent to **3 Gy** without hyperthermia. 2. **Equivalent Dose Calculation:**   - If hyperthermia is applied at **42-43°C**, the additional thermal equivalent dose is roughly **0.5-1.0 Gy per °C** above 37°C.   - For instance, heating to **42°C** (5°C above baseline) could provide an additional **2.5-5 Gy** equivalent effect. 3. **Clinical Observations:**   - Studies suggest that hyperthermia can enhance radiation effects by **10-30%**, effectively allowing dose reductions while maintaining tumor control.   - Some protocols estimate that **60 minutes at 42.5°C** can enhance radiation effects similarly to an additional **1-2 Gy**. ### Conclusion: The exact additional thermal equivalent dose depends on **temperature, duration of heating, and tumor type**, but a reasonable estimate is **0.5-1.0 Gy per °C** in the therapeutic range (40-45°C). This synergy allows for improved tumor control while potentially sparing normal tissues. | When hyperthermia is combined with radiotherapy, it can enhance the effectiveness of radiation treatment by increasing the thermal equivalent dose. The thermal equivalent dose is a measure of the biological effect of heat on tissues, and it's typically expressed in terms of the equivalent dose of radiation that would produce the same biological effect. The additional thermal equivalent dose that can be achieved when hyperthermia is combined with radiotherapy depends on several factors, including: 1. Temperature: The temperature achieved during hyperthermia treatment is a critical factor. Higher temperatures can lead to greater thermal equivalent doses. 2. Duration: The duration of hyperthermia treatment also affects the thermal equivalent dose. Longer treatment times can result in higher thermal equivalent doses. 3. Tissue type: Different tissues have varying sensitivities to heat and radiation. The thermal equivalent dose can vary depending on the type of tissue being treated. 4. Radiation dose: The dose of radiation used in conjunction with hyperthermia can also impact the thermal equivalent dose.  Studies have shown that hyperthermia can increase the thermal equivalent dose by 1-5 Gy or more, depending on the specific treatment conditions. For example: * A study published in the International Journal of Radiation Oncology, Biology, Physics found that hyperthermia increased the thermal equivalent dose by 2-3 Gy in tumors treated with radiation doses of 60-70 Gy. * Another study published in the Journal of Clinical Oncology reported that hyperthermia increased the thermal equivalent dose by 4-5 Gy in patients with locally advanced cervical cancer treated with radiation doses of 45-50 Gy. It's essential to note that these values are approximate and can vary depending on the specific treatment protocol and patient population. Additionally, the thermal equivalent dose is not a direct measure of the biological effect of hyperthermia, but rather a way to estimate the potential enhancement of radiation treatment efficacy. In summary, the additional thermal equivalent dose that can be achieved when hyperthermia is combined with radiotherapy can range from 1-5 Gy or more, depending on the treatment conditions and tissue type. However, the actual biological effect of hyperthermia can vary and should be carefully evaluated in the context of individual patient treatment plans. | The additional thermal equivalent dose achieved when hyperthermia is combined with radiotherapy can vary depending on several factors, including the type of cancer, the specific protocol used, and individual patient characteristics. However, research has shown that hyperthermia can enhance the effects of radiotherapy, potentially allowing for a reduction in the required radiation dose or an increase in the therapeutic effect. In general, hyperthermia can increase the effectiveness of radiotherapy by making cancer cells more sensitive to radiation. This is often quantified using the concept of thermal enhancement ratio (TER), which is the ratio of the radiation dose required to achieve a certain biological effect without hyperthermia to the dose required with hyperthermia. Typical TER values range from 1.2 to 2.0, meaning that hyperthermia can effectively increase the radiation dose by 20% to 100% in terms of biological effect. However, it is important to note that the actual additional thermal equivalent dose in Gy is not a fixed value and should be determined based on clinical trials and specific treatment protocols. Oncologists and medical physicists will tailor the treatment plan to the individual patient, considering factors such as tumor type, location, and patient health. |
| 10 | A patient with cT2 cN0 cM0, G3 muscle-invasive bladder cancer (MIBC) has undergone prior TUR-BT and is receiving combined radio-chemotherapy with concurrent hyperthermia. Should hyperthermia be applied once or twice per week during definitive radiotherapy? | | |
|  | DeepSeek-V3 | Llama-3.3-70B-Instruct | GPT-4o |
|  | For a patient with **cT2 cN0 cM0, G3 muscle-invasive bladder cancer (MIBC)** undergoing **definitive chemoradiotherapy (CRT) with concurrent hyperthermia**, the optimal frequency of hyperthermia sessions depends on clinical evidence and practical considerations.  ### **Current Evidence on Hyperthermia in Bladder Cancer:** 1. **Hyperthermia (HT) enhances radiotherapy (RT) and chemotherapy effects** by improving oxygenation, increasing drug uptake, and inhibiting DNA repair.  2. **Most clinical trials** (e.g., Dutch Hyperthermia Group studies) have used **twice-weekly hyperthermia** (e.g., 1-4 hours before RT) with good tolerability and efficacy.  3. **Key studies (e.g., van der Zee et al., 2000)** showed improved local control and survival with combined RT + HT vs. RT alone in bladder cancer.  ### **Recommended Hyperthermia Schedule:** - **Twice-weekly hyperthermia** (e.g., on non-consecutive days) is **preferred** based on clinical trial protocols.  - **Once-weekly HT** may be considered if logistics or patient tolerance is a concern, but efficacy may be slightly reduced.  ### **Practical Considerations:** - **HT should be administered within 1-4 hours before RT** for optimal radiosensitization.  - **Monitor toxicity** (skin reactions, bladder discomfort) since HT can enhance acute side effects.  ## **Conclusion:**  **Twice-weekly hyperthermia** is the **preferred schedule** during definitive CRT for MIBC, supported by clinical evidence. However, institutional protocols and patient-specific factors should guide final decisions.  Would you like references to specific studies or further details on hyperthermia techniques? | For a patient with cT2 cN0 cM0, G3 muscle-invasive bladder cancer (MIBC) undergoing combined radio-chemotherapy with concurrent hyperthermia, the application frequency of hyperthermia is an important consideration. Hyperthermia, in this context, refers to the application of heat to the tumor area to enhance the effectiveness of radiation therapy and chemotherapy. The optimal frequency of hyperthermia application during definitive radiotherapy can depend on several factors, including the specific protocol being followed, the equipment available, the patient's tolerance to the treatment, and the clinical evidence supporting different schedules. However, based on clinical trials and guidelines, hyperthermia is often applied twice a week during radiotherapy for various types of cancer, including bladder cancer. Applying hyperthermia twice a week can potentially offer several advantages: 1. **Enhanced Tumor Sensitivity**: Hyperthermia can increase the sensitivity of tumor cells to radiation, potentially leading to better local control rates. 2. **Synergistic Effect with Chemotherapy**: When combined with chemotherapy, hyperthermia may enhance the delivery and efficacy of chemotherapeutic agents within the tumor. 3. **Improved Outcomes**: Some studies suggest that twice-weekly hyperthermia, in conjunction with radiotherapy and chemotherapy, may lead to improved local control and possibly survival rates in certain cancers. However, the decision to apply hyperthermia once or twice a week should be individualized and based on the patient's specific condition, the treating institution's protocols, and the availability of resources. It's also crucial to monitor the patient for any adverse effects and adjust the treatment plan as necessary. In summary, for a patient with muscle-invasive bladder cancer receiving combined radio-chemotherapy, applying hyperthermia twice a week could be considered a reasonable approach, given the potential for enhanced efficacy. Nonetheless, the final decision should be made by the multidisciplinary team managing the patient's care, taking into account the latest clinical evidence and guidelines. | In the context of muscle-invasive bladder cancer (MIBC) being treated with combined radio-chemotherapy and concurrent hyperthermia, the application of hyperthermia is typically done once or twice per week. However, the optimal frequency can depend on institutional protocols and specific patient factors. Generally, hyperthermia is often applied once a week during definitive radiotherapy, but some protocols may use it twice a week to potentially enhance the therapeutic effect. It is important to follow the specific treatment protocol recommended by the oncology team, as they will tailor the approach based on the latest evidence and the patient's individual circumstances. |
| 11 | What are the typical Grade III acute toxicities associated with deep regional hyperthermia in combination with radiotherapy, and how high is the risk? | | |
|  | DeepSeek-V3 | Llama-3.3-70B-Instruct | GPT-4o |
|  | Deep regional hyperthermia (D-RHT) combined with radiotherapy (RT) is used to enhance tumor response, particularly in pelvic and abdominal malignancies. While generally well-tolerated, **Grade III acute toxicities** (severe but not life-threatening, requiring intervention) can occur. Here are the most common types and their associated risks: ### **Typical Grade III Acute Toxicities**  1. **Skin Burns/Blistering**   - Due to localized overheating at the treatment site.   - Risk: **~5-10%** (higher in areas with poor thermoregulation or prior RT).  2. **Pain at Treatment Site**   - Often musculoskeletal or visceral (e.g., pelvic pain).   - Risk: **~5-15%**, depending on tumor location.  3. **Gastrointestinal Toxicity**   - Nausea, vomiting, or diarrhea (especially in abdominal/pelvic treatments).   - Risk: **~5-10%** (higher with concurrent chemotherapy).  4. **Fatigue**   - Exacerbated by combined RT and hyperthermia.   - Risk: **~10-20%** (usually Grade II, but severe cases occur).  5. **Thermal Burns in Normal Tissues**   - Rare but possible if thermometry fails or hotspots develop.   - Risk: **<5%** with modern systems.  6. **Cardiovascular Stress**   - Tachycardia or blood pressure fluctuations due to systemic heating.   - Risk: **Low (<3%)** in properly selected patients.  ### **Risk Factors Influencing Toxicity**  - **Treatment Site**: Pelvic/abdominal regions have higher risks than extremities.  - **Prior RT**: Increases skin and tissue sensitivity.  - **Combined Therapies**: Concurrent chemo (e.g., cisplatin) raises GI and hematologic risks.  - **Patient Factors**: Obesity, poor skin integrity, or vascular diseases may increase complications.  ### **Mitigation Strategies**  - **Strict thermometry monitoring** to avoid overheating.  - **Individualized power adjustment** based on patient feedback.  - **Prophylactic antiemetics** for GI toxicity.  - **Cooling techniques** for sensitive skin areas.  ### **Conclusion**  Grade III toxicities are **uncommon (5-15% overall)** and manageable with proper protocols. The benefits of enhanced tumor control often outweigh risks, particularly in radioresistant cancers.  Would you like data from specific clinical trials? | Deep regional hyperthermia (DRHT) in combination with radiotherapy is a treatment approach used to enhance the effectiveness of radiation therapy in cancer treatment. While it can be beneficial, it also comes with potential side effects, known as acute toxicities. Grade III acute toxicities are considered severe and can significantly impact a patient's quality of life and treatment course. The typical Grade III acute toxicities associated with DRHT in combination with radiotherapy can vary depending on the treated area and individual patient factors. However, common toxicities may include: 1. **Skin Toxicity**: Severe skin reactions such as moist desquamation, where the skin peels off, or even ulceration can occur. These reactions are painful and can lead to infection if not properly managed. 2. **Gastrointestinal Toxicity**: For treatments targeting abdominal or pelvic areas, severe diarrhea, nausea, and vomiting can be significant issues. These symptoms can lead to dehydration and electrolyte imbalances if not adequately controlled. 3. **Urinary Toxicity**: Treatments involving the pelvic area can cause severe urinary frequency, urgency, or pain during urination. In some cases, there might be a decrease in bladder capacity. 4. **Fatigue**: While fatigue is a common side effect of both radiation and hyperthermia, Grade III fatigue is severe and can significantly interfere with daily activities. 5. **Pain**: Severe pain at the treatment site or in the surrounding areas can occur, requiring strong pain management strategies. The risk of developing these Grade III acute toxicities can vary widely among patients. Factors influencing this risk include the specific location and size of the tumor, the dose and schedule of radiation therapy, the temperature achieved during hyperthermia, the number of hyperthermia sessions, and individual patient characteristics such as overall health, nutritional status, and the presence of comorbid conditions. Generally, the incidence of Grade III or higher acute toxicities in patients undergoing DRHT combined with radiotherapy can range from about 10% to 30% or more, depending on the specific study and the factors mentioned above. It's crucial for patients to be closely monitored by their healthcare team during and after treatment to promptly address any emerging toxicities and to adjust the treatment plan as necessary to minimize these risks. Each patient's experience with DRHT and radiotherapy can be unique, and the risk of acute toxicities should be discussed in detail with a healthcare provider to understand the potential benefits and risks of this treatment approach for their specific situation. | Deep regional hyperthermia combined with radiotherapy is a treatment approach used to enhance the effectiveness of radiotherapy, particularly in certain types of cancer. While this combination can be beneficial, it also carries a risk of acute toxicities. Grade III acute toxicities are considered severe and can significantly impact a patient's quality of life and treatment course. Typical Grade III acute toxicities associated with this treatment combination may include: 1. **Skin Reactions**: Severe erythema, blistering, or ulceration in the area where hyperthermia is applied. 2. **Pain**: Intense pain in the treated region, which may require medication for management. 3. **Gastrointestinal Issues**: Nausea, vomiting, or diarrhea, especially if the abdominal or pelvic regions are treated. 4. **Fatigue**: Severe fatigue that affects daily activities. 5. **Hematological Effects**: Although less common, there can be significant drops in blood cell counts, leading to anemia, leukopenia, or thrombocytopenia. The risk of these Grade III toxicities varies depending on several factors, including the specific cancer type, the location of treatment, the patient's overall health, and the precise parameters of the hyperthermia and radiotherapy protocols used. Generally, the incidence of severe toxicities is relatively low, but it is crucial for healthcare providers to monitor patients closely and manage any adverse effects promptly. Patients undergoing this treatment should be informed about the potential risks and signs of severe toxicities, and they should maintain open communication with their healthcare team to address any symptoms that arise during the course of treatment. |
| 12 | What are contraindications for superficial hyperthermia? | | |
|  | DeepSeek-V3 | Llama-3.3-70B-Instruct | GPT-4o |
|  | Superficial hyperthermia, a therapeutic technique that heats tissues to 40-45°C to enhance radiation or chemotherapy effects, has several contraindications. These include: ### **Absolute Contraindications** (should not be used): 1. **Metallic Implants or Pacemakers** - Risk of overheating or interference with device function. 2. **Pregnancy** - Potential harm to the fetus due to thermal effects. 3. **Severe Cardiovascular Disease** - Risk of exacerbating heart conditions due to increased blood flow and thermal stress. 4. **Bleeding Disorders or Active Hemorrhage** - Heat may worsen bleeding. 5. **Overlying Skin Infections or Open Wounds** - Risk of spreading infection or delayed healing. 6. **Areas with Severe Peripheral Vascular Disease** - Poor circulation may lead to thermal injury. 7. **Tumors Near Critical Structures** (e.g., eyes, brain, spinal cord) - Risk of thermal damage to sensitive tissues. 8. **Previous Radiation Fibrosis** - Increased risk of tissue necrosis. ### **Relative Contraindications** (use with caution): 1. **Diabetes Mellitus with Neuropathy** - Reduced thermal sensitivity increases burn risk. 2. **Previous Thermal Injury or Burns in the Area** - Higher susceptibility to damage. 3. **Large or Deep Tumors** - Superficial heating may not penetrate effectively. 4. **Patients with Impaired Thermoregulation** (e.g., elderly, neurological disorders). 5. **Active Skin Conditions** (e.g., eczema, psoriasis) - Heat may worsen inflammation. 6. **Use of Photosensitizing Drugs** - Increased risk of adverse reactions. ### **Precautions:** - **Monitor temperature closely** to avoid burns. - **Avoid excessive heating** over bony prominences or scar tissue. - **Assess patient tolerance** during treatment. Always consult clinical guidelines and evaluate individual patient risks before proceeding. Would you like details on specific conditions? | Superficial hyperthermia is a treatment that uses heat to target and destroy cancer cells or other abnormal tissues near the surface of the body. While it can be an effective treatment for certain conditions, there are several contraindications where superficial hyperthermia should be avoided or used with caution. These include: 1. **Pregnancy**: The safety of hyperthermia during pregnancy has not been well established, and there is a potential risk of harm to the fetus. Therefore, it is generally contraindicated in pregnant women. 2. **Metal Implants**: The presence of metal implants (such as hip replacements, pacemakers, or metal fragments) can be a contraindication because the metal can heat up during the treatment, potentially causing burns or other injuries. 3. **Pacemakers and Other Implantable Electronic Devices**: The electromagnetic fields generated by hyperthermia equipment can interfere with the functioning of pacemakers and other implantable electronic devices, which could lead to life-threatening arrhythmias or other complications. 4. **Tattoos or Permanent Makeup**: Areas with tattoos or permanent makeup should be avoided because the pigments can react to the heat, leading to burns or discoloration. 5. **Open Wounds or Ulcers**: Applying heat to open wounds or ulcers can interfere with the healing process and increase the risk of infection. 6. **Sensitive Areas**: Areas with sensitive tissues, such as the eyes, testes, or certain parts of the brain, should be avoided due to the potential for damage from excessive heat. 7. **Previous Radiation Therapy**: Tissues that have been previously exposed to radiation therapy may have altered blood supply and be more susceptible to damage from heat, making them a relative contraindication for superficial hyperthermia. 8. **Certain Medical Conditions**: Conditions such as poor circulation, diabetes (which can lead to neuropathy and decreased sensation, making it harder to detect burns), and connective tissue diseases may increase the risk of complications from hyperthermia. 9. **Implantable Drug Delivery Systems**: Similar to pacemakers, the functioning of implantable drug delivery systems could potentially be affected by the heat or electromagnetic fields generated during hyperthermia treatment. 10. **Claustrophobia or Anxiety**: The treatment setup for superficial hyperthermia can sometimes cause feelings of claustrophobia or anxiety in patients, which may need to be addressed before proceeding with treatment. It's crucial for patients to discuss their medical history and any concerns with their healthcare provider before undergoing superficial hyperthermia to determine if the treatment is safe and appropriate for their specific situation. | Superficial hyperthermia is a treatment modality often used in conjunction with other therapies, such as radiation or chemotherapy, to treat certain types of cancer. It involves raising the temperature of the tumor tissue to enhance the effects of these treatments. However, there are several contraindications and considerations to keep in mind: 1. **Implanted Medical Devices**: Patients with pacemakers or other electronic implants may be at risk, as the heat and electromagnetic fields could interfere with the device's function. 2. **Metal Implants**: Metal in the treatment area can conduct heat and cause burns or other damage to surrounding tissues. 3. **Severe Cardiovascular Conditions**: Patients with significant heart disease may not tolerate the increased stress on the cardiovascular system. 4. **Infections or Open Wounds**: Hyperthermia can exacerbate infections or interfere with wound healing. 5. **Pregnancy**: The effects of hyperthermia on a developing fetus are not well understood, so it is generally avoided during pregnancy. 6. **Severe Respiratory Conditions**: Patients with compromised lung function may not tolerate the treatment well. 7. **Neurological Disorders**: Conditions that affect temperature regulation or sensation may increase the risk of adverse effects. 8. **Poor General Health**: Patients who are extremely frail or have poor performance status may not be suitable candidates. 9. **Sensitivity to Heat**: Some individuals may have a heightened sensitivity to heat, which could lead to discomfort or adverse reactions. 10. **Tumor Location**: Tumors located near critical structures that could be damaged by heat may not be suitable for this treatment. It is essential for healthcare providers to evaluate each patient's overall health, medical history, and specific circumstances before proceeding with superficial hyperthermia. |
| 13 | What are contraindications for deep regional hyperthermia? | | |
|  | DeepSeek-V3 | Llama-3.3-70B-Instruct | GPT-4o |
|  | Deep regional hyperthermia (DRH) is a therapeutic technique that raises tissue temperature to 40-45°C to enhance the effects of radiation or chemotherapy, primarily in cancer treatment. However, it has several **contraindications** due to potential risks. These include: ### **Absolute Contraindications (Not Safe Under Any Circumstances)** 1. **Metallic Implants or Pacemakers** - Risk of overheating or malfunction due to electromagnetic fields. 2. **Pregnancy** - Potential harm to the fetus from elevated temperatures. 3. **Severe Cardiovascular Disease** - Unstable angina, recent myocardial infarction, or severe heart failure (hyperthermia increases cardiac workload). 4. **Severe Cerebrovascular Disease** - Recent stroke or intracranial hemorrhage (risk of increased intracranial pressure). 5. **Active Bleeding or Hemorrhagic Disorders** - Hyperthermia can exacerbate bleeding. 6. **Severe Peripheral Vascular Disease** - Risk of thrombosis or tissue damage. 7. **Large Tumors with Necrosis or Cysts** - Risk of overheating and rupture. 8. **Severe Infection or Sepsis** - Hyperthermia may worsen systemic infection. ### **Relative Contraindications (Caution Required, Risk-Benefit Assessment Needed)** 1. **Previous Radiation Therapy in the Same Area** - Increased risk of tissue damage. 2. **Chronic Kidney or Liver Disease** - Impaired thermoregulation and drug metabolism. 3. **Diabetes with Neuropathy or Vascular Complications** - Risk of poor heat dissipation. 4. **Epilepsy or Seizure Disorders** - Hyperthermia may lower seizure threshold. 5. **Extensive Surgical Scars or Skin Grafts** - Risk of poor heat conduction and burns. 6. **Compromised Thermoregulation (e.g., elderly, very young patients)** - Higher risk of overheating. 7. **Tumors Near Critical Structures (e.g., spinal cord, eyes, brain)** - Risk of thermal damage. ### **Additional Considerations** - **Concurrent Use of Certain Chemotherapies** (e.g., cisplatin, bleomycin) may increase toxicity. - **Patients with Poor General Health (Karnofsky Performance Status < 50%)** may not tolerate treatment well. A thorough **medical evaluation** is essential before DRH to ensure patient safety. Would you like details on specific conditions? | Deep regional hyperthermia is a type of cancer treatment that uses heat to kill cancer cells. While it can be an effective treatment for certain types of cancer, there are some contraindications to consider. Here are some of the contraindications for deep regional hyperthermia: 1. **Pregnancy**: Hyperthermia can cause fetal harm, so it is contraindicated in pregnant women. 2. **Metal implants**: Patients with metal implants, such as pacemakers, artificial joints, or metal fragments, may not be suitable for deep regional hyperthermia, as the heat can cause the metal to malfunction or heat up excessively. 3. **Cardiovascular disease**: Patients with severe cardiovascular disease, such as unstable angina, recent myocardial infarction, or congestive heart failure, may not be able to tolerate the increased cardiac workload caused by hyperthermia. 4. **Poor circulation**: Patients with poor circulation, such as those with peripheral artery disease, may not be able to dissipate heat effectively, which can lead to burns or other complications. 5. **Sickle cell disease**: Patients with sickle cell disease may be at risk of sickling crises due to the heat, which can cause red blood cells to become misshapen and obstruct blood vessels. 6. **Neurological disorders**: Patients with certain neurological disorders, such as multiple sclerosis or peripheral neuropathy, may be at risk of exacerbating their condition due to the heat. 7. **Recent surgery or trauma**: Patients who have recently undergone surgery or experienced trauma may not be suitable for deep regional hyperthermia, as the heat can interfere with wound healing or exacerbate existing injuries. 8. **Tumors in sensitive areas**: Tumors located in sensitive areas, such as the brain, spinal cord, or eyes, may not be suitable for deep regional hyperthermia, as the heat can cause damage to surrounding tissues. 9. **Implantable devices**: Patients with implantable devices, such as insulin pumps or neurostimulators, may need to have their devices removed or reprogrammed before undergoing deep regional hyperthermia. 10. **Severe claustrophobia**: Patients with severe claustrophobia may not be able to tolerate the enclosed space of the hyperthermia machine. It's essential to note that these contraindications may vary depending on the specific type of deep regional hyperthermia being used and the individual patient's condition. A thorough medical evaluation and consultation with a qualified healthcare professional are necessary to determine whether deep regional hyperthermia is a suitable treatment option for a particular patient. | Deep regional hyperthermia is a treatment method used to increase the temperature of tumor tissues, often in conjunction with other cancer treatments like radiation or chemotherapy. However, there are several contraindications and considerations to keep in mind when using this therapy: 1. **Implanted Medical Devices**: Patients with certain types of implanted medical devices, such as pacemakers or defibrillators, may not be suitable candidates for deep regional hyperthermia due to the risk of interference or damage to the device. 2. **Metal Implants**: The presence of metal implants in or near the treatment area can cause uneven heating and increase the risk of burns or tissue damage. 3. **Severe Cardiovascular Conditions**: Patients with severe heart conditions may not tolerate the increased stress on the cardiovascular system caused by hyperthermia. 4. **Severe Pulmonary Conditions**: Conditions that affect breathing or lung function can be exacerbated by the treatment. 5. **Infections or Open Wounds**: Hyperthermia can worsen infections or interfere with the healing of open wounds. 6. **Pregnancy**: The effects of hyperthermia on a developing fetus are not well understood, so it is generally avoided during pregnancy. 7. **Severe Cachexia**: Patients with severe wasting or malnutrition may not tolerate the treatment well. 8. **Neurological Disorders**: Certain neurological conditions may be exacerbated by the treatment. 9. **Uncontrolled Pain**: Patients experiencing severe, uncontrolled pain may not be able to tolerate the procedure. 10. **Tumor Location**: Tumors located near critical structures that could be damaged by heat may not be suitable for this treatment. It is essential for healthcare providers to evaluate each patient's overall health, medical history, and specific circumstances before proceeding with deep regional hyperthermia. |
| 14 | Is it feasible to adequately heat a 5 cm osteolytic bone metastasis in the sacrum bone”fully encased in intact cortical bone”using deep regional hyperthermia? | | |
|  | DeepSeek-V3 | Llama-3.3-70B-Instruct | GPT-4o |
|  | Heating a 5 cm osteolytic bone metastasis in the sacrum using **deep regional hyperthermia (RF or microwave-based)** is **challenging but potentially feasible**, depending on several factors. Here is a detailed breakdown: ### **1. Technical Feasibility** - **Penetration Depth**: Deep regional hyperthermia (e.g., BSD-2000, Celsius TCS) can heat tissues at depths of **8-12 cm**, but the sacrum's **dense cortical bone** and surrounding soft tissues may **attenuate energy deposition**. - **Target Localization**: The metastasis is encased in intact cortical bone, which acts as a **thermal insulator**, making it harder to achieve therapeutic temperatures (40-44°C) inside the lesion. - **Energy Absorption**: Bone has **low electrical conductivity** compared to soft tissue, so RF/microwaves may preferentially heat surrounding muscle and fat rather than the lesion itself. ### **2. Clinical Considerations** - **Tumor Location**: The sacrum is deep and near critical structures (nerves, bowel, bladder), increasing the risk of **off-target heating**. - **Osteolytic Nature**: Lytic lesions may have **reduced blood flow**, which can limit heat dissipation and improve heating efficiency (if energy reaches the target). - **Adjuvant Therapy**: Hyperthermia is often combined with **radiation or chemotherapy** to enhance efficacy, potentially allowing lower thermal doses. ### **3. Alternative Heating Methods** - **Focused Ultrasound (MR-HIFU)**: More precise for bone lesions but may struggle with cortical bone interference - **Interstitial Hyperthermia**: Requires invasive placement of antennas directly into the tumor (not ideal if fully encased in bone). - **Microwave Ablation**: More aggressive, but may not be suitable for palliative hyperthermia. ### **4. Evidence & Clinical Use** - Some studies report **moderate success** with deep hyperthermia for pelvic tumors, but **specific data on sacral metastases is limited**. - A **pilot trial** would be needed to assess feasibility, possibly using **thermometry (MRI or probes)** to monitor temperatures. ### **Conclusion** While **not straightforward**, deep regional hyperthermia **could be attempted** with careful planning, but **success depends on**: - The specific hyperthermia system used. - Ability to focus energy on the sacrum. - Real-time temperature monitoring. - Combination with other therapies (e.g., radiation). If precise heating is critical, **MR-guided focused ultrasound (MR-HIFU)** or **interstitial techniques** might be more reliable, though more invasive. A multidisciplinary evaluation (radiation oncology, hyperthermia specialists) is recommended. | To address whether it's feasible to adequately heat a 5 cm osteolytic bone metastasis in the sacrum bone using deep regional hyperthermia, we need to consider several factors, including the principles of deep regional hyperthermia, the characteristics of the tumor and its location, and the technical capabilities of the hyperthermia equipment. Deep regional hyperthermia is a type of cancer treatment that involves applying heat to a specific area of the body. The goal is to raise the temperature of the tumor to a level that is lethal to cancer cells (typically between 40°C and 45°C) without causing significant damage to surrounding healthy tissues. This method can be particularly challenging when the tumor is located in bone, as bone has a high thermal conductivity and can act as a heat sink, potentially reducing the effectiveness of the treatment. The sacrum, being a part of the pelvis, presents a complex anatomical structure for applying hyperthermia due to its deep location and the presence of surrounding tissues and organs. An osteolytic lesion (a type of bone lesion that results from the destruction of bone tissue) within the sacrum, fully encased in intact cortical bone, poses additional challenges. The cortical bone surrounding the lesion can absorb and dissipate some of the heat applied, potentially reducing the temperature achieved within the tumor. However, advancements in deep regional hyperthermia technology, such as the use of phased array systems or other sophisticated applicators, have improved the ability to focus heat on deep-seated tumors. These systems can be designed to tailor the heat distribution to the specific anatomy and location of the tumor, potentially overcoming some of the challenges posed by the bone and surrounding tissues. The feasibility of adequately heating the tumor also depends on the specific characteristics of the osteolytic lesion, such as its size (in this case, 5 cm), shape, and the extent to which it is vascularized. Well-vascularized tumors may be more effectively heated due to the increased blood flow, which can help in dissipating heat and potentially reducing hot spots. In conclusion, while there are challenges associated with heating a 5 cm osteolytic bone metastasis in the sacrum using deep regional hyperthermia, it is not entirely infeasible. The success of the treatment would depend on various factors, including the sophistication of the hyperthermia equipment, the expertise of the treating team, the specific anatomy and physiology of the patient, and the characteristics of the tumor itself. Advanced hyperthermia technologies and careful treatment planning can potentially overcome some of the obstacles, making it possible to achieve therapeutic temperatures within the tumor. However, each case should be evaluated individually, considering all relevant factors to determine the likelihood of success and to tailor the treatment approach accordingly. | Heating a 5 cm osteolytic bone metastasis in the sacrum using deep regional hyperthermia presents significant challenges, primarily due to the location and the nature of the tumor. The sacrum is a deep-seated bone, and the presence of intact cortical bone encasing the metastasis adds an additional layer of complexity. Cortical bone has a high density and low thermal conductivity, which can impede the effective transfer of heat to the tumor site. Deep regional hyperthermia aims to raise the temperature of tumor tissues to enhance the effects of other treatments like radiation or chemotherapy. However, achieving adequate and uniform heating in deep-seated tumors, especially those surrounded by dense bone, can be difficult. The effectiveness of hyperthermia depends on several factors, including the depth of the tumor, the surrounding tissue characteristics, and the technology used. Advanced hyperthermia techniques, such as phased-array systems or magnetic resonance-guided focused ultrasound (MRgFUS), may offer some potential in targeting deep-seated tumors. These technologies can focus energy more precisely, potentially overcoming some of the challenges posed by the bone's density and depth. However, the feasibility and effectiveness would still depend on the specific clinical scenario, including the equipment available and the patient's overall condition. In practice, a multidisciplinary team would need to evaluate the specific case, considering the tumor's characteristics, the patient's anatomy, and the available technology, to determine if deep regional hyperthermia is a viable option. |
| 15 | A patient underwent an unplanned R1 resection of a G2 soft tissue sarcoma. Re-excision is not feasible, and adjuvant radiotherapy has been recommended by the tumor board. Should adjunctive deep hyperthermia be considered as a radiosensitizer? | | |
|  | DeepSeek-V3 | Llama-3.3-70B-Instruct | GPT-4o |
|  | The decision to consider adjunctive **deep hyperthermia (HT)** as a radiosensitizer in this case of an **R1-resected G2 soft tissue sarcoma (STS)** with no feasible re-excision and planned adjuvant radiotherapy (RT) should be based on the following considerations: ### **1. Evidence Supporting Hyperthermia in STS** - **Radiosensitization**: Hyperthermia (typically 40-44°C) enhances radiation effects by improving oxygenation, inhibiting DNA repair, and increasing tumor cell kill.  - **Clinical Data**:   - **Retrospective & Phase II Studies**: Some studies (e.g., Issels et al., *Lancet Oncol* 2010) suggest improved local control and survival when HT is combined with RT ± chemotherapy in high-risk STS.   - **Meta-Analyses**: A 2016 Cochrane review found HT + RT improved local control in soft tissue sarcomas compared to RT alone.   - **R1/R2 Resection Settings**: HT may be particularly beneficial in cases with positive margins (R1/R2) where residual microscopic disease is likely.  ### **2. Patient-Specific Factors** - **G2 (Intermediate-Grade) Sarcoma**: While not as aggressive as G3, an R1 resection increases recurrence risk.  - **No Feasible Re-excision**: Adjuvant RT is standard, but HT could further enhance local control.  - **Tumor Location & Feasibility of HT**: Deep hyperthermia (e.g., using regional or focused ultrasound-based systems) must be technically achievable for the tumor bed.  ### **3. Potential Risks & Limitations** - **Limited Availability**: HT is not widely accessible and requires specialized centers.  - **Toxicity**: Skin burns, pain, and rare systemic effects may occur.  - **Lack of Phase III Data**: Most evidence is from small studies; no large randomized trials confirm superiority over RT alone.  ### **4. Tumor Board Recommendation**  - If the tumor board recommended **adjuvant RT alone**, adding HT would be an **investigational but potentially beneficial** approach, particularly if the patient has high-risk features (e.g., close/positive margins, large tumor size, or deep location).  - If available, **HT + RT could be considered** after discussing risks/benefits with the patient.  ### **Conclusion**  Given the **R1 resection and inability to re-excise**, adjunctive deep hyperthermia **may be a reasonable option** to enhance radiosensitivity and improve local control, provided it is available and the patient is willing to accept its investigational nature. A multidisciplinary discussion (including radiation oncology and hyperthermia specialists) is advisable.  Would you like references to specific studies or guidelines? | In the case of an unplanned R1 resection of a G2 soft tissue sarcoma where re-excision is not feasible, and adjuvant radiotherapy has been recommended, the consideration of adjunctive deep hyperthermia as a radiosensitizer is a nuanced decision. Here's a breakdown of the factors involved: 1. **R1 Resection**: An R1 resection indicates that microscopic residual disease is present. This situation increases the risk of local recurrence, making adjuvant treatments crucial. 2. **G2 Soft Tissue Sarcoma**: The grade of the tumor (G2) suggests an intermediate-grade malignancy. The prognosis and treatment response can vary based on the grade, with higher grades generally having a worse prognosis and potentially benefiting more from aggressive adjuvant treatments. 3. **Adjuvant Radiotherapy**: Recommended by the tumor board, adjuvant radiotherapy is aimed at reducing the risk of local recurrence by targeting any remaining microscopic disease. Radiotherapy is a standard approach in such scenarios. 4. **Adjunctive Deep Hyperthermia**: Deep hyperthermia involves heating the tumor to a high temperature (usually between 40°C and 45°C) to enhance the effectiveness of radiotherapy. The rationale behind using hyperthermia as a radiosensitizer is that it can increase blood flow and oxygen delivery to the heated area, potentially making the tumor cells more sensitive to radiation. Additionally, hyperthermia can directly induce cell death and inhibit cellular repair mechanisms, further enhancing the effects of radiation. Considering these points, adjunctive deep hyperthermia could be considered as part of the treatment plan for several reasons: - **Enhanced Radiosensitivity**: Hyperthermia has been shown to increase the sensitivity of tumor cells to radiation, potentially leading to improved local control rates. - **Synergistic Effect**: The combination of hyperthermia and radiotherapy may have a synergistic effect, offering better outcomes than either treatment modality alone. - **Limited Additional Toxicity**: When properly administered, deep hyperthermia can have minimal additional toxicity compared to radiotherapy alone, although it does require specialized equipment and expertise. However, the decision to use adjunctive deep hyperthermia should be made on a case-by-case basis, considering factors such as the patient's overall health, the specific characteristics of the tumor, and the availability of hyperthermia facilities. It's also important to discuss the potential benefits and risks with the patient, as well as to consider any relevant clinical guidelines or evidence-based practices. In summary, given the scenario of an unplanned R1 resection of a G2 soft tissue sarcoma with adjuvant radiotherapy recommended and re-excision not feasible, adjunctive deep hyperthermia could be a valuable consideration as a radiosensitizer to potentially improve local control and outcomes. However, this decision should be individualized and based on a thorough discussion among the multidisciplinary tumor board. | In the context of an R1 resection of a G2 soft tissue sarcoma where re-excision is not feasible, adjuvant radiotherapy is a standard recommendation to manage residual disease and reduce the risk of local recurrence. The consideration of adjunctive deep hyperthermia as a radiosensitizer is a reasonable option in this scenario. Hyperthermia therapy involves raising the temperature of tumor tissues, which can enhance the effects of radiotherapy by increasing tumor oxygenation, disrupting DNA repair mechanisms, and sensitizing cancer cells to radiation. Clinical studies have shown that combining hyperthermia with radiotherapy can improve local control rates in certain types of soft tissue sarcomas. However, the decision to include hyperthermia should be based on several factors, including the availability of hyperthermia treatment at the facility, the specific characteristics of the tumor, the patient's overall health and preferences, and the expertise of the treating team. It is important to discuss the potential benefits and risks with the patient and consider multidisciplinary input from the oncology team. In summary, adjunctive deep hyperthermia could be considered as a radiosensitizer in this case, but it should be evaluated on an individual basis in consultation with the treating oncologists and the patient. |
| 16 | A patient presents with a 10 cm solitary plasmacytoma located in the iliac bone. Is hyperthermia a beneficial adjunct to definitive radiotherapy in this setting? | | |
|  | DeepSeek-V3 | Llama-3.3-70B-Instruct | GPT-4o |
|  | The role of **hyperthermia** as an adjunct to **definitive radiotherapy** in **solitary plasmacytoma** (particularly a 10 cm lesion in the iliac bone) is not well-established in current clinical practice. Here is a breakdown of the evidence and considerations: ### **1. Current Standard of Care for Solitary Plasmacytoma**  - **Definitive radiotherapy (RT)** is the primary treatment, with doses typically ranging from **40-50 Gy**, achieving high local control rates (~90% for solitary bone plasmacytoma).  - Large lesions (>5 cm) have a higher risk of progression to multiple myeloma, but RT remains the cornerstone. ### **2. Hyperthermia as an Adjunct to RT**  - **Mechanism**: Hyperthermia (heating tumor tissue to 40-45°C) can enhance radiotherapy by:  - Increasing tumor oxygenation (improving radiation sensitivity).  - Disrupting DNA repair mechanisms in cancer cells.  - **Evidence in Other Cancers**:   - Some benefit seen in **soft tissue sarcomas**, **cervical cancer**, and **recurrent breast cancer** when combined with RT.  - Limited data in **bone tumors** or **plasmacytoma**.  - **Challenges in Bone Lesions**:  - Heating deep-seated bony lesions (like the iliac bone) is technically difficult.  - No randomized trials support its use in plasmacytoma. ### **3. Potential Considerations for This Patient**  - **Tumor Size (10 cm)**: Larger lesions may have hypoxic regions that could theoretically benefit from hyperthermia, but clinical evidence is lacking.  - **Alternative Strategies**:  - **Higher RT doses** (e.g., up to 50 Gy) may be considered for large lesions.  - **Systemic therapy** (e.g., immunomodulatory drugs or proteasome inhibitors) if there is concern for early myeloma. ### **4. Conclusion**  - **Hyperthermia is not a standard or evidence-based adjunct for solitary plasmacytoma**, even in large lesions.  - **Best approach**: Optimize RT dose and consider systemic therapy if high-risk features (e.g., M-protein persistence, progression risk).  - **Future research**: Hyperthermia remains experimental in this setting; clinical trials would be needed to assess its role. Would you like references to specific studies on hyperthermia in bone tumors or plasmacytoma? | Hyperthermia, or the application of heat to a tumor, has been explored as an adjunct to radiation therapy for various types of cancer, including solitary plasmacytoma. The rationale behind combining hyperthermia with radiotherapy is based on the principle that heat can enhance the sensitivity of cancer cells to radiation, potentially leading to improved tumor control rates. In the context of a solitary plasmacytoma, which is a localized collection of plasma cells in the bone marrow, the effectiveness of hyperthermia as an adjunct to radiotherapy is not well-established. Solitary plasmacytomas are typically treated with definitive radiotherapy, and the role of hyperthermia in this setting is largely investigational. There are a few key points to consider: 1. **Radiosensitivity of Plasmacytoma**: Plasmacytomas are generally considered to be radiosensitive, meaning they respond well to radiation therapy. Definitive radiotherapy is often curative for solitary plasmacytomas, with high local control rates reported in the literature. 2. **Hyperthermia and Radiation**: Hyperthermia can increase the effectiveness of radiation by making cancer cells more sensitive to radiation damage. This is thought to occur through several mechanisms, including the inhibition of DNA repair, the induction of apoptosis (programmed cell death), and changes in the tumor microenvironment that make it more susceptible to radiation. 3. **Clinical Evidence**: The clinical evidence supporting the use of hyperthermia in conjunction with radiotherapy for solitary plasmacytoma is limited. Most studies on hyperthermia and radiation have focused on other types of cancer, such as sarcomas, melanomas, and certain types of carcinomas. There is a need for specific clinical trials to evaluate the efficacy and safety of hyperthermia as an adjunct to radiotherapy in patients with solitary plasmacytoma. 4. **Potential Risks and Challenges**: Hyperthermia can cause side effects, including pain, burns, and damage to surrounding normal tissues. The technical challenges of delivering hyperthermia, especially to deep-seated tumors like those in the iliac bone, can be significant. Ensuring uniform heating of the tumor while minimizing damage to adjacent tissues is a complex task. In summary, while hyperthermia has shown promise as an adjunct to radiotherapy in certain cancer types, its benefit in the treatment of solitary plasmacytoma is not well-established. Definitive radiotherapy remains the standard of care for solitary plasmacytomas, and the use of hyperthermia in this context would be considered experimental. Patients with solitary plasmacytoma should discuss their treatment options with their healthcare provider, considering the potential benefits and risks of all available therapies. | Hyperthermia can be a beneficial adjunct to radiotherapy in the treatment of certain cancers, including solitary plasmacytoma. Hyperthermia involves raising the temperature of tumor tissues, which can enhance the effects of radiotherapy by making cancer cells more sensitive to radiation. This is because hyperthermia can improve blood flow to the tumor, increase oxygenation, and interfere with DNA repair mechanisms in cancer cells, thereby enhancing the cytotoxic effects of radiation. In the case of a solitary plasmacytoma, which is a localized form of plasma cell neoplasm, the primary treatment is often radiotherapy. Adding hyperthermia could potentially improve local control of the disease by increasing the effectiveness of the radiation treatment. However, the use of hyperthermia as an adjunct therapy should be considered on a case-by-case basis, taking into account the specific characteristics of the tumor, the patient's overall health, and the availability of hyperthermia treatment facilities. It is important to consult with a multidisciplinary team, including oncologists and radiation specialists, to determine the best treatment approach for the individual patient. Clinical trials and studies may provide additional evidence on the efficacy of hyperthermia in combination with radiotherapy for solitary plasmacytoma, and these should be reviewed to make an informed decision. |
| 17 | For which tumor entities is hyperthermia recognized as a standard oncologic treatment modality in international clinical guidelines? | | |
|  | DeepSeek-V3 | Llama-3.3-70B-Instruct | GPT-4o |
|  | Hyperthermia is recognized as a **complementary** treatment modality in several tumor entities, but it is rarely considered a **standard standalone therapy** in most international clinical guidelines. However, it has been incorporated into some guidelines for specific cancers, particularly when combined with radiation therapy or chemotherapy. Here are the key tumor entities where hyperthermia has **some level of recognition** in international guidelines: ### **1. Locally Advanced or Recurrent Soft Tissue Sarcomas**   - **ESMO (European Society for Medical Oncology)** and **NCCN (National Comprehensive Cancer Network)** acknowledge hyperthermia (especially **regional hyperthermia combined with chemotherapy**) as an option for high-risk or recurrent soft tissue sarcomas.   - **EORTC (European Organisation for Research and Treatment of Cancer)** has supported its use in combination with chemotherapy (e.g., **HIPEC** for peritoneal sarcomatosis). ### **2. Cervical Cancer (Locally Advanced or Recurrent)**   - **ESTRO (European Society for Radiotherapy & Oncology)** and **ESMO** recognize **hyperthermia combined with radiation therapy** as an option for recurrent or locally advanced cervical cancer, based on randomized trials showing improved outcomes. ### **3. Breast Cancer (Chest Wall Recurrences or Inflammatory Breast Cancer)**   - Some guidelines (e.g., **German S3 guidelines**) support hyperthermia in combination with radiation for recurrent chest wall disease or inflammatory breast cancer. ### **4. Bladder Cancer (Non-Muscle Invasive, High-Risk or BCG-Unresponsive)**   - **EAU (European Association of Urology)** and **AUA (American Urological Association)** mention **hyperthermic intravesical chemotherapy (HIVEC)** as an investigational option for high-risk or BCG-unresponsive non-muscle-invasive bladder cancer (NMIBC) ### **5. Peritoneal Carcinomatosis (Colorectal, Ovarian, Gastric, Appendiceal Cancers)**   - **HIPEC (Hyperthermic Intraperitoneal Chemotherapy)** is recognized in **ESMO, NCCN, and Peritoneal Surface Oncology Group International (PSOGI)** guidelines for select cases of peritoneal metastases (e.g., colorectal, ovarian, appendiceal cancers). ### **6. Melanoma (In-Transit Metastases or Recurrent Limb Lesions)**   - **Isolated Limb Perfusion (ILP) or Isolated Limb Infusion (ILI) with hyperthermia** is mentioned in **NCCN and ESMO** guidelines for unresectable in-transit melanoma. ### **7. Glioblastoma (Recurrent or Newly Diagnosed, Investigational Use)**   - **TTFields (Tumor Treating Fields) with mild hyperthermia effects** is included in **NCCN and ESMO** guidelines for glioblastoma, though this is distinct from traditional hyperthermia. ### **Key Notes:**  - **Hyperthermia is most effective when combined with radiation or chemotherapy** (thermoradiotherapy or thermochemotherapy).  - **Strongest evidence exists for soft tissue sarcomas, cervical cancer, and peritoneal malignancies (HIPEC).**  - **Not universally accepted as a standard in all countries**, but more commonly used in Europe (Germany, Netherlands) than in the U.S.  For the most current recommendations, consulting **NCCN, ESMO, or ESTRO guidelines** for specific tumor types is advised. Would you like references for any particular guideline? | Hyperthermia, also known as thermal therapy or thermotherapy, is a type of cancer treatment that uses heat to damage or kill cancer cells. While it is not a widely recognized standard treatment modality for all types of cancer, it is recognized as a standard oncologic treatment modality in international clinical guidelines for certain tumor entities. Here are some examples: 1. **Superficial tumors**: Hyperthermia is recognized as a standard treatment modality for superficial tumors, such as: * Recurrent breast cancer (National Comprehensive Cancer Network (NCCN) guidelines) * Melanoma (European Society for Medical Oncology (ESMO) guidelines) * Squamous cell carcinoma of the skin (NCCN guidelines) 2. **Cervical cancer**: Hyperthermia is recognized as a standard treatment modality for cervical cancer, particularly for: * Advanced or recurrent cervical cancer (NCCN guidelines) * Cervical cancer with radiation therapy (International Journal of Hyperthermia guidelines) 3. **Bladder cancer**: Hyperthermia is recognized as a standard treatment modality for bladder cancer, particularly for: * Non-muscle-invasive bladder cancer (European Association of Urology (EAU) guidelines) * Recurrent bladder cancer (NCCN guidelines) 4. **Soft tissue sarcomas**: Hyperthermia is recognized as a standard treatment modality for soft tissue sarcomas, particularly for: * Recurrent or metastatic soft tissue sarcomas (NCCN guidelines) * Soft tissue sarcomas with radiation therapy (European Society for Medical Oncology (ESMO) guidelines) 5. **Brain tumors**: Hyperthermia is recognized as a standard treatment modality for certain brain tumors, particularly for:  * Recurrent glioblastoma (NCCN guidelines) * Brain metastases (International Journal of Hyperthermia guidelines) It's essential to note that hyperthermia is often used in combination with other cancer treatments, such as radiation therapy, chemotherapy, or surgery, and its effectiveness may vary depending on the specific tumor type, stage, and location. Additionally, not all hyperthermia techniques are recognized as standard treatments, and some may still be considered experimental or investigational International clinical guidelines, such as those from the NCCN, ESMO, and EAU, provide evidence-based recommendations for the use of hyperthermia in cancer treatment. However, the availability and acceptance of hyperthermia as a standard treatment modality may vary depending on the country, region, or institution. | Hyperthermia is recognized as a standard oncologic treatment modality in international clinical guidelines for certain tumor entities, often in combination with other treatments like radiotherapy or chemotherapy. Some of the tumor types where hyperthermia is considered a standard treatment option include: 1. **Soft Tissue Sarcomas**: Hyperthermia is often used in combination with chemotherapy for the treatment of soft tissue sarcomas, particularly in the limbs. 2. **Recurrent Breast Cancer**: Hyperthermia can be used in combination with radiotherapy for the treatment of recurrent breast cancer, especially in cases where the cancer is located in previously irradiated areas. 3. **Cervical Cancer**: In some guidelines, hyperthermia is used alongside radiotherapy for locally advanced cervical cancer. 4. **Head and Neck Cancers**: Hyperthermia may be used in combination with radiotherapy for certain head and neck cancers, particularly in recurrent cases. These applications are supported by clinical evidence showing that hyperthermia can enhance the effectiveness of other treatments by increasing tumor oxygenation and making cancer cells more susceptible to radiation and chemotherapy. However, the availability and use of hyperthermia can vary by region and institution, and it is not universally adopted in all clinical settings. Always refer to the specific guidelines of the region or institution for the most accurate and applicable information. |
| 18 | Which tumor type is most likely to derive significant benefit from the addition of hyperthermia? | | |
|  | DeepSeek-V3 | Llama-3.3-70B-Instruct | GPT-4o |
|  | The tumor type most likely to derive significant benefit from the addition of **hyperthermia** (HT) to standard treatments (such as radiation therapy or chemotherapy) is **locally advanced or recurrent soft tissue sarcoma**, particularly when combined with **radiation therapy**.  ### **Key Tumor Types with Strong Evidence for Hyperthermia Benefit:**  1. **Soft Tissue Sarcomas (STS)** -   - Hyperthermia combined with radiation improves local control and survival in high-risk or inoperable cases.   - Randomized trials (e.g., EORTC 62961) showed significant benefit in response rates and local progression-free survival.  2. **Locally Advanced or Recurrent Breast Cancer (Chest Wall Recurrences)** -   - Hyperthermia + radiation improves tumor response and local control in recurrent breast cancer after mastectomy.  3. **Cervical Cancer (Locally Advanced)** -   - Some studies suggest improved outcomes when hyperthermia is added to chemoradiation.  4. **Melanoma (Superficial or Nodal Metastases)** -   - Hyperthermia enhances radiation effects in radioresistant tumors like melanoma.  5. **Bladder Cancer (Non-Muscle Invasive or Locally Advanced)** -   - Hyperthermia with chemotherapy (e.g., mitomycin C) improves response in bladder cancer.  ### **Mechanism of Benefit:**  - Hyperthermia (typically 40-44°C) increases blood flow, enhances oxygen delivery, and sensitizes tumors to radiation and chemotherapy.  - It disrupts DNA repair mechanisms, making cancer cells more susceptible to treatment.  ### **Clinical Evidence:**  - **EORTC 62961 (STS):** Hyperthermia + radiation doubled complete response rates compared to radiation alone.  - **Duke University (Breast Cancer):** Hyperthermia improved local control in recurrent chest wall disease.  ### **Conclusion:**  **Soft tissue sarcomas** and **recurrent breast cancer** have the strongest evidence, but other locally advanced tumors (cervical, melanoma, bladder) may also benefit. Hyperthermia is most effective when combined with radiation or chemotherapy in well-selected cases.  Would you like details on specific clinical trials or protocols? | The tumor type that is most likely to derive significant benefit from the addition of hyperthermia is superficial or locally advanced tumors, particularly those that are resistant to radiation therapy or chemotherapy. Some examples of tumor types that may benefit from hyperthermia include: 1. **Melanoma**: Hyperthermia has been shown to improve local control and survival rates in patients with melanoma, particularly when combined with radiation therapy or chemotherapy. 2. **Soft tissue sarcomas**: Hyperthermia can enhance the effectiveness of radiation therapy and chemotherapy in treating soft tissue sarcomas, such as leiomyosarcoma and malignant fibrous histiocytoma. 3. **Breast cancer**: Hyperthermia has been used to treat recurrent or metastatic breast cancer, particularly in combination with radiation therapy or chemotherapy. 4. **Head and neck cancer**: Hyperthermia can improve local control and survival rates in patients with head and neck cancer, particularly when combined with radiation therapy or chemotherapy. 5. **Cervical cancer**: Hyperthermia has been shown to improve survival rates in patients with cervical cancer, particularly when combined with radiation therapy. In general, tumors that are most likely to benefit from hyperthermia are those that: * Are superficial or locally advanced * Are resistant to radiation therapy or chemotherapy * Have a high density of blood vessels (which can be targeted by hyperthermia) * Are located in areas where hyperthermia can be safely and effectively applied (e.g., superficial tumors) It's worth noting that hyperthermia is typically used in combination with other treatments, such as radiation therapy or chemotherapy, and its effectiveness can vary depending on the specific tumor type and treatment regimen. | Hyperthermia therapy is most likely to derive significant benefit when used in conjunction with other treatments for certain types of tumors, particularly soft tissue sarcomas, cervical cancer, and recurrent breast cancer. Hyperthermia can enhance the effectiveness of radiation therapy and certain chemotherapeutic agents, making it a valuable adjunct treatment for these cancers. Clinical studies have shown that hyperthermia can improve local control and overall outcomes in these tumor types. |
| 19 | In which tumor type do you perceive the greatest underutilized potential for hyperthermia? | | |
|  | DeepSeek-V3 | Llama-3.3-70B-Instruct | GPT-4o |
|  | Hyperthermia therapy has shown promise across several tumor types, but the **greatest underutilized potential** likely lies in **locally advanced or recurrent cervical cancer**, particularly when combined with **radiotherapy and/or chemotherapy**. Here is why: ### **1. Cervical Cancer: Strong Clinical Evidence**  - **Randomized trials** (e.g., Dutch Hyperthermia Group, Lancet 2000) demonstrated a **doubling of 3-year survival** (51% vs. 27%) when adding hyperthermia to radiotherapy for recurrent/persistent cervical cancer.  - Hyperthermia enhances radiation sensitivity in hypoxic, radioresistant tumors€”common in cervical cancer.  - Despite robust data, adoption remains limited due to logistical challenges (equipment, expertise). ### **2. Other High-Potential Candidates:**  - **Soft Tissue Sarcomas**: Hyperthermia + radiotherapy improves local control (e.g., EORTC 62961 trial).  - **Breast Cancer (chest wall recurrences)**: Significant response rates in palliative settings (e.g., 60% CR/PR with thermoradiotherapy).  - **Glioblastoma**: Emerging interest in localized hyperthermia (e.g., magnetic nanoparticles), but clinical data are still early-stage. ### **Barriers to Utilization**:  - **Technical complexity**: Requires precise temperature monitoring (41-43°C).  - **Limited access**: Few centers have hyperthermia-capable facilities.  - **Lack of awareness**: Often overlooked in guidelines despite Level I evidence for some indications. ### **Conclusion**: While hyperthermia could benefit many cancers, **cervical cancer** stands out due to strong efficacy data and a clear survival benefit that remains underused globally. Expanding access and integrating hyperthermia into multimodal protocols could significantly impact outcomes in this and other radio/chemo-sensitive tumors.  Would you like insights on specific hyperthermia techniques (e.g., RF, microwave, magnetic) for these tumors? | Hyperthermia, or the application of heat to treat cancer, has been explored in various tumor types with varying degrees of success and utilization. While it has shown promise in several cancers, there are certain tumor types where its potential might be particularly underutilized. One such tumor type is glioblastoma multiforme (GBM), a highly aggressive and malignant form of brain cancer. Glioblastoma multiforme is characterized by its rapid growth and resistance to current treatments, including surgery, radiation, and chemotherapy. The prognosis for patients with GBM remains poor, with a median survival time that has not significantly improved over the past few decades. This limited progress in treatment outcomes highlights the need for innovative and adjunctive therapies. Hyperthermia offers several potential benefits in the treatment of glioblastoma: 1. **Enhanced Drug Delivery**: Hyperthermia can increase blood flow and permeability in the tumor, potentially enhancing the delivery of chemotherapeutic agents to the tumor site. This could improve the efficacy of chemotherapy in treating GBM. 2. **Direct Cytotoxic Effects**: Heat can directly kill cancer cells or make them more susceptible to radiation and chemotherapy. In the context of GBM, where the tumor is highly resistant to conventional therapies, inducing hyperthermia could provide a novel method to target these cells. 3. **Immunomodulatory Effects**: Hyperthermia can stimulate an anti-tumor immune response. By applying heat to the tumor, it's possible to increase the expression of heat shock proteins on the surface of tumor cells, making them more recognizable and targetable by the immune system. 4. **Synergism with Other Therapies**: Hyperthermia can be used in conjunction with other treatments such as radiation therapy (thermoradiotherapy) and chemotherapy. This multimodal approach could lead to improved outcomes by targeting the tumor from multiple angles. Despite these potential benefits, the application of hyperthermia in glioblastoma is not as widespread as it could be. Several factors contribute to this underutilization, including: - **Technical Challenges**: Applying hyperthermia to brain tumors like GBM requires precise and controlled heating to avoid damaging the surrounding healthy brain tissue. Developing technologies that can safely and effectively deliver heat to deep-seated brain tumors is a significant challenge. - **Clinical Evidence**: While there is some evidence supporting the use of hyperthermia in various cancers, more clinical trials are needed to fully understand its efficacy and optimal application in GBM. - **Awareness and Adoption**: Among oncologists and neurosurgeons, there may be limited awareness or experience with hyperthermia as a treatment modality for GBM, which can hinder its adoption. In summary, glioblastoma multiforme represents a tumor type with significant unmet medical needs and potentially substantial underutilized potential for hyperthermia. Addressing the technical, clinical, and awareness challenges associated with hyperthermia in GBM could lead to improved treatment options for patients with this devastating disease. | Hyperthermia, the treatment that involves raising the temperature of tumor tissues, has shown potential in enhancing the effectiveness of other cancer treatments like radiation and chemotherapy. While it has been explored in various tumor types, one area where its potential might be underutilized is in the treatment of pancreatic cancer. Pancreatic cancer is notoriously difficult to treat due to its aggressive nature and the dense stromal tissue that surrounds the tumor, which can impede the delivery of therapeutic agents. Hyperthermia could potentially improve the efficacy of chemotherapy and radiation by increasing blood flow to the tumor, enhancing drug delivery, and sensitizing cancer cells to radiation. Additionally, hyperthermia might help in modulating the immune response, which could be beneficial given the immunosuppressive environment of pancreatic tumors. Despite these potential benefits, hyperthermia is not widely used in the treatment of pancreatic cancer, possibly due to technical challenges and the need for specialized equipment and expertise. Further research and clinical trials could help in understanding and harnessing the full potential of hyperthermia in this context. |
| 20 | Is combined hyperthermia and radiotherapy an appropriate strategy for the management of bulky tumors? | | |
|  | DeepSeek-V3 | Llama-3.3-70B-Instruct | GPT-4o |
|  | Combined **hyperthermia and radiotherapy (HT-RT)** is a promising strategy for the management of **bulky tumors**, particularly in cases where conventional treatments face limitations. Here is a detailed assessment of its appropriateness: ### **Rationale for Combining Hyperthermia with Radiotherapy** 1. **Radiosensitization**   - Hyperthermia (typically 40-44°C) enhances tumor sensitivity to radiation by:   - Increasing oxygen radical production (indirect DNA damage).   - Inhibiting DNA repair mechanisms (e.g., homologous recombination).   - Disrupting tumor vasculature, reducing hypoxia (a major cause of radioresistance).  2. **Direct Cytotoxic Effects**   - Heat alone can kill cancer cells, especially in hypoxic and acidic tumor microenvironments.  3. **Targeting Radioresistant Areas**   - Bulky tumors often contain hypoxic, necrotic, and poorly perfused regions that are resistant to radiation. Hyperthermia selectively heats these areas due to impaired thermoregulation in tumors.  ### **Clinical Evidence Supporting HT-RT for Bulky Tumors** - **Superficial Tumors (e.g., breast, melanoma, recurrent chest wall lesions):**   - Multiple randomized trials (e.g., Dutch Hyperthermia Group) show improved **local control and survival** when HT is added to RT.   - A meta-analysis (Issels et al., *Lancet Oncol.* 2010) demonstrated **doubled complete response rates** in soft tissue sarcomas.  - **Deep-Seated Tumors (e.g., cervical, prostate, glioblastoma):**   - Emerging evidence supports benefits, though heating technology (e.g., RF, microwave) remains a challenge.   - A phase III trial in cervical cancer (Franckena et al., *IJROBP* 2008) showed **improved survival** with HT-RT.  ### **Advantages for Bulky Tumors** - **Overcoming Hypoxia:** Heat improves oxygenation, making radiation more effective.  - **Synergistic Cytotoxicity:** Combined damage exceeds additive effects.  - **Potential for Dose Reduction:** May allow lower radiation doses, sparing normal tissues.  ### **Challenges & Limitations** - **Technical Difficulties:**   - Uniform heating of deep tumors is challenging (requires advanced systems like **MR-guided focused ultrasound**).  - **Toxicity Risks:**   - Overheating can damage normal tissues (e.g., burns, pain).  - **Limited Accessibility:**   - Hyperthermia is not widely available in all centers.  ### **Conclusion** Combined **hyperthermia and radiotherapy is an appropriate and evidence-supported strategy for bulky tumors**, particularly in:  - **Locally advanced or recurrent tumors** (e.g., breast, cervical, sarcoma).  - **Radioresistant hypoxic tumors** where conventional RT fails.  **Future Directions:**  - Refinement of **deep-heating technologies** (e.g., magnetic nanoparticle hyperthermia).  - Integration with **immunotherapy** (heat enhances immune responses).  Would you like specific protocol recommendations or references for particular tumor types? | Yes, combined hyperthermia and radiotherapy is a promising strategy for the management of bulky tumors. Here's why: **Rationale:** 1. **Enhanced radiosensitivity**: Hyperthermia (elevated temperature) can increase the sensitivity of tumor cells to radiation, making them more susceptible to damage and death. 2. **Increased blood flow**: Hyperthermia can increase blood flow to the tumor, which can enhance the delivery of oxygen and nutrients, making the tumor more responsive to radiation. 3. **Direct cytotoxic effects**: Hyperthermia can directly kill tumor cells or induce apoptosis (programmed cell death) by damaging cellular proteins, membranes, and DNA. **Benefits:** 1. **Improved local control**: Combined hyperthermia and radiotherapy can improve local control of bulky tumors, reducing the risk of local recurrence. 2. **Enhanced tumor response**: The combination of hyperthermia and radiotherapy can lead to a more pronounced tumor response, including increased tumor shrinkage and necrosis. 3. **Reduced toxicity**: Hyperthermia can allow for lower radiation doses, reducing the risk of toxicity to surrounding normal tissues. 4. **Increased survival**: Some studies have shown that combined hyperthermia and radiotherapy can improve overall survival rates in patients with bulky tumors. **Clinical applications:** 1. **Cervical cancer**: Combined hyperthermia and radiotherapy has been shown to improve local control and survival in patients with bulky cervical cancer. 2. **Head and neck cancer**: This combination has been used to treat bulky head and neck tumors, with promising results. 3. **Soft tissue sarcomas**: Hyperthermia and radiotherapy have been combined to treat soft tissue sarcomas, with improved local control and survival. 4. **Other tumors**: The combination is also being explored for other tumor types, including breast, lung, and brain cancers. **Challenges and limitations:** 1. **Technical challenges**: Delivering hyperthermia to deep-seated tumors can be technically challenging, requiring specialized equipment and expertise. 2. **Toxicity**: Hyperthermia can cause toxicity, such as burns, pain, and fatigue, which must be carefully managed. 3. **Tumor heterogeneity**: The response to combined hyperthermia and radiotherapy can vary depending on the tumor type, size, and location. In conclusion, combined hyperthermia and radiotherapy is a promising strategy for the management of bulky tumors, offering improved local control, enhanced tumor response, and potentially increased survival. However, technical challenges, toxicity, and tumor heterogeneity must be carefully considered and addressed to optimize treatment outcomes. | The combination of hyperthermia and radiotherapy is indeed considered an appropriate strategy for the management of certain bulky tumors. Hyperthermia involves raising the temperature of tumor tissues, typically to 40-43°C, which can enhance the effectiveness of radiotherapy. This combination can improve tumor oxygenation, increase blood flow, and make cancer cells more sensitive to the effects of radiation. Several studies have shown that hyperthermia can enhance the therapeutic effects of radiotherapy, particularly in tumors that are large or have poor blood supply, which are often more resistant to conventional treatments. The synergistic effect can lead to better local control of the tumor and potentially improve overall treatment outcomes. However, the appropriateness of this strategy depends on various factors, including the type and location of the tumor, the patient's overall health, and the availability of specialized equipment and expertise. It is important for treatment to be tailored to the individual patient, and decisions should be made in consultation with a multidisciplinary team of healthcare professionals In summary, while combined hyperthermia and radiotherapy can be an effective approach for managing bulky tumors, it is not universally applicable and should be considered on a case-by-case basis. |
| 21 | Can hyperthermia be safely and effectively administered concurrently with chemotherapy? | | |
|  | DeepSeek-V3 | Llama-3.3-70B-Instruct | GPT-4o |
|  | Yes, **hyperthermia** can be safely and effectively administered concurrently with chemotherapy, and this combination is known as **thermochemotherapy**. Clinical evidence suggests that hyperthermia enhances the efficacy of certain chemotherapeutic agents while potentially reducing side effects. Here is a breakdown of key points: ### **Mechanisms of Synergy**  1. **Enhanced Drug Uptake & Penetration** - Heat increases tumor blood flow and vascular permeability, improving drug delivery.  2. **Increased Cytotoxicity** - Some chemotherapies (e.g., cisplatin, doxorubicin, mitomycin C) are more effective at elevated temperatures (40-43°C).  3. **Inhibition of DNA Repair** - Hyperthermia impairs cancer cells' ability to repair chemotherapy-induced DNA damage.  4. **Immunomodulation** - Heat stress can stimulate immune responses against tumors.  ### **Clinical Applications**  - **Local Hyperthermia** (e.g., for breast, cervical, or soft tissue tumors) combined with chemotherapy has shown improved response rates.   - **Regional Hyperthermia** (e.g., for peritoneal carcinomatosis or deep-seated tumors) is used with intraperitoneal or systemic chemotherapy.  - **Whole-Body Hyperthermia** (limited use due to risks) may be combined with systemic chemo in select cases.  ### **Safety Considerations**  - **Temperature Control**: Must be carefully monitored (typically 40-43°C) to avoid normal tissue damage.  - **Chemo Selection**: Some drugs (e.g., platinum-based agents, alkylators) synergize well, while others (e.g., bleomycin) may have increased toxicity.  - **Patient Selection**: Not suitable for patients with cardiovascular instability or severe comorbidities.  ### **Evidence from Studies**  - **Randomized Trials**: Some studies (e.g., in cervical cancer, sarcoma, and recurrent breast cancer) show improved survival and local control when hyperthermia is added to chemo/radiation.  - **Meta-Analyses**: Suggest hyperthermia + chemo improves outcomes in certain cancers with acceptable toxicity.  ### **Conclusion**  When properly administered by experienced clinicians, **thermochemotherapy is a safe and effective strategy** for select cancers, enhancing chemo efficacy without significantly increasing side effects. However, individualized treatment planning and monitoring are essential.  Would you like details on specific cancer types or protocols? | Hyperthermia, which involves exposing cancer cells to high temperatures, can be safely and effectively administered concurrently with chemotherapy in many cases. This combination therapy is often referred to as thermochemotherapy. Research has shown that hyperthermia can enhance the effectiveness of chemotherapy in several ways: 1. **Increased drug delivery**: Hyperthermia can increase blood flow to the tumor, allowing more chemotherapy to reach the cancer cells. 2. **Enhanced drug uptake**: Heat can increase the permeability of cancer cells, making them more receptive to chemotherapy. 3. **Increased sensitivity**: Hyperthermia can make cancer cells more sensitive to the cytotoxic effects of chemotherapy. 4. **Synergistic effects**: Hyperthermia and chemotherapy can have synergistic effects, meaning that the combination of both treatments can be more effective than either treatment alone. Several studies have demonstrated the safety and efficacy of concurrent hyperthermia and chemotherapy in various types of cancer, including: 1. **Locally advanced breast cancer**: A randomized trial showed that hyperthermia combined with chemotherapy improved overall survival and local control compared to chemotherapy alone. 2. **Ovarian cancer**: A phase III trial demonstrated that hyperthermia plus chemotherapy improved progression-free survival and overall survival compared to chemotherapy alone. 3. **Cervical cancer**: A randomized trial showed that hyperthermia combined with chemotherapy and radiation improved local control and overall survival compared to chemotherapy and radiation alone. 4. **Soft tissue sarcoma**: A phase II trial demonstrated that hyperthermia plus chemotherapy improved local control and overall survival compared to chemotherapy alone. However, it's essential to note that the safety and efficacy of concurrent hyperthermia and chemotherapy depend on several factors, including: 1. **Temperature and duration**: The optimal temperature and duration of hyperthermia treatment vary depending on the type of cancer and the specific chemotherapy regimen. 2. **Chemotherapy regimen**: The choice of chemotherapy agents and their dosages can affect the safety and efficacy of concurrent hyperthermia and chemotherapy. 3. **Tumor type and location**: The effectiveness of hyperthermia and chemotherapy can vary depending on the type and location of the tumor. 4. **Patient factors**: Patient-related factors, such as overall health, performance status, and comorbidities, can influence the safety and efficacy of concurrent hyperthermia and chemotherapy. To ensure safe and effective administration of concurrent hyperthermia and chemotherapy, it's crucial to: 1. **Monitor temperature and treatment parameters**: Carefully monitor the temperature and treatment parameters to avoid overheating or underheating. 2. **Adjust chemotherapy dosages**: Adjust chemotherapy dosages as needed to minimize toxicity and optimize efficacy. 3. **Monitor patient response**: Closely monitor patient response to treatment, including toxicity and efficacy, to make adjustments as needed. 4. **Collaborate with a multidisciplinary team**: Work with a multidisciplinary team, including medical oncologists, radiation oncologists, and hyperthermia specialists, to develop and implement an effective treatment plan. In summary, concurrent hyperthermia and chemotherapy can be safely and effectively administered in many cases, but it's essential to carefully consider the specific treatment parameters, patient factors, and tumor characteristics to optimize outcomes. | Hyperthermia, which involves raising the temperature of body tissues, is sometimes used as an adjunct treatment to enhance the effectiveness of chemotherapy and radiation therapy. The idea is that higher temperatures can make cancer cells more susceptible to the effects of these treatments. However, the safety and effectiveness of administering hyperthermia concurrently with chemotherapy depend on several factors, including the type of cancer, the specific chemotherapy drugs being used, and the overall health of the patient. **Safety and Effectiveness:** 1. **Type of Cancer:** Hyperthermia is more commonly used for certain types of cancer, such as soft tissue sarcomas, cervical cancer, and some recurrent cancers. Its effectiveness can vary depending on the cancer type and stage. 2. **Chemotherapy Drugs:** Some chemotherapy drugs may have enhanced effects when used with hyperthermia. However, the combination can also increase the risk of side effects, so it is crucial to consider the specific drugs involved. 3. **Patient Health:** The overall health and medical history of the patient play a significant role in determining whether hyperthermia can be safely administered. Patients with certain conditions may not tolerate the increased temperatures well. 4. **Clinical Setting:** Hyperthermia should be administered in a controlled clinical setting by experienced professionals. The equipment and techniques used must be precise to ensure that the targeted tissues reach the desired temperature without causing damage to surrounding healthy tissues. 5. **Research and Trials:** There is ongoing research and clinical trials to better understand the optimal conditions and protocols for combining hyperthermia with chemotherapy. Some studies have shown promising results, but more research is needed to establish standardized guidelines. In conclusion, while hyperthermia can potentially enhance the effectiveness of chemotherapy, it must be carefully planned and monitored. Patients should consult with their oncologist and a multidisciplinary medical team to determine the best treatment approach for their specific situation. |
| 22 | Who founded hyperthermia for cancer treatment? | | |
|  | DeepSeek-V3 | Llama-3.3-70B-Instruct | GPT-4o |
|  | The use of **hyperthermia** (elevating body or tissue temperature to treat cancer) has ancient roots, but its modern scientific development involved multiple contributors. Here are key figures and milestones: 1. **Ancient Origins** “ Hyperthermia-like treatments were used by **Hippocrates (460-370 BCE)** and ancient Egyptians, who applied heat to tumors.  2. **19th Century** “ German physician **Carl D. W. Busch (1826-1881)** documented tumor regression in a sarcoma patient after high fever.  3. **Early 20th Century** “ **William B. Coley (1862-1936)** used fever-inducing toxins ("Coley's toxins") to treat cancer, indirectly supporting hyperthermia™s role.  4. **Modern Hyperthermia (1960s-1980s)** “ Researchers like **Hermann von Ardenne (Germany)** and **John Overgaard (Denmark)** pioneered localized and whole-body hyperthermia techniques.  5. **Technological Advances** “ **Radiologists and oncologists** later refined methods like **radiofrequency, microwave, and magnetic hyperthermia**, integrating it with radiation/chemotherapy.  While no single "founder" exists, these contributions established hyperthermia as an adjunctive cancer therapy. Today, it is studied in **clinical trials** for its ability to sensitize tumors to conventional treatments.  Would you like details on specific hyperthermia techniques? | The concept of using heat to treat cancer, also known as hyperthermia, has been around for thousands of years. The ancient Greeks, Chinese, and Egyptians all used heat to treat various ailments, including cancer. However, the modern concept of hyperthermia as a cancer treatment was developed in the late 19th and early 20th centuries by several researchers, including: 1. William Coley (1862-1936): An American surgeon who is often credited with being the first to use hyperthermia to treat cancer. In the 1890s, Coley used a combination of heat and bacterial toxins to treat sarcomas, a type of cancer. 2. James Ewing (1866-1943): An American pathologist who studied the effects of heat on cancer cells. In the early 20th century, Ewing demonstrated that heat could kill cancer cells and proposed the use of hyperthermia as a cancer treatment. 3. Karl von Ardenne (1905-1992): A German physicist who developed the first hyperthermia machine in the 1950s. Von Ardenne's machine used microwave energy to heat cancer cells, and his work laid the foundation for modern hyperthermia treatments. In the 1970s and 1980s, hyperthermia research gained momentum, and several researchers made significant contributions to the field, including: 1. George M. Hahn (1925-2013): An American radiobiologist who conducted extensive research on hyperthermia and its effects on cancer cells. 2. J. Robert Stauffer (1932-2012): An American engineer who developed new hyperthermia technologies, including the use of radiofrequency energy to heat cancer cells. Today, hyperthermia is a recognized cancer treatment that is used in combination with other therapies, such as radiation and chemotherapy, to treat a variety of cancers, including breast, prostate, and brain cancer. | The use of hyperthermia for cancer treatment has a long history, with its roots tracing back to ancient civilizations. However, in terms of modern medical practice, hyperthermia as a cancer treatment was developed and refined over the 20th century by various researchers and clinicians.  One of the key figures in the development of hyperthermia therapy was Dr. William B. Coley, who in the late 19th and early 20th centuries used heat as part of his cancer treatment regimen, although he is more famously known for his work with bacterial toxins (Coley's toxins).  In the mid-20th century, researchers like Dr. Manfred von Ardenne in Germany and Dr. Suguru Sugahara in Japan made significant contributions to the scientific understanding and clinical application of hyperthermia in oncology.  It's important to note that hyperthermia is often used in combination with other treatments like radiation and chemotherapy, and its development has been a collaborative effort across many researchers and institutions worldwide. |
|  | Likert Scale rating | | |
|  | DeepSeek-V3 | Llama-3.3-70B-Instruct | GPT-4o |
|  | 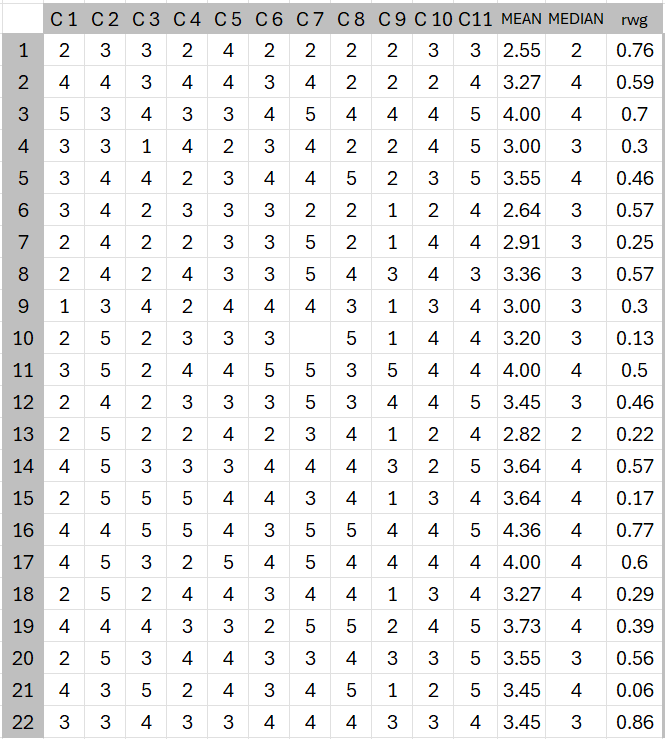 | 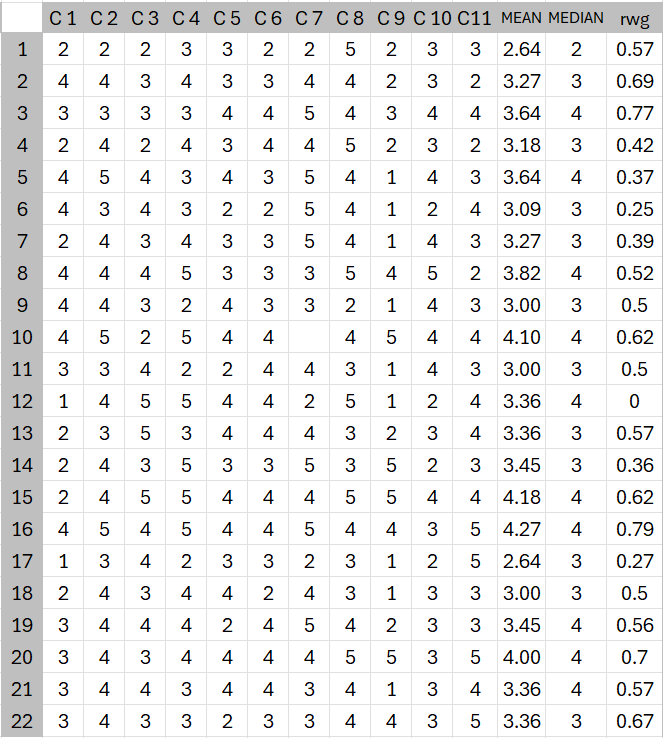 | 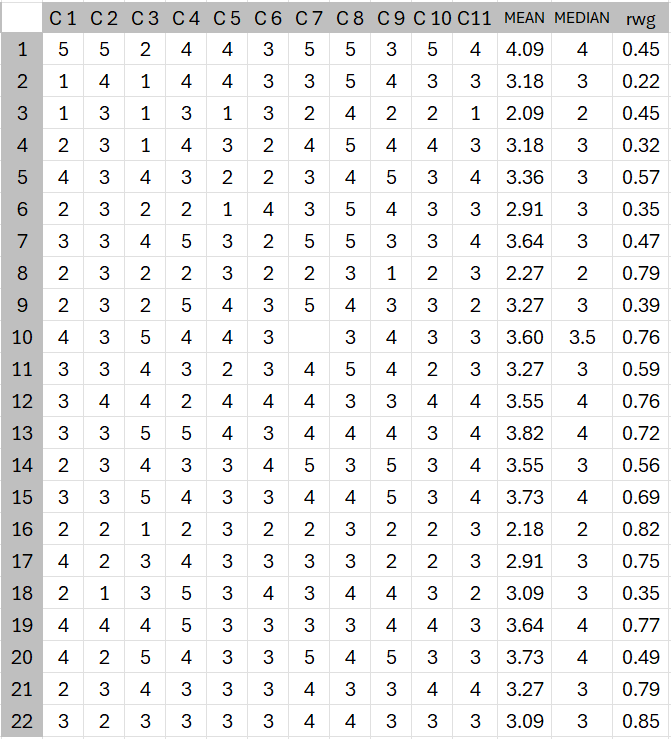 |
|  | Potentially harmful? | | |
|  | DeepSeek-V3 | Llama-3.3-70B-Instruct | GPT-4o |
|  | 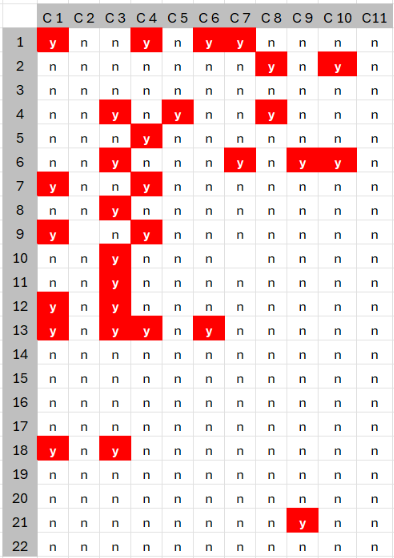 | 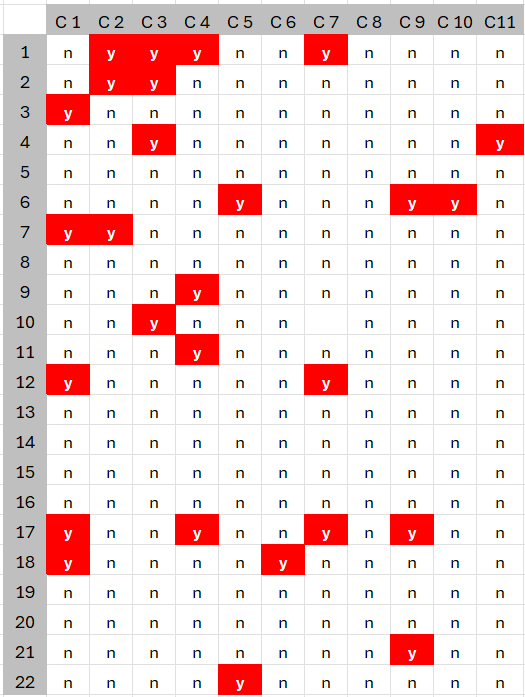 | 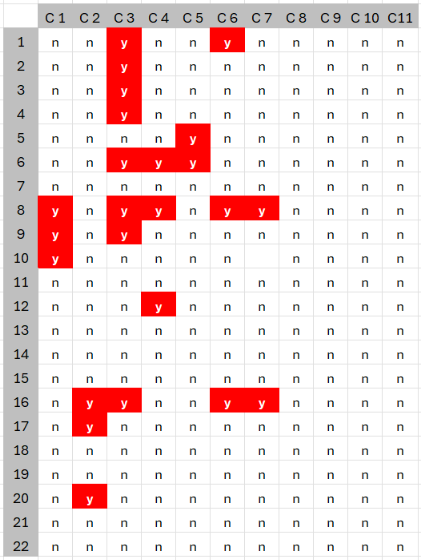 |

Supplementary Table S2.

**Physics questions, responses and ratings:** For each of the 18 physics questions posed to the three large Language Models (LLMs), the three LLM responses are presented side by side for comparison. The left column shows DeepSeek-V3, the middle column Llama-3.3-70B-Instruct, and the right column GPT-4o. The question number is shown in the upper left corner in each case. Subsequently, the ratings of the clinical reviewers are listed for each question and for each LLM. The reviewers are anonymized as P1 – P7. For the Likert scale rating, the quality of each individual response was assessed using a 5-point Likert scale (1 = very bad, 2 = bad, 3 = acceptable, 4 = good, 5 = very good). The Intraclass Correlation Coefficient (ICC) measures interrater agreement and reliability, with values interpreted as poor (<0.5), moderate (0.5-0.75), good (0.75-0.9), and excellent (>0.90). Furthermore, the experts indicated whether they believed an answer could be potentially harmful if used in clinical decision-making. If yes, this is marked with a ‘y = yes’ and highlighted in red.

| 23 | List all physical, chemical and biological effects which must be considered in hyperthermia for treatment planning? | | |
| --- | --- | --- | --- |
|  | DeepSeek-V3 | Llama-3.3-70B-Instruct | GPT-4o |
|  | Hyperthermia treatment planning requires careful consideration of various **physical, chemical, and biological effects** to ensure efficacy and safety. Below is a comprehensive list of key factors: ### **1. Physical Effects**  - **Temperature Distribution**: Uniformity and control of heat deposition in target tissues.  - **Thermal Conductivity & Perfusion**: Heat dissipation due to blood flow and tissue properties.  - **Power Deposition & Penetration Depth**: Efficiency of energy absorption (e.g., RF, microwave, ultrasound).  - **Thermal Diffusion & Convection**: Heat spread due to tissue properties and blood flow.  - **Thermal Dose (CEM43)**: Equivalent minutes at 43°C to quantify biological effect.  - **Hot Spots & Cold Spots**: Risk of overtreatment or undertreatment due to inhomogeneous heating.  - **Thermal Boundary Effects**: Differences in heating at tissue interfaces (e.g., fat vs. muscle).  - **Thermal Monitoring**: Real-time temperature measurement (e.g., thermocouples, MRI thermometry).  ### **2. Chemical Effects**  - **pH Changes**: Acidosis due to increased metabolic activity.  - **Oxygenation & Hypoxia**: Altered oxygen supply affecting treatment sensitivity.  - **Reactive Oxygen Species (ROS)**: Increased oxidative stress leading to cell damage.  - **Protein Denaturation**: Unfolding of proteins at elevated temperatures.  - **Lipid Peroxidation**: Membrane damage due to heat-induced oxidation.  - **Ion Flux Changes**: Disruption of cellular ion homeostasis (e.g., Ca²º, Kº).  ### **3. Biological Effects**  - **Cell Death Mechanisms**: Apoptosis, necrosis, and autophagy induction.  - **Heat Shock Proteins (HSPs)**: Upregulation of protective stress proteins (e.g., HSP70, HSP90).  - **Vascular Effects**: Increased blood flow (hyperemia) or vascular damage (coagulation).  - **Immune Response**: Activation of immune cells (e.g., dendritic cells, macrophages).  - **Radiosensitization & Chemosensitization**: Enhanced effects when combined with radiation/chemotherapy.  - **Tumor Microenvironment**: Hypoxia, acidosis, and interstitial fluid pressure changes.  - **Thermotolerance**: Adaptive resistance due to repeated heating.  - **Tissue Damage Thresholds**: Safe temperature limits for normal vs. tumor tissues.  ### **Conclusion**  Effective hyperthermia treatment planning must integrate these factors to optimize **thermal dose delivery**, **minimize side effects**, and **maximize therapeutic outcomes**. Advanced modeling, real-time monitoring, and combination therapies (e.g., with radiation or immunotherapy) are often employed to address these complexities.  Would you like a deeper dive into any specific aspect? | Hyperthermia, which involves exposing cancer cells to high temperatures to damage or kill them, requires careful consideration of various physical, chemical, and biological effects for effective treatment planning. Here's a comprehensive list of factors to consider: ### Physical Effects 1. **Temperature Distribution**: Achieving uniform temperature distribution within the tumor while minimizing heat exposure to surrounding healthy tissues. 2. **Heat Transfer Mechanisms**: Understanding conduction, convection, and radiation to effectively deliver heat to the target area. 3. **Thermal Dosimetry**: Quantifying the thermal dose (a measure of the cumulative effect of temperature over time) to ensure it is sufficient to kill cancer cells without excessively damaging normal tissues. 4. **Blood Flow**: Considering how blood flow affects heat distribution, as it can both bring heat to and remove heat from the targeted area. 5. **Tissue Properties**: Variations in thermal conductivity, specific heat, and density among different tissues can influence heat penetration and distribution. ### Chemical Effects 1. **pH Changes**: Hyperthermia can alter the pH of the tumor environment, potentially affecting drug delivery and efficacy. 2. **Oxygenation Levels**: Heat can influence oxygen delivery to tissues, impacting the effectiveness of treatments that rely on oxygen, such as radiation therapy. 3. **Drug Activation**: Some chemotherapeutic agents are more effective at higher temperatures, making hyperthermia a potential adjuvant therapy. 4. **Metabolic Changes**: Hyperthermia can alter metabolic pathways within cells, potentially making them more susceptible to damage. ### Biological Effects 1. **Cellular Sensitivity**: Cancer cells are generally more sensitive to heat than normal cells, but the degree of sensitivity can vary among different cell types. 2. **Thermotolerance**: Repeated exposure to heat can induce thermotolerance in some cells, reducing the effectiveness of hyperthermia. 3. **Apoptosis and Necrosis**: Hyperthermia can induce programmed cell death (apoptosis) or immediate cell death (necrosis), depending on the temperature and duration of treatment. 4. **Immune System Activation**: Hyperthermia can stimulate an immune response against tumor cells, potentially enhancing treatment outcomes. 5. **Tumor Microenvironment**: The extracellular matrix, blood vessels, and other components of the tumor microenvironment can influence heat distribution and the biological response to hyperthermia. 6. **Gene Expression Changes**: Heat shock can alter gene expression, potentially affecting cell survival pathways, drug resistance, and the immune response. 7. **Stem Cell Sensitivity**: Cancer stem cells, which are thought to be responsible for tumor recurrence, may have different sensitivities to hyperthermia compared to more differentiated cancer cells. ### Considerations for Treatment Planning - **Imaging and Monitoring**: Using techniques like MRI, CT, or ultrasound to monitor temperature distribution and tumor response during treatment. - **Personalized Medicine**: Tailoring hyperthermia protocols based on individual patient characteristics, such as tumor type, size, location, and blood flow patterns. - **Combination Therapies**: Integrating hyperthermia with other treatments like chemotherapy, radiation therapy, or immunotherapy to enhance efficacy. - **Side Effects Management**: Minimizing and managing potential side effects, such as pain, burns, or damage to surrounding tissues. By carefully considering these physical, chemical, and biological effects, healthcare providers can develop more effective and personalized hyperthermia treatment plans for cancer patients. | Hyperthermia, the therapeutic use of heat to treat cancer, involves a complex interplay of physical, chemical, and biological effects. When planning hyperthermia treatment, several factors must be considered to optimize efficacy and minimize side effects. Here are the key effects in each category: ### Physical Effects: 1. **Temperature Distribution**: Ensuring uniform heating of the target tissue while sparing surrounding healthy tissue. 2. **Heat Delivery Method**: Selection of appropriate technology (e.g., microwave, radiofrequency, ultrasound) to achieve desired temperature. 3. **Thermal Conductivity**: Understanding how heat spreads through different tissues. 4. **Blood Flow**: Increased blood flow can dissipate heat, affecting temperature distribution. 5. **Tissue Perfusion**: Variations in blood supply can lead to uneven heating. 6. **Thermal Dosimetry**: Accurate measurement and control of temperature over time. ### Chemical Effects: 1. **pH Changes**: Heat can alter the pH of tissues, affecting cellular metabolism and drug efficacy. 2. **Reactive Oxygen Species (ROS) Production**: Heat can increase ROS, leading to oxidative stress and cell damage. 3. **Protein Denaturation**: Elevated temperatures can cause proteins to unfold, affecting cellular functions. 4. **Lipid Membrane Fluidity**: Heat can alter cell membrane properties, impacting permeability and signaling. ### Biological Effects: 1. **Cell Death Mechanisms**: Induction of apoptosis, necrosis, or other forms of cell death in cancer cells. 2. **Heat Shock Proteins (HSPs)**: Upregulation of HSPs can protect cells from heat-induced damage and affect immune responses. 3. **Immune Response Modulation**: Hyperthermia can enhance immune system activity against tumor cells. 4. **Tumor Microenvironment**: Changes in the tumor microenvironment, such as increased oxygenation, can affect treatment outcomes. 5. **Cell Cycle Effects**: Heat can disrupt the cell cycle, making cells more susceptible to other treatments like radiation or chemotherapy. 6. **Angiogenesis**: Heat can influence the formation of new blood vessels, impacting tumor growth and treatment response. Considering these effects is crucial for effective hyperthermia treatment planning, as they influence both the direct impact on tumor cells and the overall therapeutic outcome. |
| 24 | How should quality assurance (QA) be performed for a superficial hyperthermia device? | | |
|  | DeepSeek-V3 | Llama-3.3-70B-Instruct | GPT-4o |
|  | Quality assurance (QA) for a **superficial hyperthermia device** is critical to ensure patient safety, treatment efficacy, and regulatory compliance. Below is a structured approach to performing QA for such devices: ### **1. Regulatory & Standards Compliance** - Ensure compliance with relevant standards (e.g., **IEC 60601-2-35** for medical electrical equipment, **AAMI/ANSI HE48** for hyperthermia devices). - Follow **FDA 510(k)** or **CE Marking** requirements if applicable. ### **2. Pre-Treatment QA Checks** #### **A. Device Calibration & Performance Verification** - **Temperature Accuracy**: Verify using a calibrated thermometer or thermal phantom. - **Power Output**: Ensure the device delivers the correct power (W/cm²) as per specifications. - **Uniformity Testing**: Check heat distribution using thermal imaging or phantoms. - **Cooling System**: Verify proper functioning of surface cooling (if applicable). #### **B. Safety Checks** - **Skin Temperature Monitoring**: Ensure sensors are accurate (±0.5°C tolerance). - **Overheating Protection**: Test emergency shut-off mechanisms. - **Electrical Safety**: Verify insulation, grounding, and leakage currents (IEC 60601-1). - **EMI/RFI Interference**: Ensure no interference with other medical devices. ### **3. Treatment-Specific QA** - **Patient-Specific Setup Validation**:  - Correct applicator placement (size, contact with skin).  - Confirmation of treatment parameters (frequency, power, duration). - **Real-Time Monitoring**:  - Continuous skin temperature feedback.  - Automated adjustments to prevent burns. ### **4. Post-Treatment QA** - **Data Logging & Review**:  - Record treatment parameters (time, temperature, power).  - Compare with expected values. - **Patient Feedback & Adverse Event Reporting**:  - Document any discomfort, burns, or unexpected reactions. ### **5. Periodic Maintenance & Long-Term QA** - **Quarterly/Annual Calibration**: Recalibrate sensors and power output. - **Preventive Maintenance**: Inspect cables, cooling systems, and applicators. - **Software Updates**: Ensure firmware is up-to-date with bug fixes. ### **6. Staff Training & Documentation** - **Operator Training**: Ensure proper handling, emergency protocols. - **QA Logs**: Maintain records of all tests, calibrations, and incidents. ### **7. Risk Management (ISO 14971)** - **Failure Mode & Effects Analysis (FMEA)**: Identify potential risks (e.g., overheating, sensor failure). - **Mitigation Strategies**: Implement safeguards (e.g., redundant sensors, alarms). ### **Tools for QA Testing** - **Thermal Phantoms** (e.g., agar-based or liquid-based). - **Infrared Cameras** for heat distribution analysis. - **Calibrated Thermocouples/RTDs** for temperature validation. - **Power Meters** for RF/microwave output verification. ### **Conclusion** A robust QA program for superficial hyperthermia devices ensures **safety, accuracy, and reproducibility** of treatments. Regular testing, staff training, and adherence to standards are essential for optimal performance. Would you like a more detailed checklist for any specific aspect? | Quality assurance (QA) for a superficial hyperthermia device is crucial to ensure the safe and effective delivery of heat to tumors or other targeted tissues. Here's a comprehensive outline of how QA should be performed for a superficial hyperthermia device: **Pre-Treatment QA** 1. **Device inspection**: Regularly inspect the device for any signs of wear, damage, or malfunction. 2. **Electrical safety testing**: Perform electrical safety tests, such as leakage current and ground continuity tests, to ensure the device is safe for patient use. 3. **Temperature calibration**: Calibrate the device's temperature sensors to ensure accurate temperature measurements. 4. **Power output verification**: Verify the device's power output to ensure it is within the specified range. **Treatment QA** 1. **Patient positioning and setup**: Verify that the patient is properly positioned and the device is correctly set up for treatment. 2. **Treatment planning**: Review the treatment plan to ensure that the correct temperature, power, and treatment time are used. 3. **Temperature monitoring**: Continuously monitor the temperature during treatment to ensure it remains within the desired range. 4. **Power monitoring**: Monitor the power output during treatment to ensure it remains within the specified range. **Post-Treatment QA** 1. **Treatment log review**: Review the treatment log to ensure that the treatment was delivered as planned. 2. **Temperature and power data analysis**: Analyze the temperature and power data to ensure that the treatment was delivered within the desired ranges. 3. **Patient feedback**: Obtain feedback from the patient regarding any discomfort, pain, or other adverse effects during treatment. **Periodic QA** 1. **Annual calibration**: Perform annual calibration of the device's temperature sensors and power output. 2. **Quarterly quality control checks**: Perform quarterly quality control checks, such as electrical safety testing and temperature calibration, to ensure the device is functioning correctly. 3. **Device maintenance**: Perform regular maintenance tasks, such as cleaning and replacing worn parts, to ensure the device remains in good working condition. **Additional Considerations** 1. **Staff training**: Ensure that all staff members operating the device have received proper training and are familiar with the device's operation and QA procedures. 2. **Patient education**: Educate patients on the treatment procedure, potential side effects, and any necessary precautions. 3. **Documentation**: Maintain accurate and detailed records of all QA activities, treatment plans, and patient data. By following these QA procedures, you can ensure that your superficial hyperthermia device is functioning correctly and safely, and that patients receive effective and safe treatments. | Quality assurance (QA) for a superficial hyperthermia device is crucial to ensure its safety, efficacy, and reliability. Here are some key steps and considerations for performing QA on such a device: 1. **Regulatory Compliance:**  - Ensure the device meets all relevant regulatory standards and guidelines, such as those from the FDA, CE marking in Europe, or other local regulatory bodies.  - Adhere to standards like ISO 13485 for medical device quality management systems. 2. **Device Calibration:**  - Regularly calibrate the device to ensure accurate temperature delivery. This includes checking sensors and control systems.  - Use traceable calibration equipment and maintain records of calibration activities. 3. **Performance Testing:**  - Conduct performance tests to verify that the device delivers the correct temperature uniformly across the treatment area.  - Test under various conditions to simulate different clinical scenarios. 4. **Safety Checks:**  - Perform electrical safety tests to ensure there are no risks of electrical shock or malfunction.  - Check for proper insulation and grounding. 5. **Software Validation:**  - If the device includes software, perform thorough software validation to ensure it functions correctly and safely.  - Test for potential software bugs or vulnerabilities. 6. **Mechanical Inspection:**  - Inspect the physical components of the device for wear and tear, damage, or defects.  - Ensure all moving parts operate smoothly and safely. 7. **User Training and Competency:**  - Ensure that all users are adequately trained in operating the device safely and effectively.  - Provide clear instructions and training materials. 8. **Documentation and Record Keeping:**  - Maintain detailed records of all QA activities, including test results, calibration data, and maintenance logs.  - Document any incidents or malfunctions and the corrective actions taken. 9. **Routine Maintenance:**  - Establish a routine maintenance schedule to keep the device in optimal working condition.  - Replace any worn or defective parts promptly. 10. **Feedback and Continuous Improvement:**  - Collect feedback from users to identify potential issues or areas for improvement.  - Implement a continuous improvement process to enhance device performance and safety. 11. **Risk Management:**  - Conduct a risk assessment to identify potential hazards associated with the device.  - Implement risk mitigation strategies and regularly review them. By following these steps, you can ensure that a superficial hyperthermia device operates safely and effectively, providing the intended therapeutic benefits to patients. |
| 25 | How should QA be performed for the treatment planning software of superficial hyperthermia device? | | |
|  | DeepSeek-V3 | Llama-3.3-70B-Instruct | GPT-4o |
|  | Quality Assurance (QA) for **treatment planning software (TPS)** in **superficial hyperthermia devices** is crucial to ensure accurate, safe, and effective treatment delivery. Below is a structured approach to performing QA for such software: ### **1. Software Validation & Commissioning** - **Initial Verification:**   - Ensure the software is correctly installed and licensed.   - Validate against manufacturer specifications and regulatory standards (e.g., IEC 60601-2-35, FDA 510(k) if applicable).  - **Benchmark Testing:**   - Compare software predictions (e.g., temperature distributions, SAR patterns) against experimental or reference data (phantom studies, published benchmarks).   - Verify accuracy of **electromagnetic (EM) and thermal modeling** algorithms.  ### **2. Routine QA Tests** #### **A. Input Data Integrity** - Verify correct import of:   - Patient anatomy (CT/MRI scans, contours).   - Applicator positioning and coupling conditions.   - Tissue properties (dielectric, thermal parameters).  #### **B. Treatment Plan Accuracy** - **Dose Calculation Verification:**   - Compare predicted **Specific Absorption Rate (SAR)** and temperature distributions against reference cases.   - Test edge cases (e.g., irregular surfaces, heterogeneous tissues).  - **Applicator Modeling:**    - Ensure correct representation of EM field patterns for different applicators.   - Validate against measured data (e.g., SAR measurements in phantoms).  #### **C. System Performance** - **Algorithm Consistency:**   - Re-run known cases to check reproducibility.   - Test convergence of thermal and EM solvers.  - **Numerical Stability:**   - Check for artifacts (e.g., unrealistic hot/cold spots).   - Verify mesh resolution sensitivity.  #### **D. Safety & Clinical Feasibility** - **Overheating Prevention:**   - Ensure software flags unsafe temperature thresholds.   - Validate **thermal dose limits** (e.g., CEM43).  - **Treatment Time & Power Optimization:**   - Verify that suggested power levels and durations are clinically reasonable.  ### **3. User Interface & Workflow Testing** - **Usability:**   - Ensure intuitive navigation, correct display of isotherms/SAR maps.  - **Data Export/Import:**   - Verify compatibility with DICOM, PACS, or treatment records.  - **Error Handling:**   - Test software response to invalid inputs (e.g., missing contours, unrealistic parameters).  ### **4. Documentation & Compliance** - **Logging & Audit Trails:**   - Ensure software records all user actions and plan modifications.  - **Regulatory Compliance:**   - Maintain records for audits (e.g., FDA, CE, ISO 13485).  ### **5. Frequency of QA** - **Pre-Treatment Checks:**   - Validate each new patient plan against measured data (if possible).  - **Periodic Testing:**   - Monthly: Full validation of key functionalities.   - After Software Updates: Re-verify critical algorithms.  ### **6. Tools for QA** - **Phantom Studies:**   - Use tissue-equivalent phantoms with embedded thermocouples or infrared cameras.  - **Independent Software Checks:**   - Cross-validate with open-source EM/thermal solvers (e.g., COMSOL, Sim4Life).  ### **Conclusion** A robust QA program for superficial hyperthermia TPS should combine **pre-clinical validation, routine checks, and user training** to ensure both technical accuracy and clinical safety. Collaboration with medical physicists and engineers is essential for maintaining compliance and optimizing treatment outcomes. Would you like specific protocols for a particular hyperthermia system (e.g., BSD Medical, Celsius TCS)? | Performing Quality Assurance (QA) for the treatment planning software of a superficial hyperthermia device is crucial to ensure the accuracy and reliability of the treatment plans. Here's a comprehensive outline of how QA should be performed: **Pre-QA Checklist** 1. **Review documentation**: Familiarize yourself with the treatment planning software's user manual, technical specifications, and any relevant regulatory guidelines (e.g., FDA, IEC, or ASTM standards). 2. **Verify software version**: Ensure that the software version being used is the latest and has been properly installed, configured, and validated. 3. **Check hardware compatibility**: Verify that the computer hardware and peripherals (e.g., monitors, printers) meet the software's system requirements. **QA Tests** 1. **Geometric Accuracy**: * Test the software's ability to accurately represent the patient's anatomy and the hyperthermia device's geometry. * Verify that the software correctly calculates distances, angles, and volumes. 2. **Dose Calculation**: * Validate the software's dose calculation algorithm using known test cases or benchmark datasets. * Check the accuracy of dose calculations for various hyperthermia treatment protocols. 3. **Treatment Plan Verification**: * Create test treatment plans with varying parameters (e.g., frequency, power, treatment time). * Verify that the software correctly generates treatment plans and calculates dose distributions. 4. **Data Import/Export**: * Test the software's ability to import and export data from various sources (e.g., CT, MRI, DICOM). * Verify that data is accurately transferred and reformatted. 5. **User Interface and Workflow**: * Evaluate the software's user interface for usability, intuitiveness, and consistency. * Verify that the workflow is logical and efficient. 6. **Error Handling and Reporting**: * Test the software's error handling and reporting mechanisms. * Verify that errors are correctly identified, reported, and logged. 7. **Security and Access Control**: * Evaluate the software's security features, such as user authentication, authorization, and data encryption. * Verify that access controls are in place to prevent unauthorized access or modifications. **Testing Methods** 1. **Manual testing**: Perform manual testing using test cases and datasets to verify the software's functionality. 2. **Automated testing**: Utilize automated testing tools to execute repetitive tests and validate the software's performance. 3. **Comparison with other software**: Compare the treatment planning software with other similar software or benchmark datasets to validate its accuracy.  **Frequency and Schedule** 1. **Initial QA**: Perform an initial comprehensive QA when the software is first installed or updated. 2. **Periodic QA**: Schedule regular QA tests (e.g., quarterly, semiannually) to ensure ongoing software performance and accuracy. 3. **Version updates**: Perform QA tests after each software version update to verify that changes have not introduced new errors or affected existing functionality. **Documentation and Reporting** 1. **QA report**: Generate a detailed QA report documenting the tests performed, results, and any issues or errors encountered. 2. **Corrective actions**: Document any corrective actions taken to address issues or errors found during QA testing. 3. **Software validation**: Maintain a record of software validation, including the QA report, to demonstrate compliance with regulatory requirements. By following this comprehensive QA framework, you can ensure that the treatment planning software for your superficial hyperthermia device is accurate, reliable, and compliant with regulatory requirements. | Quality assurance (QA) for the treatment planning software of a superficial hyperthermia device is crucial to ensure accurate and safe delivery of treatment. Here are some key steps and considerations for performing QA: 1. **Software Validation:**  - **Verification and Validation:** Ensure that the software meets its specifications and intended use. This includes verifying algorithms, dose calculations, and treatment parameters.  - **Testing:** Conduct rigorous testing under various scenarios to ensure the software behaves as expected. This includes unit testing, integration testing, and system testing. 2. **Data Integrity:**  - **Input Data Verification:** Check the accuracy and integrity of input data, such as patient images and treatment parameters.  - **Output Data Verification:** Ensure that the output data, such as treatment plans and dose distributions, are accurate and reliable. 3. **User Interface:**  - **Usability Testing:** Evaluate the user interface for ease of use, clarity, and error prevention. Ensure that the interface allows for intuitive navigation and input.  - **Error Handling:** Test the software is ability to handle errors gracefully and provide meaningful error messages to the user. 4. **Interoperability:**  - **Compatibility Testing:** Ensure the software is compatible with other systems and devices used in the treatment process, such as imaging systems and treatment delivery devices.  - **Data Exchange:** Verify that data exchange between systems is accurate and secure. 5. **Performance Testing:**  - **Speed and Efficiency:** Test the software is performance in terms of speed and resource usage, especially when handling large datasets or complex treatment plans.  - **Scalability:** Ensure the software can handle increased loads or additional features without degradation in performance. 6. **Security:**  - **Data Security:** Implement measures to protect patient data and treatment plans from unauthorized access or breaches.  - **Access Control:** Ensure that only authorized personnel can access and modify treatment plans. 7. **Documentation:**  - **Comprehensive Documentation:** Maintain detailed documentation of the software is design, functionality, and testing procedures.  - **User Manuals:** Provide clear and comprehensive user manuals and training materials for clinicians and technicians. 8. **Regular Updates and Maintenance:**  - **Software Updates:** Regularly update the software to fix bugs, improve performance, and add new features.  - **Maintenance Schedule:** Establish a routine maintenance schedule to ensure the software remains in optimal working condition. 9. **Clinical Validation:**  - **Clinical Trials:** Conduct clinical trials or studies to validate the software is effectiveness and safety in a clinical setting.  - **Feedback Loop:** Establish a feedback loop with clinicians to gather insights and improve the software based on real-world usage. 10. **Regulatory Compliance:**  - **Compliance with Standards:** Ensure the software complies with relevant regulatory standards and guidelines, such as those from the FDA, CE, or other local regulatory bodies.  - **Audit Trails:** Maintain audit trails for all changes and updates to the software to ensure traceability and accountability. By following these steps, you can ensure that the treatment planning software for a superficial hyperthermia device is reliable, safe, and effective for clinical use. Regular QA processes help in maintaining high standards of patient care and treatment outcomes. |
| 26 | How should QA be conducted for a deep regional hyperthermia system? | | |
|  | DeepSeek-V3 | Llama-3.3-70B-Instruct | GPT-4o |
|  | Quality Assurance (QA) for a **Deep Regional Hyperthermia System (DRHS)** is critical to ensure patient safety, treatment accuracy, and system reliability. Below is a structured approach to conducting QA for such a system: ### **1. Pre-Treatment QA Checks** #### **A. System Calibration & Performance Verification** - **Temperature Accuracy**: Verify using phantoms or reference probes. - **Power Output Validation**: Ensure the system delivers the correct power (e.g., RF/microwave output). - **Frequency Stability**: Confirm the system operates at the intended frequency. - **Coupling Medium Check**: Ensure proper conductivity and temperature of water bolus (if used). #### **B. Patient-Specific QA** - **Treatment Planning Validation**:   - Verify correct patient positioning and applicator placement.   - Ensure proper coupling between applicator and patient.  - **Thermometry System Check**:   - Calibrate invasive/non-invasive temperature probes.   - Verify real-time temperature monitoring accuracy.  ### **2. Periodic QA (Weekly/Monthly)** - **Electromagnetic Field (EMF) Distribution Check**:   - Use phantoms to verify homogeneity and penetration depth.  - **Safety Interlocks & Alarms**:   - Test emergency shut-off, overheating protection, and power failure response.  - **Software & Hardware Integrity**:   - Validate treatment planning software updates.   - Check for hardware wear (e.g., cables, applicators).  ### **3. Annual QA (Comprehensive Testing)** - **Full System Recalibration**:   - Verify all sensors, power delivery, and cooling systems.  - **EMC (Electromagnetic Compatibility) Testing**:   - Ensure no interference with other medical devices.  - **Dosimetry Verification**:   - Compare measured vs. predicted SAR (Specific Absorption Rate).  ### **4. Documentation & Compliance** - **QA Logs**: Maintain records of all tests, calibrations, and incidents.  - **Regulatory Compliance**: Ensure adherence to IEC 60601, AAMI, or local regulatory standards.  - **Staff Training**: Verify that operators are trained in QA procedures.  ### **5. Post-Treatment Review** - **Treatment Data Analysis**:   - Compare planned vs. delivered thermal dose.  - **Adverse Event Reporting**:   - Document any deviations or complications.  ### **Key Considerations** - **Phantom Testing**: Use tissue-equivalent phantoms for reproducible QA.  - **Interdisciplinary Collaboration**: Involve medical physicists, engineers, and clinicians.  - **Risk Management**: Implement corrective actions for any QA failures.  By following this structured QA approach, a **Deep Regional Hyperthermia System** can maintain high treatment accuracy, patient safety, and regulatory compliance. Would you like specific protocols for any of these steps? | Conducting Quality Assurance (QA) for a deep regional hyperthermia system is crucial to ensure the safe and effective delivery of heat to tumors while minimizing damage to surrounding healthy tissues. Here's a comprehensive outline of how QA should be conducted: **Pre-Treatment QA** 1. **System calibration**: Verify that the system is properly calibrated, and all components are functioning within specifications. 2. **Power output measurement**: Measure the power output of the system to ensure it matches the manufacturer's specifications. 3. **Frequency and amplitude stability**: Verify that the system's frequency and amplitude are stable and within the specified range. 4. **Electromagnetic compatibility (EMC)**: Ensure that the system does not interfere with other medical devices or equipment in the treatment area. **Treatment Planning QA** 1. **Patient modeling**: Verify that the patient's anatomy is accurately represented in the treatment planning system. 2. **Treatment plan review**: Review the treatment plan to ensure that it is consistent with the prescribed treatment protocol. 3. **Dose calculation**: Verify that the calculated dose distribution is accurate and consistent with the treatment plan. **Treatment Delivery QA** 1. **Temperature monitoring**: Monitor the temperature at multiple points during treatment to ensure that the desired temperature is achieved and maintained. 2. **Power output monitoring**: Monitor the power output during treatment to ensure that it remains within the specified range. 3. **Treatment time and duration**: Verify that the treatment time and duration are consistent with the treatment plan. 4. **Patient comfort and safety**: Monitor the patient's comfort and safety during treatment, and adjust the treatment parameters as needed. **Post-Treatment QA** 1. **Treatment log review**: Review the treatment log to ensure that all treatment parameters were delivered as planned. 2. **Temperature data analysis**: Analyze the temperature data to ensure that the desired temperature was achieved and maintained. 3. **Dose verification**: Verify that the delivered dose is consistent with the planned dose. 4. **Patient follow-up**: Conduct regular follow-up with the patient to monitor their response to treatment and adjust the treatment plan as needed. **Periodic QA** 1. **Annual system calibration**: Perform annual calibration of the system to ensure that it remains within specifications. 2. **Quarterly system maintenance**: Perform quarterly maintenance on the system to ensure that all components are functioning properly. 3. **Staff training and education**: Provide regular training and education to staff on the safe and effective use of the deep regional hyperthermia system. **Additional Considerations**  1. **Commissioning**: Perform a thorough commissioning process when the system is first installed or after any significant upgrades or repairs. 2. **Quality control (QC) checks**: Perform regular QC checks on the system, including daily, weekly, and monthly checks, to ensure that it is functioning properly. 3. **Patient-specific QA**: Perform patient-specific QA for each treatment, including verification of the treatment plan and monitoring of the treatment delivery. By following this comprehensive QA program, you can ensure the safe and effective delivery of deep regional hyperthermia treatments, which is essential for achieving optimal patient outcomes. | Quality assurance (QA) for a deep regional hyperthermia system is crucial to ensure the safety, efficacy, and reliability of the treatment. Here are some key steps and considerations for conducting QA for such a system: 1. **System Calibration:**  - Regularly calibrate the hyperthermia system to ensure accurate temperature delivery.  - Verify the calibration of temperature sensors and ensure they are functioning correctly. 2. **Equipment Checks:**  - Inspect the applicators, power supply, and control units for any signs of wear or damage.  - Ensure that all cables and connections are secure and functioning properly. 3. **Temperature Monitoring:**  - Validate the accuracy of temperature measurement devices, such as thermocouples or fiber-optic sensors.  - Conduct phantom tests to simulate patient conditions and verify temperature distribution. 4. **Treatment Planning:**  - Review and verify treatment plans to ensure they are tailored to the patient's specific anatomy and tumor location.  - Use imaging modalities (e.g., MRI, CT) to assist in accurate treatment planning. 5. **Safety Protocols:**  - Ensure that all safety interlocks and emergency shut-off mechanisms are operational.  - Regularly review and update safety protocols and procedures. 6. **Performance Testing:**  - Conduct regular performance tests to assess the system's ability to deliver the prescribed thermal dose.  - Use standardized phantoms to evaluate the system's performance under controlled conditions. 7. **Staff Training:**  - Ensure that all operators and clinical staff are adequately trained in the use of the hyperthermia system.  - Provide ongoing education and training to keep staff updated on best practices and new developments. 8. **Documentation and Record Keeping:**  - Maintain detailed records of all QA activities, including calibration, maintenance, and performance tests.  - Document any incidents or deviations from standard operating procedures and the corrective actions taken. 9. **Regulatory Compliance:**  - Ensure that the system and its operation comply with relevant regulatory standards and guidelines.  - Stay informed about updates to regulations and incorporate them into the QA process. 10. **Patient Feedback:**  - Collect and review patient feedback to identify any issues related to comfort or treatment efficacy.  - Use this feedback to make necessary adjustments to the system or treatment protocols. By following these steps, you can help ensure that the deep regional hyperthermia system operates safely and effectively, providing optimal treatment outcomes for patients. Regular QA activities are essential to maintaining the integrity and performance of the system over time. |
| 27 | How should QA be performed for the treatment planning software of deep hyperthermia device? | | |
|  | DeepSeek-V3 | Llama-3.3-70B-Instruct | GPT-4o |
|  | Quality Assurance (QA) for **treatment planning software (TPS)** in **deep hyperthermia devices** is critical to ensure accurate, safe, and effective hyperthermia treatments. Below is a structured approach to performing QA for such software: ### **1. Software Validation & Compliance** - **Regulatory Standards**: Ensure compliance with relevant standards (e.g., IEC 60601-2-25, IEC 62304 for medical device software, FDA 21 CFR Part 820, EU MDR). - **Vendor Qualification**: Verify that the software has been validated by the manufacturer (e.g., through ISO 13485 certification). - **Installation Qualification (IQ)**: Confirm proper installation, including hardware/software compatibility checks. ### **2. Functional Testing** - **Input Data Verification**:  - Ensure accurate import of patient data (CT/MRI scans, contours, and treatment parameters).  - Validate segmentation and registration accuracy (e.g., organ/tumor delineation). - **Algorithm Validation**:  - Verify the accuracy of **SAR (Specific Absorption Rate)** and **temperature distribution** calculations.  - Compare against benchmark datasets (e.g., phantom studies or published reference cases). - **Treatment Plan Evaluation**:  - Check dose conformity, hot/cold spot detection, and safety margins.  - Validate optimization algorithms (e.g., steering, power deposition control). ### **3. Performance Testing**  - **Reproducibility & Consistency**:  - Run identical cases multiple times to ensure consistent results.  - Test edge cases (e.g., extreme patient anatomies, high-power scenarios). - **Speed & Stability**:  - Assess computation time for large datasets.  - Check for crashes or memory leaks under heavy usage. ### **4. Safety & Risk Management** - **Error Handling**:  - Test invalid inputs (e.g., incorrect file formats, missing data).  - Ensure fail-safe mechanisms (e.g., automatic shutdown if unsafe conditions are detected). - **Interoperability**:  - Verify seamless integration with **hyperthermia devices** (e.g., BSD-2000, Celsius TCS).  - Check DICOM/HL7 compatibility for data exchange with other systems. ### **5. User Interface & Workflow Testing** - **Usability**:  - Ensure intuitive navigation and clear visualization (e.g., 3D SAR/temperature maps).  - Validate warnings/alerts for critical parameters (e.g., excessive power, proximity to critical organs). - **Training & Documentation**:  - Confirm that user manuals and training materials are accurate and up-to-date. ### **6. Clinical Validation** - **Phantom Studies**:  - Compare TPS predictions with **measured SAR/temperature distributions** in phantoms. - **Patient-Specific QA**:  - Perform retrospective validation on past treatments (if available).  - Use **infrared thermography** or invasive probes (where applicable) for in vivo validation. ### **7. Periodic QA & Maintenance** - **Regular Audits**:  - Revalidate software after updates or hardware changes.  - Monitor log files for anomalies. - **Backup & Recovery**:  - Ensure automated backups of treatment plans and patient data. ### **8. Documentation & Reporting** - Maintain a **QA log** for all tests, including:  - Test cases, results, deviations, and corrective actions.  - Software version, calibration dates, and responsible personnel. ### **Key Tools for QA** - **Phantoms** (e.g., agar-based, MR/CT-compatible). - **SAR measurement systems** (e.g., fiber-optic thermometry, E-field probes). - **Independent dose calculation tools** (for cross-verification). By following this structured QA approach, you can ensure that the **deep hyperthermia TPS** operates safely, accurately, and reliably in clinical settings. Would you like specific details on any of these steps? | Performing Quality Assurance (QA) for the treatment planning software of a deep hyperthermia device is crucial to ensure the accuracy and reliability of the treatment plans. Here's a comprehensive outline of how QA should be performed: **Pre-QA Checklist** 1. **Documentation**: Review the software's user manual, technical specifications, and any relevant regulatory documents (e.g., FDA clearance). 2. **Training**: Ensure that the QA team has received adequate training on the software and its functionality. 3. **Test Environment**: Set up a test environment that mimics the clinical environment, including the same hardware and software configurations. **QA Tests** 1. **Installation and Configuration**: Verify that the software is installed correctly and configured according to the manufacturer's instructions. 2. **Data Input and Validation**: Test the software's ability to accept and validate patient data, including anatomy, tumor location, and treatment parameters. 3. **Treatment Planning Algorithm**: Verify that the treatment planning algorithm is functioning correctly, including: * Dose calculation accuracy * Temperature distribution prediction * Optimization of treatment parameters (e.g., power, frequency, and duration) 4. **Data Export and Import**: Test the software's ability to export and import treatment plans, including compatibility with other systems (e.g., treatment delivery devices).  5. **User Interface and Workflow**: Evaluate the user interface and workflow to ensure they are intuitive, efficient, and minimize the risk of user error. 6. **Error Handling and Reporting**: Test the software's error handling and reporting capabilities, including: * Error detection and notification * Error logging and reporting * Recovery from errors or system crashes 7. **Security and Access Control**: Verify that the software has adequate security measures, including: * User authentication and authorization * Data encryption and access controls * Audit trails and logging 8. **Performance and Scalability**: Test the software's performance and scalability, including: * Computational speed and efficiency * Memory usage and optimization * Ability to handle large datasets and complex treatment plans 9. **Comparison with Established Treatment Planning Systems**: Compare the software's treatment planning results with established treatment planning systems (e.g., commercial systems or in-house developed systems). 10. **Clinical Workflow Integration**: Test the software's integration with the clinical workflow, including: * Compatibility with other systems (e.g., electronic health records, imaging systems) * Ability to generate treatment plans that can be executed by the deep hyperthermia device **QA Metrics and Acceptance Criteria** 1. **Accuracy**: Treatment planning results should be accurate to within a specified tolerance (e.g., ±5%). 2. **Precision**: Treatment planning results should be precise, with minimal variability between repeated calculations. 3. **Completeness**: The software should be able to generate complete treatment plans, including all necessary parameters and settings. 4. **Consistency**: The software should produce consistent results across different users, datasets, and treatment planning scenarios. 5. **User Acceptance**: The software should meet the needs and expectations of the clinical users, including ease of use, efficiency, and effectiveness. **Frequency and Schedule** QA tests should be performed: 1. **Initially**: Before the software is released for clinical use. 2. **Periodically**: At regular intervals (e.g., quarterly, annually) to ensure ongoing compliance with regulatory requirements and clinical standards. 3. **After Software Updates**: After any software updates or changes to ensure that the updates have not introduced new errors or affected existing functionality. By following this comprehensive QA framework, you can ensure that the treatment planning software for the deep hyperthermia device is accurate, reliable, and safe for clinical use. | Quality assurance (QA) for the treatment planning software of a deep hyperthermia device is crucial to ensure accurate and safe delivery of therapy. Here are some key steps and considerations for performing QA on such software: 1. **Understand the System Requirements:**  - Review the software specifications, user manuals, and regulatory requirements.  - Ensure compliance with relevant standards, such as IEC 60601 for medical electrical equipment and ISO 13485 for medical devices. 2. **Develop a QA Protocol:**  - Create a comprehensive QA protocol that outlines the procedures, frequency, and criteria for testing.  - Include both initial validation and routine checks. 3. **Software Validation:**  - Perform initial validation to ensure the software meets all functional requirements.  - Conduct unit testing, integration testing, and system testing.  - Validate algorithms used for treatment planning, such as those for thermal dose calculations and electromagnetic field distributions. 4. **Data Integrity and Security:**  - Verify that patient data is handled securely and that data integrity is maintained.  - Test data import/export functions and ensure compliance with data protection regulations like HIPAA. 5. **User Interface Testing:**  - Evaluate the user interface for usability and accessibility.  - Ensure that the software provides clear and accurate information to the user. 6. **Accuracy and Precision Testing:**  - Test the accuracy of the treatment planning calculations.  - Use phantom models to simulate treatment scenarios and compare the software is output with expected results. 7. **Interoperability Testing:**  - Ensure the software can effectively communicate with other systems, such as imaging devices and treatment delivery systems.  - Test compatibility with different hardware configurations. 8. **Performance Testing:**  - Assess the software is performance under various conditions, including stress testing for high workloads.  - Monitor response times and resource usage. 9. **Error Handling and Safety Features:**  - Test the software is ability to handle errors gracefully and provide meaningful error messages.  - Verify the implementation of safety features, such as treatment interruption and emergency stop functions. 10. **Routine QA Checks:**  - Establish a schedule for routine QA checks, including daily, weekly, and monthly tests.  - Include checks for software updates and patches. 11. **Documentation and Reporting:**  - Maintain detailed records of all QA activities, including test results and any corrective actions taken.  - Ensure that documentation is clear, complete, and easily accessible for audits. 12. **Training and Competency:**  - Ensure that all personnel involved in QA are adequately trained and competent.  - Provide ongoing training to keep staff updated on new features and procedures. 13. **Feedback and Continuous Improvement:**  - Implement a system for collecting user feedback and addressing any issues that arise.  - Use feedback to continuously improve the software and QA processes. By following these steps, you can help ensure that the treatment planning software for a deep hyperthermia device operates safely and effectively, providing reliable support for patient treatments. |
| 28 | How should QA be conducted for a capacitive hyperthermia system? | | |
|  | DeepSeek-V3 | Llama-3.3-70B-Instruct | GPT-4o |
|  | Quality Assurance (QA) for a **Capacitive Hyperthermia System** (CHS) involves rigorous testing to ensure safety, accuracy, and effectiveness in delivering controlled heat therapy. Below is a structured approach to conducting QA for such a system: ### **1. Pre-Testing Checks** - **Documentation Review**: Verify compliance with regulatory standards (e.g., IEC 60601, FDA, CE). - **System Calibration**: Ensure temperature sensors, RF generators, and control algorithms are calibrated. - **Safety Interlocks**: Test emergency shut-off mechanisms and patient protection features. ### **2. Performance Testing** - **Temperature Accuracy & Uniformity**   - Use **phantom models** (tissue-equivalent materials) to measure heat distribution.   - Validate against predefined treatment parameters (e.g., 40-45°C for hyperthermia).   - Check for hot spots using **infrared thermography** or **fiber-optic probes**. - **RF Power & Frequency Stability**   - Measure output power consistency (e.g., 13.56 MHz or 27.12 MHz for capacitive systems).   - Ensure minimal leakage radiation (EMI/EMC testing).  - **Electrode & Coupling Verification**   - Test electrode contact quality (impedance matching).   - Ensure uniform energy distribution between electrodes.  ### **3. Safety & Reliability Testing** - **Patient Safety**   - Skin temperature monitoring to prevent burns.   - Check for electrical isolation (no stray currents).   - Validate alarms for overheating or malfunction.  - **System Robustness**   - Stress testing under prolonged operation.   - Fail-safe mechanisms (e.g., automatic shutdown on fault detection).  ### **4. Software & Control Validation** - **Treatment Planning Software**   - Verify dose calculation accuracy.   - Test user interface for correct parameter input/output.  - **Real-Time Monitoring**   - Ensure feedback loops adjust power dynamically based on temperature readings.  ### **5. Post-Treatment Evaluation** - **Data Logging & Reporting**   - Confirm session records (time, temperature, power levels) are accurate.   - Check for reproducibility across multiple sessions.  ### **6. Periodic Maintenance & Recalibration** - Schedule routine checks for electrode degradation, sensor drift, and software updates.  ### **Standards & References**  - **IEC 60601-2-35** (Medical electrical equipment - Part 2-35: Particular requirements for the basic safety and essential performance of heating devices using RF or microwave energy).  - **ISO 14708-3** (Active implantable medical devices).  ### **Conclusion**  A robust QA program for a capacitive hyperthermia system should combine **performance validation, safety checks, and regulatory compliance** to ensure reliable and safe clinical application. Regular audits and phantom-based testing are essential for maintaining system integrity.  Would you like specific protocols for any of these steps? | Conducting Quality Assurance (QA) for a capacitive hyperthermia system is crucial to ensure its safe and effective operation. Here's a comprehensive outline of the QA process: **Pre-Treatment QA** 1. **System calibration**: Verify that the system is properly calibrated to deliver the desired temperature and power output. 2. **Electromagnetic compatibility (EMC) testing**: Ensure that the system does not interfere with other medical devices or equipment in the treatment area. 3. **Safety inspections**: Check for any signs of wear, damage, or malfunction in the system, including the applicator, cables, and control unit. 4. **Patient preparation**: Verify that the patient is properly prepared for treatment, including removal of metal objects, jewelry, and other contraindications. **Treatment QA** 1. **Power output verification**: Measure the power output of the system to ensure it matches the prescribed treatment parameters. 2. **Temperature monitoring**: Verify that the system is accurately monitoring and controlling the temperature during treatment. 3. **Applicator positioning**: Ensure that the applicator is properly positioned and secured to deliver the desired treatment. 4. **Patient monitoring**: Continuously monitor the patient's vital signs and comfort level during treatment. **Post-Treatment QA** 1. **Treatment log review**: Review the treatment log to ensure that the prescribed treatment parameters were delivered accurately. 2. **Patient assessment**: Assess the patient's response to treatment, including any adverse effects or complications. 3. **System performance evaluation**: Evaluate the system's performance during treatment, including any issues or errors that occurred. 4. **Maintenance and repair**: Perform routine maintenance and repair tasks as needed to ensure the system remains in good working order. **Periodic QA** 1. **Annual system calibration**: Perform annual calibration of the system to ensure accuracy and precision. 2. **Quarterly safety inspections**: Conduct quarterly safety inspections to identify any potential issues or hazards. 3. **Staff training and education**: Provide ongoing training and education to staff on the safe and effective operation of the system. 4. **Quality control checks**: Perform regular quality control checks on the system, including verification of power output, temperature monitoring, and applicator positioning. **Additional Considerations** 1. **Regulatory compliance**: Ensure that the QA process complies with relevant regulatory requirements, such as FDA guidelines or international standards (e.g., IEC 60601-1). 2. **Clinical trial protocols**: If the system is being used in a clinical trial, ensure that the QA process is integrated into the trial protocol and follows Good Clinical Practice (GCP) guidelines. 3. **Risk management**: Identify and mitigate potential risks associated with the system, including patient safety risks, equipment failure, and user error. By following this comprehensive QA process, you can ensure that your capacitive hyperthermia system is operating safely and effectively, and that patients receive high-quality care. | Quality assurance (QA) for a capacitive hyperthermia system is crucial to ensure its safety, efficacy, and reliability. Here are some key steps and considerations for conducting QA on such a system: 1. **Understand System Specifications:**  - Review the manufacturer's specifications and guidelines for the system.  - Understand the intended use, operating parameters, and safety features. 2. **Develop a QA Protocol:**  - Create a detailed QA protocol that outlines the procedures, frequency, and criteria for testing.  - Include both routine checks and comprehensive evaluations. 3. **Calibration:**  - Regularly calibrate the system to ensure accurate temperature delivery and control.  - Use calibrated thermometers and sensors to verify temperature readings. 4. **Electrical Safety Checks:**  - Perform electrical safety tests to ensure there are no risks of electrical shock or malfunction.  - Check for proper grounding and insulation. 5. **Functional Testing:**  - Test all system functions, including power settings, timers, and alarms.  - Verify that the system can achieve and maintain the desired temperature range. 6. **Performance Testing:**  - Conduct performance tests to ensure uniform heating and proper energy distribution.  - Use phantom models or test materials to simulate clinical conditions. 7. **Software Verification:**  - If the system includes software, verify that it functions correctly and is free of bugs.  - Ensure that software updates are tested before implementation. 8. **Safety Features:**  - Test all safety features, such as emergency shut-off, over-temperature alarms, and cooling systems.  - Ensure that all safety interlocks are functioning properly. 9. **Documentation:**  - Maintain detailed records of all QA activities, including test results, calibration data, and maintenance logs.  - Document any issues found and the corrective actions taken. 10. **Training and Competency:**  - Ensure that all personnel involved in operating and maintaining the system are properly trained.  - Conduct regular competency assessments. 11. **Regular Maintenance:**  - Follow the manufacturer's recommended maintenance schedule.  - Replace worn or damaged components as needed. 12. **Review and Update QA Procedures:**  - Regularly review and update QA procedures to incorporate new standards, technologies, or findings.  - Stay informed about industry best practices and regulatory requirements. By following these steps, you can help ensure that a capacitive hyperthermia system operates safely and effectively, providing the intended therapeutic benefits to patients. |
| 29 | How should QA be performed for the treatment planning software of a capacitive hyperthermia device? | | |
|  | DeepSeek-V3 | Llama-3.3-70B-Instruct | GPT-4o |
|  | Quality Assurance (QA) for **treatment planning software (TPS)** in a **capacitive hyperthermia device** is critical to ensure **safety, accuracy, and efficacy** of hyperthermia treatments. Below is a structured approach to performing QA for such software: ### **1. Software Validation & Compliance** - **Regulatory Standards**: Ensure compliance with **IEC 62304** (Medical Device Software), **IEC 60601-1** (Safety), and **FDA/CE/MDR** requirements. - **Vendor Qualification**: Verify that the software has been **validated by the manufacturer** and has proper **documentation (DHR, DMR, IQ/OQ/PQ)**. - **Version Control**: Confirm the correct **software version** is installed and check for **updates/patches**. ### **2. Functional Testing** - **Treatment Plan Accuracy**:  - Verify **dose calculation accuracy** against known test cases (phantom studies).  - Ensure **correct patient anatomy import** (CT/MRI compatibility).  - Validate **electrode positioning and coupling** simulations. - **Algorithm Verification**:  - Check **SAR (Specific Absorption Rate) distribution** accuracy.  - Validate **thermal modeling** (if applicable). - **Interoperability**:  - Test **DICOM import/export** functionality.  - Ensure compatibility with **treatment delivery systems**. ### **3. Safety & Performance Checks** - **Error Handling**:  - Test **invalid input detection** (e.g., incorrect patient data, unrealistic parameters).  - Verify **alarm/warning systems** for unsafe conditions (e.g., overheating, poor electrode contact). - **Treatment Interruption & Recovery**:  - Simulate **power failures** or **software crashes** to ensure safe recovery. - **User Access Control**:  - Verify **role-based permissions** (e.g., only authorized users can modify treatment plans). ### **4. Usability & Workflow Testing** - **User Interface (UI) Evaluation**:  - Ensure **intuitive navigation** and **clear parameter inputs**.  - Check for **real-time feedback** (e.g., temperature distribution visualization). - **Workflow Efficiency**  - Test **planning time** and **ease of adjustments**.  - Validate **automated vs. manual mode** consistency. ### **5. Data Integrity & Security** - **Patient Data Protection**:  - Ensure **HIPAA/GDPR compliance** (encryption, audit logs). - **Backup & Recovery**:  - Test **data backup** procedures and **restoration** after failure. ### **6. Clinical Validation & Benchmarking** - **Phantom Studies**:  - Compare **simulated vs. measured** temperature distributions. - **Clinical Correlation**:  - Review **patient treatment outcomes** to ensure consistency with TPS predictions. ### **7. Periodic QA & Maintenance** - **Scheduled Revalidation**:  - Perform **annual/biannual software checks**. - **Change Management**:  - Document and test any **software updates or patches**. - **User Training & Feedback**:  - Ensure staff are trained and report any anomalies. ### **8. Documentation & Reporting** - **QA Logs**:  - Maintain records of **tests, deviations, and corrective actions**. - **Audit Trail**:  - Track **user actions, parameter changes, and system events**. ### **Conclusion** A robust QA program for **capacitive hyperthermia TPS** should combine **functional testing, safety validation, usability assessment, and ongoing monitoring** to ensure **reliable and safe treatment delivery**. Collaboration between **medical physicists, engineers, and clinicians** is essential for effective QA implementation. Would you like a specific checklist or risk assessment template for this process? | Performing Quality Assurance (QA) for the treatment planning software of a capacitive hyperthermia device is crucial to ensure the accuracy, safety, and efficacy of the treatment. Here's a comprehensive approach to QA for the treatment planning software: **Pre-QA Activities** 1. **Review of documentation**: Review the software's user manual, technical documentation, and any relevant regulatory documents (e.g., FDA 510(k) clearance) to understand the software's functionality, limitations, and intended use. 2. **Familiarization with the device**: Understand the capacitive hyperthermia device's hardware and software components, including the treatment planning software, to ensure a comprehensive QA approach. **QA Tests** 1. **Verification of input parameters**: Validate that the software correctly accepts and processes input parameters, such as patient anatomy, tumor location, and treatment settings (e.g., frequency, power, and duration). 2. **Treatment plan generation**: Verify that the software generates treatment plans that are consistent with the input parameters and device capabilities. 3. **Dose calculation and distribution**: Validate the accuracy of the dose calculation and distribution algorithms, including the calculation of specific absorption rate (SAR) and temperature distributions. 4. **Comparison with benchmark cases**: Compare the software's output with benchmark cases or published data to ensure accuracy and consistency. 5. **Testing of boundary conditions**: Test the software's behavior at boundary conditions, such as maximum and minimum input values, to ensure robustness and error handling. 6. **User interface and usability**: Evaluate the user interface for clarity, consistency, and ease of use, ensuring that it does not introduce errors or inconsistencies. 7. **Data export and import**: Verify that the software correctly exports and imports data, including treatment plans and patient information. 8. **Compatibility testing**: Test the software's compatibility with different operating systems, hardware configurations, and network environments. **Specific QA Tests for Capacitive Hyperthermia** 1. **Electromagnetic field simulations**: Validate the software's electromagnetic field simulations, including the calculation of electric field distributions and SAR. 2. **Thermal modeling**: Verify the accuracy of the thermal modeling algorithms, including the calculation of temperature distributions and heat transfer. 3. **Treatment protocol validation**: Validate the software's ability to generate treatment protocols that are consistent with established clinical guidelines and protocols. **QA Tools and Methodologies** 1. **Automated testing tools**: Utilize automated testing tools, such as scripting languages (e.g., Python) or specialized software testing frameworks (e.g., Pytest), to streamline the testing process. 2. **Phantom and simulation studies**: Use phantom and simulation studies to validate the software's performance in a controlled environment. 3. **Clinical data analysis**: Analyze clinical data to validate the software's performance in real-world scenarios. **QA Schedule and Frequency** 1. **Initial QA**: Perform an initial QA before the software is released for clinical use. 2. **Periodic QA**: Schedule periodic QA (e.g., quarterly or annually) to ensure the software remains accurate and reliable over time. 3. **Version control and change management**: Implement version control and change management processes to ensure that any software updates or changes are thoroughly tested and validated. **QA Team and Responsibilities** 1. **QA team composition**: Assemble a QA team with expertise in software testing, medical physics, and hyperthermia treatment. 2. **Responsibilities**: Define clear responsibilities and roles within the QA team, including testing, validation, and documentation. By following this comprehensive QA approach, you can ensure that the treatment planning software for the capacitive hyperthermia device is accurate, reliable, and safe for clinical use. | Quality assurance (QA) for the treatment planning software of a capacitive hyperthermia device is crucial to ensure accurate and safe treatment delivery. Here are some key steps and considerations for performing QA on such software: 1. **Understand the System Requirements:**  - Review the software specifications, user manuals, and regulatory requirements.  - Understand the clinical objectives and how the software integrates with the hardware. 2. **Software Verification and Validation:**  - **Verification:** Ensure the software meets its specified requirements. This includes checking algorithms, data inputs, and outputs.  - **Validation:** Confirm that the software fulfills its intended use in the clinical environment. 3. **Test Plan Development:**  - Develop a comprehensive test plan that includes test cases covering all functional aspects of the software.  - Include scenarios for normal operation, edge cases, and potential failure modes. 4. **Functional Testing:**  - Test the software is ability to create, modify, and store treatment plans.  - Verify the accuracy of dose calculations and treatment parameters.  - Ensure the software correctly interfaces with the device hardware. 5. **Usability Testing:**  - Evaluate the user interface for ease of use, clarity, and error prevention.  - Conduct tests with end-users to identify any usability issues. 6. **Performance Testing:**  - Assess the software is performance under various conditions, including stress testing to ensure stability and reliability.  - Check for response times and processing speed. 7. **Safety and Risk Management:**  - Perform risk analysis to identify potential hazards associated with software use.  - Implement and verify safety features, such as alarms and error messages. 8. **Data Integrity and Security:**  - Ensure the software maintains data integrity during storage, retrieval, and transmission.  - Verify compliance with data protection regulations and standards. 9. **Regression Testing:**  - Conduct regression tests after any software updates or changes to ensure existing functionality is not adversely affected. 10. **Documentation and Reporting:**  - Maintain detailed records of all tests performed, including test conditions, results, and any issues identified.  - Document any corrective actions taken and retesting performed. 11. **Training and Competency:**  - Ensure that all users are adequately trained on the software and understand its capabilities and limitations.  - Regularly update training materials to reflect software updates or changes. 12. **Regular Reviews and Updates:**  - Schedule regular reviews of the QA process to incorporate new findings, technology updates, and regulatory changes.  - Update the software and QA procedures as necessary to maintain compliance and performance. By following these steps, you can help ensure that the treatment planning software for a capacitive hyperthermia device is reliable, safe, and effective for clinical use. |
| 30 | Provide a list of commercially available devices for superficial and deep hyperthermia therapy? | | |
|  | DeepSeek-V3 | Llama-3.3-70B-Instruct | GPT-4o |
|  | Hyperthermia therapy is used in oncology to heat tumors, enhancing the effects of radiation or chemotherapy. It can be classified into **superficial** (treating tissues up to a few centimeters deep) and **deep** (targeting tumors deeper in the body). Below is a list of commercially available devices for both types: ### **Superficial Hyperthermia Devices**  1. **BSD-500 (Pyrexar Medical / BSD Medical)**   - Uses microwave energy (915 MHz or 433 MHz) for localized heating.   - Applicators for breast, chest wall, and skin lesions.  2. **Celsius TCS (Celsius42+ GmbH, Germany)**   - Water-filtered infrared-A (wIRA) hyperthermia.   - Used for superficial tumors (e.g., melanoma, recurrent breast cancer).  3. **HYPERcollar (Medlogix, Italy)**   - Focused microwave hyperthermia for head and neck tumors.  4. **Alba 4D (Pyrexar Medical)**   - Non-contact ultrasound-based hyperthermia for superficial lesions.  5. **OncoTherm EHY-2000 (OncoTherm GmbH, Germany)**   - Capacitive radiofrequency (RF) hyperthermia for superficial and some deep-seated tumors.  ### **Deep Hyperthermia Devices**  1. **BSD-2000 (Pyrexar Medical / BSD Medical)**   - Uses phased-array RF (100-150 MHz) for deep-seated tumors (e.g., pelvic, abdominal).   - Often combined with radiation therapy.  2. **OncoTherm EHY-3000 (OncoTherm GmbH, Germany)**   - Capacitive RF hyperthermia for deep tumors (e.g., prostate, cervix, liver).  3. **Synchrotherm (Medlogix, Italy)**   - RF-based deep hyperthermia system.  4. **HYPERcollar3D (Erasmus MC, Netherlands / Medlogix)**   - Advanced microwave hyperthermia for head and neck cancers.  5. **Sonalleve MR-HIFU (Profound Medical)**   - Magnetic Resonance-guided High-Intensity Focused Ultrasound (MR-HIFU) for deep-seated tumors (e.g., uterine fibroids, bone metastases).  6. **TheraBionic P1 (TheraBionic GmbH, Germany)**   - Non-invasive RF hyperthermia combined with modulated electromagnetic fields for liver tumors.  ### **Emerging & Research-Oriented Systems**  - **Nanotherm (MagForce AG, Germany)** - Magnetic nanoparticle hyperthermia (approved for brain tumors in Europe).  - **Endo-Hyperthermia (Various)** - Intraluminal RF or microwave applicators for esophageal or prostate heating.  ### **Key Considerations**  - **Superficial devices** are typically microwave or infrared-based.  - **Deep hyperthermia** often uses RF, phased-array microwaves, or HIFU.  - Some systems (e.g., BSD-2000, Sonalleve) integrate imaging (MRI/ultrasound) for precise targeting.  Would you like details on specific clinical applications or regulatory approvals for any device? | Here is a list of commercially available devices for superficial and deep hyperthermia therapy: **Superficial Hyperthermia Devices:** 1. **Alba Hyperthermia System** (Alba Hyperthermia, Italy): A microwave-based system for superficial hyperthermia treatment of tumors. 2. **BSD-2000 Hyperthermia System** (BSD Medical, USA): A radiofrequency-based system for superficial hyperthermia treatment of tumors. 3. **Celsius TCS** (Celsius42, Germany): A radiofrequency-based system for superficial hyperthermia treatment of tumors. 4. **Hyperthermia System HT-100** (Pyrexar Medical, USA): A microwave-based system for superficial hyperthermia treatment of tumors. 5. **Oncotherm EHY-2000** (Oncotherm, Hungary): A radiofrequency-based system for superficial hyperthermia treatment of tumors. **Deep Hyperthermia Devices:** 1. **BSD-2000 3D/MR Hyperthermia System** (BSD Medical, USA): A radiofrequency-based system for deep hyperthermia treatment of tumors, integrated with MRI for temperature monitoring. 2. **Celsius TCS-2** (Celsius42, Germany): A radiofrequency-based system for deep hyperthermia treatment of tumors. 3. **Hyperthermia System HT-1000** (Pyrexar Medical, USA): A microwave-based system for deep hyperthermia treatment of tumors. 4. **Oncotherm EHY-3010** (Oncotherm, Hungary): A radiofrequency-based system for deep hyperthermia treatment of tumors. 5. **Sonalleve MR-HIFU** (Profound Medical, Canada): A high-intensity focused ultrasound (HIFU) system for deep hyperthermia treatment of tumors, integrated with MRI for temperature monitoring. **Hybrid Devices (Superficial and Deep Hyperthermia):** 1. **Alba Hyperthermia System** (Alba Hyperthermia, Italy): Can be used for both superficial and deep hyperthermia treatment of tumors. 2. **BSD-2000 Hyperthermia System** (BSD Medical, USA): Can be used for both superficial and deep hyperthermia treatment of tumors. 3. **Oncotherm EHY-2000** (Oncotherm, Hungary): Can be used for both superficial and deep hyperthermia treatment of tumors. | Hyperthermia therapy is a type of medical treatment in which body tissue is exposed to higher temperatures to damage and kill cancer cells or make them more sensitive to the effects of radiation and certain anticancer drugs. There are several devices available for both superficial and deep hyperthermia therapy. Here are some examples: ### Superficial Hyperthermia Devices 1. **BSD-500** - Developed by Pyrexar Medical, this device is used for superficial hyperthermia treatment. It uses microwave energy to heat superficial tumors. 2. **EHY-100** - Manufactured by Oncotherm, this device is designed for superficial hyperthermia and uses capacitive coupling to deliver heat to the targeted area. 3. **ALBA 4D** - Produced by MedLogix, this device is used for superficial hyperthermia and features a flexible applicator for treating various body surfaces. ### Deep Hyperthermia Devices 1. **BSD-2000** - Also from Pyrexar Medical, this device is used for deep regional hyperthermia. It uses phased array technology to focus energy on deep-seated tumors. 2. **EHY-2030** - Another device from Oncotherm, this is used for deep hyperthermia and employs modulated electro-hyperthermia (mEHT) to target deep tumors. 3. **SIGMA-Eye/MR** - Developed by Dr. Sennewald Medizintechnik GmbH, this device is used for deep hyperthermia and is often combined with MRI for precise targeting. 4. **Thermotron RF-8** - Manufactured by Yamamoto Vinita Co., Ltd., this device is used for deep hyperthermia and utilizes radiofrequency energy to heat deep-seated tumors. These devices are typically used in specialized medical centers and require trained personnel to operate. The choice of device depends on the specific medical condition being treated, the location and size of the tumor, and the overall treatment plan. Always consult with a healthcare professional to determine the most appropriate treatment options. |
| 31 | What is the optimal treatment frequency (in MHz) for a superficial hyperthermia device? | | |
|  | DeepSeek-V3 | Llama-3.3-70B-Instruct | GPT-4o |
|  | The optimal treatment frequency for a **superficial hyperthermia device** typically falls within the range of **434 MHz to 915 MHz**, depending on the desired depth of penetration and heating efficiency. Here is a breakdown: 1. **434 MHz**   - **Depth**: Penetrates deeper (~3-4 cm) but with less superficial heating.   - **Use**: Suitable for slightly deeper subcutaneous tumors.  2. **915 MHz**   - **Depth**: More superficial (~1-2 cm) with better energy absorption in shallow tissues.   - **Use**: Preferred for very superficial lesions (e.g., skin cancer, chest wall recurrences).  ### Key Considerations: - **Higher frequencies (e.g., 915 MHz)** provide more localized heating but have limited penetration.  - **Lower frequencies (e.g., 434 MHz)** offer deeper penetration but may require careful control to avoid overheating deeper tissues.  - **Clinical protocols** often use **433-434 MHz in Europe** and **915 MHz in the U.S.** due to regulatory allocations.  ### Optimal Choice: - For **strictly superficial tumors (‰¤1 cm depth)**, **915 MHz** is often optimal.  - For **slightly deeper (1-3 cm) superficial treatments**, **434 MHz** may be more effective.  Would you like guidance on power settings or applicator selection as well? | The optimal treatment frequency for a superficial hyperthermia device depends on several factors, including the depth of the tumor, the size of the treatment area, and the type of tissue being treated. However, based on the physics of electromagnetic heating and clinical experience, the following frequency ranges are commonly used for superficial hyperthermia: * 433 MHz: This frequency is commonly used for superficial hyperthermia, as it provides a good balance between penetration depth and heating efficiency. It is often used for treating tumors located 1-3 cm below the skin surface. * 915 MHz: This frequency is also used for superficial hyperthermia, particularly for treating smaller tumors or those located closer to the skin surface (less than 1 cm deep). It provides a more focused heating pattern and is often used in combination with other frequencies. * 2.45 GHz: This frequency is sometimes used for superficial hyperthermia, particularly for treating very superficial tumors or those located in areas with limited blood flow. However, it has a shorter penetration depth and may not be as effective for deeper tumors. It's worth noting that the optimal frequency may vary depending on the specific application and the individual patient's anatomy and physiology. Some superficial hyperthermia devices may use multiple frequencies or a frequency-hopping approach to optimize heating and minimize hot spots. In general, the choice of frequency for superficial hyperthermia depends on the following factors: * Penetration depth: Lower frequencies (e.g., 433 MHz) penetrate deeper into tissue, while higher frequencies (e.g., 2.45 GHz) have a shorter penetration depth. * Heating efficiency: Higher frequencies tend to heat tissue more efficiently, but may also produce more hot spots. * Tumor size and location: Larger tumors or those located deeper in tissue may require lower frequencies, while smaller tumors or those located closer to the skin surface may be treated with higher frequencies. * Blood flow: Areas with high blood flow may require higher frequencies to achieve adequate heating. Ultimately, the optimal treatment frequency for a superficial hyperthermia device should be determined based on the specific clinical application and the individual patient's needs, and may require experimentation and adjustment during treatment. | Superficial hyperthermia is a treatment method that involves raising the temperature of superficial tissues, typically to enhance the effectiveness of other treatments like radiation therapy. The optimal frequency for superficial hyperthermia devices is generally in the range of 433 MHz to 915 MHz. These frequencies are effective for heating tissues at a superficial level, typically up to a few centimeters deep. The choice of frequency can depend on several factors, including the specific design of the device, the depth of tissue that needs to be targeted, and the type of cancer or condition being treated. It's important for the treatment to be carefully controlled and monitored by medical professionals to ensure safety and effectiveness. Always consult with a healthcare provider or a specialist in hyperthermia treatment for specific recommendations and protocols. |
| 32 | What is the optimal treatment frequency (in MHz) for a deep regional hyperthermia device? | | |
|  | DeepSeek-V3 | Llama-3.3-70B-Instruct | GPT-4o |
|  | The optimal treatment frequency for a **deep regional hyperthermia (DRH) device** typically falls in the range of **70-150 MHz**, with **100-120 MHz** being the most commonly used in clinical practice.  ### **Key Considerations for Frequency Selection:** 1. **Penetration Depth vs. Energy Absorption**   - Lower frequencies (e.g., 70-90 MHz) penetrate deeper but may have lower energy absorption in deep-seated tumors.   - Higher frequencies (e.g., 100-150 MHz) provide better energy absorption but may have slightly reduced penetration.  2. **Clinical Applications**   - **Pelvic and abdominal tumors** (e.g., cervical, prostate, pancreatic cancers) often use **90-120 MHz** for optimal depth and heating efficiency.   - **Thoracic and deep-seated tumors** may benefit from slightly lower frequencies (e.g., 70-100 MHz) for better penetration.  3. **Device-Specific Factors**   - Systems like the **BSD-2000 (Pyrexar/Oncotherm)** typically operate at **100-120 MHz**.   - Some advanced systems allow **frequency modulation** to optimize heating patterns.  ### **Conclusion:**  The **most widely recommended frequency** for deep regional hyperthermia is **around 100-120 MHz**, balancing penetration and effective energy deposition. However, the exact frequency may be adjusted based on tumor location, size, and patient-specific factors.  Would you like details on specific hyperthermia systems or clinical protocols? | The optimal treatment frequency for a deep regional hyperthermia device depends on several factors, including the depth of the tumor, the size of the treatment area, and the type of tissue being treated. However, based on various studies and clinical experiences, the following frequencies are commonly used for deep regional hyperthermia: * 100-200 MHz: This frequency range is often used for deep regional hyperthermia, as it provides a good balance between penetration depth and heating efficiency. Frequencies in this range can heat tumors located at depths of up to 10-15 cm. * 70-100 MHz: This frequency range is sometimes used for deeper tumors, as it provides slightly better penetration depth than higher frequencies. However, the heating efficiency may be lower, and the treatment time may be longer. * 200-300 MHz: This frequency range is often used for more superficial tumors or for smaller treatment areas. Frequencies in this range provide higher heating efficiency, but the penetration depth is lower, typically limited to 5-10 cm. It's worth noting that the optimal frequency may vary depending on the specific device and treatment protocol being used. Some devices may use a single frequency, while others may use a range of frequencies or even multiple frequencies simultaneously. In general, the choice of frequency for deep regional hyperthermia depends on the specific clinical scenario and the goals of treatment. The treatment frequency should be selected to provide the optimal balance between heating efficiency, penetration depth, and treatment time, while minimizing side effects and ensuring patient safety. In the United States, the FDA has approved several deep regional hyperthermia devices for the treatment of various types of cancer, including the BSD-2000 (100 MHz) and the Celsius TCS (70-100 MHz). However, the specific frequency used may vary depending on the device and the treatment protocol. | The optimal treatment frequency for a deep regional hyperthermia device typically ranges from 70 to 150 MHz. This frequency range is chosen because it allows for effective penetration of radiofrequency energy into deep tissues while minimizing absorption by superficial tissues. The exact frequency used can depend on various factors, including the specific device, the size and location of the tumor, and the patient's anatomy. It's important to follow the manufacturer's guidelines and clinical protocols when determining the appropriate frequency for treatment. Additionally, treatment should be administered by trained medical professionals to ensure safety and efficacy. |
| 33 | Which thermometric parameters are most predictive and prognostic for hyperthermia treatment efficacy? | | |
|  | DeepSeek-V3 | Llama-3.3-70B-Instruct | GPT-4o |
|  | The efficacy of hyperthermia treatment (HT) in oncology depends on several thermometric parameters, with the most predictive and prognostic factors being: ### **1. Temperature Metrics**  - **Intratumoral Temperature (Ttumor)**   - **Minimum (Tmin)** - Critical for ensuring no cold spots.   - **Maximum (Tmax)** - Should be controlled to avoid thermal damage.   - **Average (Tave)** - Correlates with treatment efficacy.   - **T90 (Temperature achieved in 90% of measured points)** - Strong predictor of response.   - **T10 (Temperature exceeded in 10% of points)** - Indicates potential overtreatment.   - **Thermal Dose (CEM43)**   - **Cumulative Equivalent Minutes at 43°C (CEM43)** - Integrates time and temperature, strongly linked to cell kill and clinical outcomes.   - Formula:   \[  CEM43 = \int_{0}^{t} R^{(43 - T)} \, dt \quad (R = 0.5 \text{ for } T \geq 43°C, \ R = 0.25 \text{ for } T < 43°C)  \]  - Higher CEM43 values (>10-60 min) correlate with better tumor control. ### **2. Spatial and Temporal Uniformity**  - **Thermal Heterogeneity Index (THI)** - Measures uniformity of heating.   - **Percentage of Tumor Volume Above 40-42°C** - Ensures adequate coverage.  ### **3. Physiological and Perfusion-Based Parameters**  - **Tumor Blood Flow (Perfusion)** - Affects heat dissipation; low perfusion improves heating efficacy.   - **Thermal Washout Rate** - Faster cooling reduces effectiveness.   - **Oxygenation (pO2)** - Hypoxic regions may require higher temperatures.  ### **4. Combined Parameters (Predictive Models)**  - **Thermal Dose + Radiation Dose (TER: Thermal Enhancement Ratio)** - Predicts synergy with radiotherapy.   - **Arrhenius-Based Damage Models** - Estimate cell death based on time-temperature profiles.  ### **Clinical Evidence** - **T90 > 40.5°C** and **CEM43 > 10 min** are strong predictors of improved local control and survival in cancers like cervical, breast, and soft-tissue sarcomas.  - **Tmax > 42.5°C** may enhance immunogenic effects but risks normal tissue damage.  ### **Conclusion** The most **prognostic** thermometric parameters are **T90 and CEM43**, while **thermal uniformity and perfusion** help optimize treatment planning. Real-time thermometry (MRI, fiber optics) is essential for accurate monitoring.  Would you like insights on specific cancer types or thermometry techniques? | Hyperthermia treatment, which involves exposing cancer cells to high temperatures to damage or kill them, relies on precise temperature control and monitoring to ensure efficacy and minimize side effects. The thermometric parameters most predictive and prognostic for hyperthermia treatment efficacy include: 1. **Intratumoral temperature**: The temperature within the tumor itself is a critical parameter. A minimum temperature of 40°C to 41°C is often considered necessary for effective hyperthermia treatment, with temperatures above 42°C being more cytotoxic to cancer cells. 2. **Temperature distribution**: The uniformity of temperature distribution within the tumor is essential. Non-uniform heating can lead to underheated areas, reducing treatment efficacy, and overheated areas, increasing the risk of normal tissue damage. 3. **Temperature duration**: The duration of heating is also important. Prolonged heating times (e.g., 30-60 minutes) can increase the cytotoxic effects of hyperthermia, but may also increase the risk of normal tissue damage. 4. **Cumulative equivalent minutes (CEM)**: CEM is a parameter that takes into account both temperature and duration of heating. It is calculated by integrating the temperature over time and is used to estimate the thermal dose delivered to the tumor. 5. **Thermal dose**: The thermal dose, which is a measure of the total amount of heat energy delivered to the tumor, is a prognostic factor for hyperthermia treatment efficacy. A higher thermal dose is associated with improved treatment outcomes. 6. **Tumor temperature homogeneity**: The homogeneity of temperature distribution within the tumor can be quantified using parameters such as the temperature uniformity index (TUI) or the thermal dose uniformity index (TDUI). 7. **Blood flow and perfusion**: Tumor blood flow and perfusion can affect the delivery of heat to the tumor. Parameters such as blood flow velocity, perfusion rate, and oxygenation status can influence the efficacy of hyperthermia treatment. 8. **Tumor oxygenation**: Tumor oxygenation status can impact the efficacy of hyperthermia treatment. Hypoxic tumors may be more resistant to heat damage, while well-oxygenated tumors may be more susceptible. 9. **pH and lactate levels**: Tumor pH and lactate levels can also influence the efficacy of hyperthermia treatment. Acidic environments (low pH) and high lactate levels can indicate a more aggressive tumor phenotype and may require adjusted hyperthermia treatment protocols. These thermometric parameters can be measured using various techniques, including: 1. **Invasive thermometry**: Using thermocouples or thermistors inserted into the tumor. 2. **Non-invasive thermometry**: Using techniques such as magnetic resonance thermometry (MRT), ultrasound thermometry, or infrared thermography. 3. **Imaging techniques**: Such as positron emission tomography (PET), computed tomography (CT), or magnetic resonance imaging (MRI) to assess tumor temperature, perfusion, and oxygenation. By monitoring and controlling these thermometric parameters, clinicians can optimize hyperthermia treatment protocols to improve treatment efficacy and minimize side effects. | In the context of hyperthermia treatment, which is often used as an adjunct to cancer therapies, several thermometric parameters are considered important for predicting and assessing treatment efficacy. These parameters help in understanding how effectively the hyperthermia treatment is being delivered and its potential impact on the tumor. The most predictive and prognostic thermometric parameters include: 1. **Temperature Distribution**: The spatial distribution of temperature within the tumor and surrounding tissues is crucial. Achieving a uniform temperature distribution ensures that the entire tumor receives adequate heat, which is necessary for effective treatment. 2. **Maximum Temperature (Tmax)**: The highest temperature reached within the tumor is a critical parameter. Higher maximum temperatures are generally associated with better treatment outcomes, as they can enhance the sensitivity of cancer cells to radiation and chemotherapy. 3. **Minimum Temperature (Tmin)**: The lowest temperature within the target area is also important. Ensuring that the minimum temperature is above a certain threshold is necessary to achieve therapeutic effects across the entire tumor. 4. **Average Temperature (Tavg)**: The mean temperature within the tumor can provide an overall assessment of the heat delivered. It is often used to evaluate the consistency and adequacy of the treatment. 5. **Time-Temperature Integral (TTI)**: This parameter considers both the temperature achieved and the duration for which it is maintained. It is a cumulative measure that reflects the total thermal dose delivered to the tumor. 6. **Thermal Dose (CEM43°C T90)**: This is a standardized measure of thermal dose, expressed as the equivalent time at 43°C. The T90 value indicates the temperature exceeded by 90% of the tumor volume, providing a robust measure of treatment adequacy. 7. **Temperature Homogeneity**: The uniformity of temperature distribution is important for ensuring that all parts of the tumor receive sufficient heat. Poor homogeneity can lead to under-treatment of some areas. 8. **Rate of Temperature Increase**: The speed at which the target temperature is reached can influence treatment outcomes. Rapid heating may be more effective in certain contexts but must be balanced against the risk of damage to surrounding healthy tissues. These parameters are typically monitored using invasive or non-invasive thermometry techniques, such as thermocouples, fiber-optic sensors, or magnetic resonance thermometry. The choice of parameters and their target values can vary depending on the specific clinical protocol, tumor type, and treatment goals. |
| 34 | What metrics or indicators can be used to assess the quality of a given hyperthermia session? | | |
|  | DeepSeek-V3 | Llama-3.3-70B-Instruct | GPT-4o |
|  | Assessing the quality of a hyperthermia session involves evaluating both **treatment delivery** and **biological response** to ensure therapeutic efficacy while minimizing adverse effects. Here are key metrics and indicators: ### **1. Temperature-Based Metrics**  - **Target Temperature Range**:   - *Mild Hyperthermia*: 40-42°C (for sensitization to chemo/radiation).   - *Moderate/Abiative Hyperthermia*: >42-45°C (direct cytotoxic effects).  - **Temperature Distribution**:   - **T90**: Temperature exceeded by 90% of monitored points (indicates uniformity).   - **T10**: Temperature exceeded by 10% of points (identifies hotspots).   - **CEM43**: Cumulative equivalent minutes at 43°C (thermal dose metric).  - **Thermal Gradient**: Spatial uniformity (e.g., ‰¤1°C variation in target).  ### **2. Time-Dependent Parameters**  - **Duration at Target Temperature**:   - Typically 30-60 minutes for mild hyperthermia.   - Shorter for higher temperatures (e.g., 4-6 minutes at 50°C for ablation).  - **Heating Rate**: Speed of reaching target temperature (avoiding overshoot).  ### **3. Biological/Physiological Indicators**  - **Tumor Perfusion Changes**:   - Measured via Doppler ultrasound or dynamic contrast-enhanced MRI.  - **Tissue Oxygenation**:   - Hypoxia reduction (e.g., via EPR oximetry or PET imaging).  - **Biomarkers**:   - Heat shock proteins (HSP70, HSP90) as stress response markers.   - Circulating tumor DNA (ctDNA) for treatment efficacy.  ### **4. Imaging & Real-Time Monitoring**  - **MRI Thermometry**: Gold standard for non-invasive temperature mapping.  - **Ultrasound Thermometry**: Less precise but real-time.  - **Infrared Thermography**: Surface temperature monitoring (superficial treatments).  ### **5. Clinical & Safety Metrics**  - **Adverse Events**:   - Skin burns, pain, or systemic effects (e.g., thermal toxicity).  - **Patient Tolerance**:   - Subjective pain scales (e.g., VAS) or sedation requirements.  - **Equipment Performance**:   - Power delivery consistency, applicator positioning accuracy.  ### **6. Post-Treatment Outcomes**  - **Tumor Response**:   - RECIST criteria (imaging) or pathological complete response (pCR).  - **Synergy with Adjuvant Therapies**:   - Enhanced chemo/radiation efficacy (e.g., increased drug uptake).  ### **Summary Table**  \| **Category** \| **Key Metrics** \| **Tools/Methods** \|  \|--------------------\|-----------------------------------------\|---------------------------------------\|  \| **Temperature** \| T90, T10, CEM43, gradient \| MRI thermometry, fiber-optic probes \|  \| **Time** \| Duration at target, heating rate \| Real-time monitoring systems \|  \| **Biological** \| HSPs, perfusion, oxygenation \| Biomarkers, imaging (PET, DCE-MRI) \|  \| **Safety** \| Adverse events, pain scores \| Clinical assessment, patient reports \|  \| **Efficacy** \| Tumor shrinkage, synergy with therapies \| Post-treatment imaging/histopathology \|  Optimal hyperthermia requires balancing these metrics to maximize therapeutic effect while ensuring patient safety. Real-time feedback (e.g., MRI-guided systems) is ideal for precision. | Assessing the quality of a hyperthermia session involves evaluating its effectiveness in achieving the desired therapeutic outcomes while ensuring patient safety. Hyperthermia, which involves exposing cancer cells to high temperatures to damage or kill them, requires careful monitoring to ensure that the treatment is delivered as planned and that the patient tolerates it well. Several metrics and indicators can be used to assess the quality of a hyperthermia session: 1. **Temperature Distribution and Homogeneity**: This is crucial for ensuring that the target tissue (tumor) reaches and maintains the desired temperature (usually between 40°C and 45°C) for the specified duration, while minimizing overheating of surrounding healthy tissues. 2. **Specific Absorption Rate (SAR)**: This measures the amount of electromagnetic energy absorbed by the body when exposed to electromagnetic fields. It's an important parameter for ensuring that the energy deposition is as intended, without causing undue harm to the patient. 3. **Thermal Dose**: This refers to the cumulative effect of temperature over time. It's quantified in terms of equivalent minutes at 43°C (CEM43), which helps in standardizing the thermal dose delivered during hyperthermia sessions. 4. **Tumor Response**: Post-treatment assessment of the tumor's response to hyperthermia, including changes in size, metabolic activity (as measured by PET scans), and histological changes, can indicate the effectiveness of the treatment. 5. **Patient Tolerance and Toxicity**: Monitoring for adverse effects such as pain, burns, or systemic toxicity (e.g., fever, nausea) during and after the treatment session. The severity and incidence of these side effects can impact the quality of the session. 6. **Technical Performance**: This includes the functionality and reliability of the hyperthermia equipment, the accuracy of temperature monitoring systems, and the ability to maintain a stable and controlled thermal environment throughout the session. 7. **Quality of Life (QoL) Metrics**: Assessing changes in the patient's quality of life, such as improvements in symptoms, functional status, and overall well-being, can provide insight into the broader impact of the hyperthermia treatment. 8. **Imaging and Diagnostic Feedback**: Utilizing imaging techniques (e.g., MRI, CT scans) before, during, and after hyperthermia to assess anatomical and functional changes in the tumor and surrounding tissues. This feedback can help in adjusting treatment parameters for subsequent sessions. 9. **Patient Compliance and Satisfaction**: High patient compliance and satisfaction rates can indicate that the treatment is tolerable and effective, contributing to the overall quality of the hyperthermia session. 10. **Multidisciplinary Team (MDT) Review**: Regular review of patient outcomes and treatment delivery by a multidisciplinary team of oncologists, radiologists, physicists, and other healthcare professionals can help in identifying areas for improvement and ensuring that hyperthermia sessions meet high standards of quality. By considering these metrics and indicators, healthcare providers can comprehensively assess the quality of hyperthermia sessions, making adjustments as necessary to optimize treatment outcomes and patient safety. | Assessing the quality of a hyperthermia session, which is often used as a treatment modality in oncology, involves several metrics and indicators to ensure both efficacy and safety. Here are some key factors to consider: 1. **Temperature Monitoring**:   - **Target Temperature**: Achieving and maintaining the desired therapeutic temperature (usually between 40°C and 43°C) in the tumor tissue is crucial.  - **Temperature Distribution**: Uniformity of temperature across the target area should be assessed to ensure effective treatment. 2. **Treatment Duration**:  - The duration for which the target temperature is maintained is critical. Typically, a session lasts between 30 to 60 minutes, depending on the protocol. 3. **Thermal Dose**:  - This is a cumulative measure of the temperature over time, often expressed in equivalent minutes at a reference temperature (e.g., CEM43°C T90, which is the cumulative equivalent minutes at 43°C for 90% of the target volume). 4. **Patient Tolerance and Safety**:  - Monitoring for any adverse effects or discomfort experienced by the patient during the session.  - Ensuring that normal tissues surrounding the target area are not overheated. 5. **Imaging and Monitoring**:  - Use of imaging techniques like MRI or ultrasound to monitor temperature distribution and ensure accurate targeting.  - Real-time feedback systems to adjust parameters as needed. 6. **Technical Parameters**:  - Calibration and performance of the hyperthermia equipment, including applicators and sensors.  - Consistency and reliability of the heating method used (e.g., microwave, radiofrequency, ultrasound). 7. **Biological Response**:  - Assessment of tumor response post-treatment, which may include imaging studies or biomarkers to evaluate changes in tumor size or metabolic activity.  8. **Patient Outcomes**:  - Long-term follow-up to assess the impact on tumor control, survival rates, and quality of life. 9. **Documentation and Protocol Adherence**:  - Ensuring that the session follows established protocols and that all parameters and outcomes are thoroughly documented. By evaluating these metrics, clinicians can assess the quality of a hyperthermia session and make necessary adjustments to optimize treatment outcomes. |
| 35 | Is invasive temperature monitoring during hyperthermia sessions clinically justified? | | |
|  | DeepSeek-V3 | Llama-3.3-70B-Instruct | GPT-4o |
|  | Invasive temperature monitoring during hyperthermia sessions can be clinically justified in certain scenarios, but its necessity depends on the treatment goals, tumor location, and available non-invasive alternatives. Here is a breakdown of key considerations: ### **1. Clinical Justification for Invasive Monitoring** - **Accuracy**: Invasive probes (e.g., fiber-optic or thermocouple sensors) provide **real-time, precise temperature measurements** at specific tissue depths, which is critical for ensuring therapeutic efficacy (typically 40-45°C) while avoiding overheating. - **Deep-Seated Tumors**: For **deep or internally located tumors** (e.g., pelvic, abdominal, or brain malignancies), non-invasive methods (MRI thermometry, microwave radiometry) may lack sufficient resolution, making invasive probes necessary. - **Quality Assurance**: Some clinical protocols (e.g., **CEM43°C thermal dose modeling**) require exact temperature data to ensure tumor cytotoxicity while sparing normal tissue. ### **2. When Non-Invasive Methods May Suffice** - **Superficial Hyperthermia**: For skin or near-surface tumors, **infrared thermography or ultrasound-based thermometry** may be adequate. - **MRI-Guided Hyperthermia**: MR thermometry (e.g., proton resonance frequency shift) offers non-invasive, 3D temperature mapping and is increasingly used in focused ultrasound (HIFU) and RF hyperthermia. ### **3. Risks & Limitations of Invasive Monitoring** - **Procedural Risks**: Insertion may cause bleeding, infection, or discomfort. - **Limited Spatial Coverage**: Single-point measurements may miss temperature heterogeneity. - **Interference with Treatment**: Metallic probes can distort electromagnetic fields in RF or microwave hyperthermia. ### **4. Guidelines & Evidence** - **ESTRO & ESHO Recommendations**: Support invasive monitoring when precise thermal dosing is critical (e.g., **thermoradiotherapy for cervical cancer**). - **Clinical Studies**: Some trials (e.g., **DEEP study for recurrent breast cancer**) have used invasive probes to validate non-invasive thermometry. ### **Conclusion** Invasive monitoring is **justified when**: - High precision is required for deep or critical structures. - Non-invasive methods are unreliable or unavailable. - The clinical protocol demands direct thermal dosimetry. However, **non-invasive alternatives should be preferred** when feasible, balancing accuracy with patient safety. The decision should be individualized based on tumor characteristics, available technology, and institutional expertise. | Invasive temperature monitoring during hyperthermia sessions is a topic of ongoing debate among medical professionals. Hyperthermia, also known as thermal therapy or heat treatment, is a type of cancer treatment that involves exposing cancer cells to high temperatures to damage or kill them. Invasive temperature monitoring involves inserting a probe or catheter into the body to measure the temperature of the tumor or surrounding tissue directly. This can be done using various techniques, such as: 1. Intratumoral temperature probes: These are inserted directly into the tumor to measure its temperature. 2. Interstitial temperature probes: These are inserted into the tissue surrounding the tumor to measure the temperature of the surrounding tissue. 3. Catheter-based temperature monitoring: This involves inserting a catheter into a blood vessel or other body cavity to measure the temperature of the surrounding tissue. The potential benefits of invasive temperature monitoring during hyperthermia sessions include: 1. **Accurate temperature measurement**: Invasive monitoring can provide more accurate temperature measurements than non-invasive methods, such as infrared thermometry or magnetic resonance imaging (MRI). 2. **Real-time feedback**: Invasive monitoring can provide real-time feedback on the temperature of the tumor or surrounding tissue, allowing for adjustments to be made to the hyperthermia treatment in real-time. 3. **Improved treatment efficacy**: By ensuring that the tumor is heated to the optimal temperature, invasive monitoring may help improve the efficacy of hyperthermia treatment. However, invasive temperature monitoring during hyperthermia sessions also has some potential drawbacks and limitations: 1. **Invasiveness**: Inserting a probe or catheter into the body can be invasive and may cause discomfort, pain, or other complications, such as infection or bleeding. 2. **Risk of tissue damage**: Inserting a probe or catheter into the tumor or surrounding tissue can cause tissue damage or disrupt the tumor's blood supply, which may affect the treatment outcome. 3. **Limited availability**: Invasive temperature monitoring may not be widely available or may require specialized equipment and expertise. 4. **Cost**: Invasive temperature monitoring may be more expensive than non-invasive methods, which could be a barrier to its adoption. The clinical justification for invasive temperature monitoring during hyperthermia sessions depends on various factors, including: 1. **Tumor type and location**: Invasive monitoring may be more suitable for certain types of tumors, such as those located in deep tissues or with complex anatomy. 2. **Treatment protocol**: Invasive monitoring may be necessary for certain hyperthermia treatment protocols, such as those that require precise temperature control or real-time feedback. 3. **Patient-specific factors**: Invasive monitoring may be more suitable for patients with certain medical conditions or those who are at high risk of complications from non-invasive monitoring methods. In conclusion, invasive temperature monitoring during hyperthermia sessions can be clinically justified in certain situations, such as when accurate temperature measurement is crucial for treatment efficacy or when non-invasive methods are not feasible. However, the decision to use invasive monitoring should be made on a case-by-case basis, taking into account the potential benefits and drawbacks, as well as the individual patient's needs and circumstances. | Invasive temperature monitoring during hyperthermia sessions can be clinically justified in certain situations, depending on the specific clinical context and the goals of the treatment. Hyperthermia therapy, which involves raising the temperature of body tissues to enhance the effectiveness of treatments like radiation therapy or chemotherapy, requires precise temperature control to ensure efficacy and safety. Here are some reasons why invasive temperature monitoring might be justified: 1. **Accuracy and Precision**: Invasive methods, such as using thermistors or thermocouples inserted into body cavities or tissues, provide more accurate and precise temperature measurements compared to non-invasive methods. This accuracy is crucial for ensuring that the target tissues reach the desired therapeutic temperature without causing damage to surrounding healthy tissues. 2. **Critical Situations**: In cases where the therapeutic window is narrow, and precise temperature control is essential to avoid complications, invasive monitoring can provide the necessary data to adjust treatment parameters in real-time. 3. **Research and Development**: In clinical trials or research settings, where detailed data on tissue response to hyperthermia is needed, invasive monitoring can provide valuable insights that non-invasive methods might not capture. 4. **Patient-Specific Factors**: Some patients may have conditions or anatomical considerations that make non-invasive monitoring less reliable. In such cases, invasive monitoring might be the best option to ensure effective treatment. However, invasive temperature monitoring also comes with risks, such as infection, discomfort, and potential complications from the insertion of probes. Therefore, the decision to use invasive monitoring should be based on a careful assessment of the risks and benefits, considering the specific clinical scenario, the patient's condition, and the available resources. In many cases, non-invasive methods, such as infrared thermography or external probes, may be sufficient and preferred due to their lower risk profile. Ultimately, the choice of monitoring technique should be guided by clinical judgment, patient safety, and the specific requirements of the hyperthermia treatment protocol. |
| 36 | What are the advantages and limitations of capacitive hyperthermia techniques? | | |
|  | DeepSeek-V3 | Llama-3.3-70B-Instruct | GPT-4o |
|  | Capacitive hyperthermia (also known as capacitive-coupled hyperthermia or radiofrequency capacitive hyperthermia) is a non-invasive or minimally invasive technique used in cancer therapy to selectively heat tumor tissues. Below are its key **advantages** and **limitations**: ### **Advantages:** 1. **Selective Heating of Deep Tissues**   - Capacitive hyperthermia can penetrate deeper tissues (up to 10 cm) compared to some other hyperthermia methods, making it useful for treating deep-seated tumors. 2. **Non-Invasive or Minimally Invasive**   - Unlike surgical interventions, capacitive hyperthermia typically uses external electrodes, reducing trauma and recovery time. 3. **Synergistic with Radiotherapy and Chemotherapy**   - Heating tumors can enhance the effectiveness of radiation and certain chemotherapeutic agents by improving oxygenation and drug uptake in cancer cells. 4. **Localized Treatment**   - The heating effect is focused on the tumor region, minimizing damage to surrounding healthy tissues when properly controlled. 5. **Minimal Systemic Side Effects**   - Unlike systemic treatments (e.g., chemotherapy), hyperthermia primarily affects the targeted area, reducing systemic toxicity. 6. **Applicable to Various Tumor Types**   - Effective for solid tumors in organs like the liver, prostate, breast, and soft tissues. ### **Limitations:** 1. **Uneven Heating and Hot Spots**   - Due to differences in tissue conductivity, overheating (hot spots) can occur in fatty or high-resistance tissues, while underheating may happen in highly vascularized regions. 2. **Limited Penetration in Some Cases**   - While deeper than some methods, capacitive hyperthermia may still struggle with very large or highly heterogeneous tumors. 3. **Dependence on Tumor Location and Size**   - Superficial tumors may be easier to treat than deep or irregularly shaped ones. 4. **Patient Discomfort and Pain**   - High-intensity heating can cause pain or burns if not carefully monitored. 5. **Technical Challenges in Temperature Control**   - Maintaining uniform and precise temperature (typically 40-45°C) is difficult, requiring real-time thermometry (e.g., MRI thermometry or invasive probes). 6. **Limited Clinical Adoption**   - While promising, capacitive hyperthermia is not yet a standard treatment in all oncology centers, partly due to variability in outcomes and the need for specialized equipment. 7. **Potential for Skin Burns**   - Since electrodes are placed on the skin, improper application can lead to superficial burns. ### **Conclusion:**  Capacitive hyperthermia is a valuable adjunctive cancer therapy with benefits in enhancing conventional treatments, but its effectiveness depends on precise control of heating parameters and tumor characteristics. Ongoing advancements in imaging and temperature monitoring may help overcome current limitations. | Capacitive hyperthermia is a type of hyperthermia treatment that uses electromagnetic fields to heat cancer cells, causing them to die or become more susceptible to other treatments. The advantages and limitations of capacitive hyperthermia techniques are: **Advantages:** 1. **Non-invasive**: Capacitive hyperthermia is a non-invasive treatment, which means it does not require surgery or insertion of probes into the body. 2. **Localized heating**: The technique allows for localized heating of the tumor, minimizing damage to surrounding healthy tissues. 3. **Deep penetration**: Capacitive hyperthermia can penetrate deep into the body, making it suitable for treating tumors located in internal organs. 4. **Flexibility**: The technique can be used to treat a variety of cancer types, including breast, prostate, and liver cancer. 5. **Low toxicity**: Capacitive hyperthermia is generally well-tolerated, with minimal side effects compared to other cancer treatments. 6. **Combination therapy**: Capacitive hyperthermia can be combined with other cancer treatments, such as chemotherapy and radiation therapy, to enhance their effectiveness. **Limitations:** 1. **Limited depth of penetration**: While capacitive hyperthermia can penetrate deep into the body, its effectiveness may decrease with increasing depth, making it less suitable for very deep-seated tumors. 2. **Heterogeneous heating**: The heating pattern can be heterogeneous, leading to uneven temperature distributions within the tumor, which may affect treatment outcomes. 3. **Tumor size and location**: The technique may be less effective for large tumors or those located in areas with limited access, such as the brain or spinal cord. 4. **Blood flow and cooling**: Blood flow can cool the tumor, reducing the effectiveness of the treatment, and may require additional measures to mitigate this effect. 5. **Technical challenges**: Capacitive hyperthermia requires sophisticated equipment and expertise to ensure accurate and safe treatment delivery. 6. **Limited clinical data**: While capacitive hyperthermia has shown promise in preclinical and clinical studies, more research is needed to fully establish its efficacy and optimal treatment protocols. 7. **Cost and availability**: Capacitive hyperthermia equipment and treatment may not be widely available or affordable, limiting access to this treatment option. 8. **Potential side effects**: While generally well-tolerated, capacitive hyperthermia can cause side effects such as pain, skin burns, or nerve damage, particularly if not properly monitored and managed. Overall, capacitive hyperthermia is a promising treatment option for certain types of cancer, offering advantages such as non-invasiveness and localized heating. However, its limitations, including technical challenges and limited clinical data, must be carefully considered and addressed to ensure effective and safe treatment delivery. | Capacitive hyperthermia is a technique used in medical treatments, particularly in oncology, to raise the temperature of body tissues to enhance the effects of other treatments like radiation or chemotherapy. Here are some of the advantages and limitations of capacitive hyperthermia: ### Advantages: 1. **Selective Heating:**  - Capacitive hyperthermia can selectively target tumor tissues due to their different electrical properties compared to normal tissues, potentially minimizing damage to healthy cells. 2. **Enhanced Treatment Efficacy:**  - When combined with radiation or chemotherapy, hyperthermia can increase the sensitivity of cancer cells to these treatments, potentially improving their effectiveness. 3. **Non-Invasive:**  - The technique is generally non-invasive, which reduces the risk of infection and other complications associated with surgical procedures. 4. **Pain Management:**  - Hyperthermia can sometimes help in reducing pain associated with tumors, providing symptomatic relief to patients. 5. **Improved Blood Flow:**  - The heat can increase blood flow to the treated area, which may enhance the delivery of chemotherapeutic agents to the tumor. ### Limitations: 1. **Depth of Penetration:**  - Capacitive hyperthermia may have limitations in heating deep-seated tumors effectively, as the energy may not penetrate deeply enough to reach all tumor cells. 2. **Temperature Control:**  - Maintaining the precise temperature required for effective treatment without damaging surrounding healthy tissues can be challenging. 3. **Equipment and Expertise:**  - The technique requires specialized equipment and trained personnel, which may not be available in all treatment centers. 4. **Side Effects:**  - Patients may experience side effects such as burns, blisters, or discomfort in the treated area due to the heat. 5. **Limited Standalone Efficacy:**  - Hyperthermia is often not effective as a standalone treatment and is typically used in conjunction with other therapies. 6. **Patient Variability:**  - The effectiveness of capacitive hyperthermia can vary significantly between patients, depending on factors like tumor type, location, and individual patient characteristics. Overall, while capacitive hyperthermia offers several benefits, particularly as an adjunct to other cancer treatments, its limitations must be carefully considered, and it should be applied in appropriate clinical contexts. |
| 37 | What are the advantages and limitations of radiative hyperthermia techniques? | | |
|  | DeepSeek-V3 | Llama-3.3-70B-Instruct | GPT-4o |
|  | Radiative hyperthermia techniques, which use electromagnetic radiation (e.g., microwaves, radiofrequency, or infrared) to heat tissues for therapeutic purposes, offer several advantages and limitations in medical applications, particularly in cancer treatment. ### **Advantages of Radiative Hyperthermia:** 1. **Non-Invasive or Minimally Invasive:**   - Many radiative hyperthermia methods (e.g., microwave ablation, radiofrequency ablation) are minimally invasive, reducing surgical risks.   - Some techniques (e.g., focused ultrasound) are entirely non-invasive. 2. **Localized Treatment:**   - Can selectively target tumors while sparing surrounding healthy tissue, especially when combined with imaging guidance (MRI, ultrasound).  3. **Synergistic with Other Therapies:**   - Enhances the effectiveness of chemotherapy and radiotherapy by increasing tumor blood flow and oxygen levels, making cancer cells more susceptible to treatment.  4. **Controlled Heating:**   - Advanced systems allow precise temperature control (typically 40-45°C for hyperthermia, higher for ablation), minimizing damage to normal tissues.  5. **Repeatable and Adjustable:**   - Can be applied in multiple sessions if needed, unlike some irreversible surgical procedures.  6. **Potential for Immune Stimulation:**   - Hyperthermia may trigger immune responses against cancer cells, aiding in systemic anti-tumor effects.  ### **Limitations of Radiative Hyperthermia:** 1. **Limited Penetration Depth:**   - High-frequency waves (e.g., microwaves) have shallow tissue penetration, making deep-seated tumors harder to treat effectively.  2. **Non-Uniform Heating:**   - Heterogeneous tissue properties (e.g., fat, muscle, bone) can cause uneven heat distribution, leading to under- or over-treated areas.  3. **Thermal Damage Risks:**   - Overheating can cause burns, necrosis, or damage to adjacent critical structures (e.g., nerves, blood vessels).  4. **Technical Complexity:**   - Requires precise real-time temperature monitoring (e.g., via MRI thermometry) and skilled operators to avoid complications.  5. **Limited Efficacy in Certain Cancers:**   - Some tumors (e.g., highly vascularized or heat-resistant cancers) may not respond well to hyperthermia alone.  6. **Cost and Accessibility:**   - Advanced hyperthermia systems (e.g., magnetic nanoparticle hyperthermia, focused ultrasound) can be expensive and require specialized facilities.  ### **Conclusion:**  Radiative hyperthermia is a promising adjunctive cancer therapy with benefits like localized treatment and synergy with other modalities. However, challenges such as penetration depth, heating uniformity, and technical demands must be addressed for broader clinical adoption. Ongoing advancements in imaging and heating technologies aim to overcome these limitations.  Would you like details on specific hyperthermia techniques (e.g., RF ablation, microwave hyperthermia, magnetic hyperthermia)? | Radiative hyperthermia techniques, which involve the use of electromagnetic radiation (such as microwaves, radiofrequencies, or infrared light) to heat tissues, have several advantages and limitations. Here are some of the key benefits and drawbacks: **Advantages:** 1. **Non-invasive**: Radiative hyperthermia is a non-invasive treatment, which means that it does not require surgical incisions or insertion of probes into the body. 2. **Deep penetration**: Electromagnetic radiation can penetrate deep into tissues, allowing for the treatment of tumors located at various depths. 3. **Selective heating**: Radiative hyperthermia can be designed to selectively heat cancer cells, minimizing damage to surrounding healthy tissues. 4. **Flexibility**: Radiative hyperthermia can be used to treat a wide range of tumor types and locations, including superficial and deep-seated tumors. 5. **Combination with other therapies**: Radiative hyperthermia can be combined with other cancer treatments, such as chemotherapy, radiation therapy, or immunotherapy, to enhance their effectiveness. 6. **Minimally toxic**: Radiative hyperthermia is generally considered to be a minimally toxic treatment, with few side effects compared to other cancer therapies. **Limitations:** 1. **Heterogeneous heating**: The heating pattern produced by radiative hyperthermia can be heterogeneous, leading to uneven temperature distributions within the tumor. 2. **Limited control**: It can be challenging to control the temperature and heating pattern in real-time, which may lead to overheating or underheating of the tumor. 3. **Tissue variability**: The electrical and thermal properties of tissues can vary significantly, making it difficult to predict the heating pattern and temperature distribution. 4. **Depth limitations**: While radiative hyperthermia can penetrate deep into tissues, its effectiveness may decrease with increasing depth due to attenuation of the electromagnetic radiation. 5. **Scalability**: Radiative hyperthermia may not be suitable for large tumors or tumors with complex geometries, as it can be difficult to achieve uniform heating. 6. **Cost and accessibility**: Radiative hyperthermia equipment can be expensive, and access to these treatments may be limited in some regions or countries. 7. **Lack of standardization**: There is currently a lack of standardization in radiative hyperthermia protocols, which can make it challenging to compare results across different studies and institutions. 8. **Potential side effects**: While generally considered safe, radiative hyperthermia can cause side effects such as skin burns, pain, or discomfort, particularly if the treatment is not carefully monitored and controlled. Overall, radiative hyperthermia techniques offer a promising approach for cancer treatment, but their effectiveness and safety depend on careful treatment planning, monitoring, and control. Ongoing research aims to address the limitations and improve the efficacy of these techniques. | Radiative hyperthermia is a medical treatment technique that involves raising the temperature of body tissues to damage and kill cancer cells or make them more susceptible to the effects of radiation and chemotherapy. This technique uses electromagnetic radiation, such as microwaves or radiofrequency waves, to generate heat. Here are some of the advantages and limitations of radiative hyperthermia: ### Advantages: 1. **Targeted Treatment:**  - Radiative hyperthermia can be precisely targeted to specific areas, minimizing damage to surrounding healthy tissues. 2. **Enhanced Efficacy of Other Treatments:**  - It can enhance the effectiveness of other cancer treatments, such as radiation therapy and chemotherapy, by making cancer cells more sensitive to these treatments. 3. **Non-Invasive:**  - As a non-invasive technique, it reduces the risks and recovery time associated with surgical procedures. 4. **Minimal Side Effects:**  - Compared to other cancer treatments, radiative hyperthermia generally has fewer and less severe side effects. 5. **Improved Blood Flow:**  - The heat can increase blood flow to the tumor area, potentially improving the delivery of chemotherapy drugs. 6. **Potential for Repeated Use:**  - It can often be repeated multiple times, which is beneficial for managing recurrent tumors. ### Limitations: 1. **Limited Penetration Depth:**  - The effectiveness of radiative hyperthermia can be limited by the depth of penetration of the electromagnetic waves, making it less effective for deep-seated tumors. 2. **Technical Challenges:**  - Precise control of temperature and accurate targeting can be technically challenging, requiring specialized equipment and expertise. 3. **Variable Response:**  - Not all tumors respond equally to hyperthermia, and the response can vary depending on the type and location of the tumor. 4. **Potential for Overheating:**  - There is a risk of overheating, which can damage healthy tissues if not carefully monitored and controlled. 5. **Limited Availability:**  - Access to radiative hyperthermia may be limited due to the need for specialized equipment and trained personnel, which may not be available in all treatment centers. 6. **Cost:**  - The cost of the equipment and treatment can be high, potentially limiting its availability to patients. Overall, while radiative hyperthermia offers several advantages, particularly as an adjunct to other cancer treatments, its limitations must be carefully considered in the context of individual patient needs and treatment goals. |
| 38 | What is the most accurate method to calculate the expected temperature distribution during a capacitive hyperthermia treatment? | | |
|  | DeepSeek-V3 | Llama-3.3-70B-Instruct | GPT-4o |
|  | The most accurate method to calculate the expected temperature distribution during **capacitive hyperthermia treatment** involves a combination of **bioheat modeling, numerical simulations, and patient-specific parameterization**. Here is a structured approach: ### **1. Pennes Bioheat Equation (PBE)** The **Pennes Bioheat Transfer Equation** is the most widely used model for hyperthermia simulations: \[ \rho c \frac{\partial T}{\partial t} = \nabla \cdot (k \nabla T) + \rho_b c_b \omega_b (T_b - T) + Q_{met} + Q_{ext} \] - **\(T\)**: Tissue temperature  - **\(\rho, c, k\)**: Density, specific heat, and thermal conductivity of tissue  - **\(\rho_b, c_b, \omega_b, T_b\)**: Blood density, specific heat, perfusion rate, and arterial temperature  - **\(Q_{met}\)**: Metabolic heat generation (often negligible in hyperthermia)  - **\(Q_{ext}\)**: External heat deposition from capacitive RF electrodes  ### **2. Electromagnetic (EM) Modeling for \(Q_{ext}\)** Since capacitive hyperthermia uses **radiofrequency (RF) electrodes**, the **specific absorption rate (SAR)** must be computed: \[ Q_{ext} = \sigma \|E\|^2 \] - **\(\sigma\)**: Tissue electrical conductivity  - **\(E\)**: Electric field strength (from Maxwell is equations)  A **Finite Element Method (FEM)** or **Finite Difference Time Domain (FDTD)** simulation can solve the EM field distribution in heterogeneous tissues. ### **3. Patient-Specific Parameters** - **Dielectric & Thermal Properties**: Use **MRI/CT-based segmentation** to assign tissue-specific \(\sigma, k, \rho, c, \omega_b\).  - **Blood Perfusion (\(\omega_b\))**: Can be estimated from **perfusion imaging (DCE-MRI, PET)** or literature values.  - **Electrode Configuration**: Model the **geometry, frequency, and power settings** of the capacitive applicator. ### **4. Numerical Solution Methods** - **Finite Element Analysis (FEA)**: COMSOL, ANSYS, or custom solvers for coupled EM-thermal simulations.  - **Finite Difference/Volume Methods**: For simpler geometries.  - **Monte Carlo Methods**: For probabilistic variations in tissue properties.  ### **5. Validation & Calibration** - **Phantom Experiments**: Compare simulations with **gel phantom measurements** under controlled RF heating.  - **Clinical Thermometry**: Use **fiber-optic probes or MR thermometry** to validate in vivo.  ### **6. Advanced Considerations** - **Dynamic Perfusion Changes**: Some models incorporate **temperature-dependent perfusion** (e.g., Arrhenius-based damage models).  - **3D Transient Simulations**: Time-dependent analysis for treatment optimization.  ### **Conclusion** The **most accurate method** combines: 1. **Pennes Bioheat Equation** with **patient-specific EM and thermal properties**.  2. **High-resolution FEM/FDTD simulations** for SAR and temperature distribution.  3. **Experimental validation** using phantoms or clinical data.  For clinical applications, **real-time MR thermometry** can further refine predictions during treatment. Would you like details on a specific aspect (e.g., electrode modeling, perfusion effects)? | Calculating the expected temperature distribution during a capacitive hyperthermia treatment is a complex task that involves solving the bioheat equation, which describes the heat transfer within biological tissues. The most accurate method to calculate the expected temperature distribution would involve a combination of numerical modeling and experimental validation. Here's a step-by-step approach: 1. **Finite Element Method (FEM) modeling**: Use a FEM software (e.g., COMSOL, ANSYS) to create a 3D model of the tumor and surrounding tissues. This model should include the electrical properties (e.g., conductivity, permittivity) and thermal properties (e.g., specific heat, thermal conductivity) of the tissues. 2. **Electromagnetic simulation**: Simulate the electromagnetic field distribution within the tissue using the FEM model. This will provide the specific absorption rate (SAR) distribution, which represents the amount of electromagnetic energy absorbed by the tissue. 3. **Bioheat equation**: Solve the bioheat equation, which describes the heat transfer within the tissue. The bioheat equation takes into account the SAR distribution, blood perfusion, and metabolic heat generation. The equation is typically solved using a numerical method, such as the finite difference method or the finite element method. 4. **Thermal properties**: Use accurate thermal properties for the tissues, including specific heat, thermal conductivity, and blood perfusion rates. These properties can be found in the literature or measured experimentally. 5. **Boundary conditions**: Apply realistic boundary conditions to the model, including the temperature of the surrounding tissues, the cooling effect of blood flow, and the heat loss to the environment. 6. **Validation**: Validate the model by comparing the simulated temperature distribution with experimental measurements or clinical data. 7. **Iterative refinement**: Refine the model by adjusting the input parameters and boundary conditions to improve the accuracy of the simulated temperature distribution. Some additional considerations to improve the accuracy of the calculation: * **Tissue heterogeneity**: Account for the heterogeneity of tissue properties, such as the variation in electrical and thermal properties between different tissue types. * **Blood flow**: Include the effects of blood flow on heat transfer, such as the cooling effect of blood flow and the heat transfer between blood vessels and surrounding tissues. * **Temperature-dependent properties**: Use temperature-dependent properties for the tissues, such as the variation in electrical conductivity and thermal conductivity with temperature. * **Non-uniform heating**: Account for non-uniform heating patterns, such as the formation of hot spots or cold spots, which can affect the temperature distribution. By following this approach and considering the complexities of heat transfer in biological tissues, you can develop an accurate model of the expected temperature distribution during a capacitive hyperthermia treatment. | Calculating the expected temperature distribution during capacitive hyperthermia treatment involves complex modeling due to the interaction of electromagnetic fields with biological tissues. The most accurate methods typically involve numerical simulations that account for the specific properties of the tissues, the geometry of the treatment area, and the parameters of the hyperthermia device. Here are some key approaches: 1. **Finite Element Method (FEM):** This is one of the most widely used numerical techniques for simulating temperature distribution. FEM allows for detailed modeling of complex geometries and heterogeneous tissue properties. It solves the bioheat transfer equation, often the Pennes' bioheat equation, to predict temperature changes. 2. **Finite Difference Time Domain (FDTD):** This method is used to solve Maxwell's equations for electromagnetic fields and can be coupled with thermal models to predict temperature distribution. It is particularly useful for simulating the interaction of electromagnetic fields with tissues. 3. **Computational Fluid Dynamics (CFD):** While more commonly used for fluid flow, CFD can be adapted to model heat transfer in tissues, especially when considering blood flow and its cooling effect. 4. **Monte Carlo Simulations:** These are used less frequently for direct temperature calculations but can be useful for modeling the stochastic nature of tissue properties and their impact on heat distribution. 5. **Hybrid Models:** Combining different methods, such as FEM for thermal modeling and FDTD for electromagnetic field simulation, can provide more comprehensive and accurate results. For the most accurate predictions, these models require detailed input data, including tissue-specific thermal and electrical properties, accurate anatomical models (often derived from medical imaging), and precise parameters of the hyperthermia device. Validation with experimental or clinical data is also crucial to ensure the reliability of the simulations. |
| 39 | What is the most accurate method to calculate the expected temperature distribution during a radiative hyperthermia treatment? | | |
|  | DeepSeek-V3 | Llama-3.3-70B-Instruct | GPT-4o |
|  | The most accurate method to calculate the expected temperature distribution during **radiative hyperthermia treatment** involves a combination of **bioheat transfer modeling, computational simulations, and patient-specific data integration**. Here is a step-by-step breakdown of the key components: ### **1. Pennes Bioheat Transfer Equation (BHTE)** The **Pennes equation** is the most widely used model for predicting temperature distribution in biological tissues during hyperthermia: \[ \rho c \frac{\partial T}{\partial t} = \nabla \cdot (k \nabla T) + \omega_b \rho_b c_b (T_b - T) + Q_{met} + Q_{ext} \] Where: - \(T\) = Tissue temperature (°C) - \(\rho\) = Tissue density (kg/m³) - \(c\) = Tissue specific heat (J/kg·K) - \(k\) = Thermal conductivity (W/m·K) - \(\omega_b\) = Blood perfusion rate (kg/m³·s) - \(\rho_b, c_b\) = Blood density and specific heat - \(T_b\) = Arterial blood temperature (~37°C) - \(Q_{met}\) = Metabolic heat generation (W/m³) - \(Q_{ext}\) = External heat source (e.g., RF, microwave, laser) ### **2. Incorporating Electromagnetic (EM) Heating** For radiative hyperthermia (e.g., RF, microwave, or laser heating), the **specific absorption rate (SAR)** must be calculated: \[ SAR = \frac{\sigma \|E\|^2}{2\rho} \] Where: - \(\sigma\) = Tissue electrical conductivity (S/m) - \(E\) = Electric field (V/m) - \(\rho\) = Tissue density (kg/m³) The **external heat source term** \(Q_{ext}\) is then: \[ Q_{ext} = SAR \cdot \rho \] ### **3. Numerical Methods for Solving the Model** - **Finite Element Method (FEM)** or **Finite Difference Time Domain (FDTD)** simulations are used to solve the coupled EM and thermal equations. - Commercial software (e.g., COMSOL Multiphysics, ANSYS) or custom solvers can be employed. ### **4. Patient-Specific Parameters for Accuracy** - **Tissue properties** (thermal/electrical conductivity, perfusion) vary with tissue type (e.g., muscle, fat, tumor). - **Medical imaging (MRI, CT, ultrasound)** helps define anatomical geometry and tissue boundaries. - **Real-time thermometry** (e.g., MR thermometry, fiber-optic probes) can refine simulations. ### **5. Validation & Calibration** - Compare simulated results with **experimental measurements** (phantom studies or clinical data). - Adjust model parameters (e.g., perfusion rates) to match observed temperature distributions. ### **6. Advanced Considerations** - **Dynamic perfusion changes** (blood flow increases with temperature). - **Nonlinear thermal properties** (tissue damage thresholds, Arrhenius models). - **Multi-physics coupling** (fluid dynamics for large vessels). ### **Conclusion** The most accurate method combines: 1. **Pennes BHTE** with **EM heating (SAR)**. 2. **Patient-specific imaging data** for geometry and properties. 3. **High-fidelity numerical simulations (FEM/FDTD)**. 4. **Experimental validation** for calibration. Would you like recommendations on specific software tools or clinical implementation strategies? | Calculating the expected temperature distribution during a radiative hyperthermia treatment is a complex task that involves solving the bioheat transfer equation, which describes the heat transfer mechanisms in biological tissues. The most accurate method to calculate the expected temperature distribution is through numerical simulations using computational models, specifically: 1. **Finite Element Method (FEM)**: This method is widely used to solve the bioheat transfer equation in complex geometries, such as the human body. FEM discretizes the computational domain into smaller elements, allowing for the calculation of temperature distributions at each node. 2. **Finite Difference Method (FDM)**: Similar to FEM, FDM discretizes the computational domain into a grid, and the bioheat transfer equation is solved at each grid point. 3. **Monte Carlo Method**: This method uses statistical simulations to model the behavior of photons and heat transfer in tissues. It is particularly useful for modeling complex radiation interactions, such as scattering and absorption. 4. **Computational Fluid Dynamics (CFD)**: CFD can be used to model blood flow and heat transfer in tissues, which is essential for accurate temperature distribution calculations. To increase the accuracy of these simulations, it's essential to consider the following factors: 1. **Tissue properties**: Accurate values of thermal conductivity, specific heat capacity, and density for each tissue type are crucial. 2. **Blood flow**: Modeling blood flow and its effects on heat transfer is essential, as it can significantly impact temperature distributions. 3. **Radiation interactions**: Accurate modeling of radiation interactions, such as absorption, scattering, and reflection, is necessary to predict the temperature distribution. 4. **Boundary conditions**: Appropriate boundary conditions, such as heat transfer coefficients and temperature values, must be applied to the computational model. 5. **Validation**: Experimental validation of the computational model is essential to ensure its accuracy and reliability. Some popular software packages for simulating radiative hyperthermia treatments include: 1. **COMSOL Multiphysics**: A commercial software package that uses FEM to solve various physics problems, including bioheat transfer. 2. **ANSYS**: A commercial software package that offers a range of simulation tools, including FEM and CFD, for bioheat transfer and fluid dynamics. 3. **OpenFOAM**: An open-source software package that uses FVM (Finite Volume Method) to solve various physics problems, including bioheat transfer. 4. **MATLAB**: A high-level programming language and software package that can be used to implement custom simulations using FEM, FDM, or other numerical methods. By using these computational models and considering the factors mentioned above, researchers and clinicians can accurately predict the temperature distribution during radiative hyperthermia treatments, which is essential for optimizing treatment planning and ensuring patient safety. | Calculating the expected temperature distribution during a radiative hyperthermia treatment involves complex modeling due to the interaction of electromagnetic fields with biological tissues. The most accurate methods typically involve a combination of computational techniques and empirical data. Here are some key approaches: 1. **Finite Element Method (FEM):** This numerical technique is widely used for solving complex physical problems, including heat transfer in biological tissues. FEM can model the spatial distribution of temperature by solving the bioheat transfer equation, which accounts for heat conduction, perfusion, and metabolic heat generation. 2. **Bioheat Transfer Equation:** The Pennes' bioheat equation is commonly used to model heat transfer in tissues. It considers the effects of blood perfusion, metabolic heat production, and thermal conduction. Modifications to this equation can include more detailed models of perfusion and anisotropic thermal properties. 3. **Electromagnetic Field Simulation:** Accurate modeling of the electromagnetic field distribution is crucial, as it determines the initial energy deposition in tissues. Techniques like the Finite-Difference Time-Domain (FDTD) method or the Finite Element Method (FEM) can be used to simulate the electromagnetic field distribution. 4. **Thermal Dose Calculation:** The Arrhenius model or CEM43 (Cumulative Equivalent Minutes at 43°C) can be used to predict thermal damage and ensure that the treatment is within therapeutic limits. 5. **Patient-Specific Modeling:** Incorporating patient-specific anatomical data from imaging modalities like MRI or CT scans can improve the accuracy of the model. This allows for personalized treatment planning and more precise predictions of temperature distribution. 6. **Validation with Experimental Data:** Combining computational models with experimental data from phantom studies or clinical measurements can help validate and refine the models, improving their accuracy. 7. **Multiphysics Simulation Software:** Tools like COMSOL Multiphysics or ANSYS can integrate electromagnetic and thermal simulations, providing a comprehensive platform for modeling hyperthermia treatments. By combining these methods, researchers and clinicians can achieve a more accurate prediction of temperature distribution during radiative hyperthermia treatments, leading to more effective and safer therapeutic outcomes. |
| 40 | Design a perfect hyperthermia device capable to perform superficial, deep and capacitive hyperthermia treatment. | | |
|  | DeepSeek-V3 | Llama-3.3-70B-Instruct | GPT-4o |
|  | Designing a **perfect hyperthermia device** capable of performing **superficial, deep, and capacitive hyperthermia** requires a multi-modal approach that integrates different heating mechanisms while ensuring safety, precision, and adaptability. Below is a comprehensive design concept: ### **Multi-Modal Hyperthermia Device: "ThermoFlex Pro"**  *(Superficial, Deep, and Capacitive Hyperthermia Capabilities)*  #### **1. Core Heating Technologies**  To cover all treatment depths, the device combines:  - **Radiofrequency (RF) Heating** (for deep tissue)  - **Microwave (MW) Heating** (for superficial/intermediate)  - **Capacitive Coupling (CC) Heating** (for focused, non-invasive energy transfer)  #### **2. Key Components & Features**  ##### **A. Multi-Frequency RF/MW Generator**  - **Frequency Range:**   - **RF (100 kHz - 1 MHz)** for deep tissue penetration (up to 8-10 cm).   - **MW (433 MHz - 2.45 GHz)** for superficial/intermediate heating (1-4 cm).  - **Adjustable Power (10W - 200W)** for controlled thermal dose.  - **Automatic impedance matching** for efficient energy transfer.  ##### **B. Capacitive Hyperthermia System**  - **Dual-electrode capacitive coupling** for deep, non-radiative heating.  - **Dielectric pads** to enhance energy focus and reduce skin burns.  - **Real-time current monitoring** to prevent overheating.  ##### **C. Adaptive Applicator Array**  - **Interchangeable applicators** for different treatment depths:   - **Superficial:** Flexible microwave patches (for skin/subcutaneous).   - **Deep:** RF phased-array applicators (for tumors in muscle/organs).   - **Capacitive:** Conductive plates with cooling gel interface.  - **3D scanning & auto-positioning** for optimal energy focus.  ##### **D. Real-Time Temperature Monitoring & Feedback**  - **Multi-point thermocouples & infrared sensors** for surface temp.  - **MRI/Ultrasound thermometry integration** for deep tissue monitoring.  - **AI-driven thermal modeling** to adjust power in real-time.  ##### **E. Safety & Control System**  - **Automatic shutoff** if overheating detected.  - **Patient-specific treatment planning** (based on tumor size/location).  - **EMI shielding** to prevent interference with other devices.  #### **3. Treatment Modes**  - **Superficial Mode:** Microwave applicators for skin cancers, wound healing.  - **Deep Mode:** RF phased-array for prostate, liver, or breast tumors.  - **Capacitive Mode:** Non-invasive deep heating for musculoskeletal pain.  #### **4. User Interface & Software**  - **Touchscreen console** with preset protocols.  - **AI-assisted thermal dose optimization.**  - **Cloud-based patient data logging.**  #### **5. Advantages Over Existing Systems**  œ… **All-in-one solution** (no need for separate devices).  œ… **Precision heating** with real-time feedback.  œ… **Minimal side effects** (cooling, impedance control).  œ… **Adaptable to various clinical needs** (oncology, physiotherapy).  ### **Conclusion**  The **ThermoFlex Pro** would be a **breakthrough hyperthermia device**, combining **RF, microwave, and capacitive heating** in a single, intelligent system. By integrating **real-time thermometry, adaptive applicators, and AI-driven control**, it ensures **safe, effective, and personalized hyperthermia therapy** for both superficial and deep-seated conditions.  Would you like additional details on any specific component? | What a fascinating task! Here's a design for a perfect hyperthermia device capable of performing superficial, deep, and capacitive hyperthermia treatment: **Device Name:** Hyperthermia Triplex (HTX) **Overview:** The Hyperthermia Triplex (HTX) is a non-invasive, multi-modal hyperthermia device designed to provide superficial, deep, and capacitive hyperthermia treatment for various cancer types and other medical conditions. The device combines advanced technologies, including radiofrequency (RF), microwave, and capacitive heating, to deliver precise and controlled heat to tumors or affected areas. **Components:** 1. **Superficial Hyperthermia Module (SHM):** * Frequency: 13.56 MHz (ISM band) * Power output: 0-100 W * Applicator: Flexible, water-cooled, and conformal to the treatment area * Temperature control: ±0.1°C accuracy 2. **Deep Hyperthermia Module (DHM):** * Frequency: 100-200 MHz (RF range) * Power output: 0-500 W * Applicator: Phased array, with 4-8 elements, for focused energy delivery * Temperature control: ±0.1°C accuracy 3. **Capacitive Hyperthermia Module (CHM):** * Frequency: 8-27.12 MHz (capacitive range) * Power output: 0-200 W * Applicator: Capacitive electrodes, with adjustable spacing and size * Temperature control: ±0.1°C accuracy 4. **Control Unit:** * Touchscreen interface for easy operation and treatment planning * Real-time temperature monitoring and control * Integrated safety features, including automatic shutdown and alarm systems 5. **Cooling System:** * Chilled water circulation for applicator cooling * Air-cooled heat exchanger for efficient heat dissipation 6. **Patient Interface:** * Comfortable, adjustable treatment bed with built-in temperature sensors * Integrated patient monitoring system (e.g., ECG, blood pressure, oxygen saturation) **Operating Modes:** 1. **Superficial Hyperthermia:** SHM module is used for treating superficial tumors (e.g., skin, breast, and head and neck cancers). 2. **Deep Hyperthermia:** DHM module is used for treating deeper-seated tumors (e.g., prostate, liver, and brain cancers). 3. **Capacitive Hyperthermia:** CHM module is used for treating tumors with capacitive coupling (e.g., bone, lung, and pancreatic cancers). 4. **Combination Therapy:** HTX can be used in combination with other cancer treatments, such as radiation therapy, chemotherapy, or immunotherapy. **Advanced Features:** 1. **Real-time Temperature Monitoring:** Invasive and non-invasive temperature sensors provide accurate temperature feedback during treatment. 2. **Treatment Planning Software:** Integrated software for treatment planning, simulation, and optimization. 3. **Automated Treatment Protocol:** Pre-programmed treatment protocols for various cancer types and treatment sites. 4. **Data Analysis and Storage:** Comprehensive data analysis and storage for treatment outcomes, patient information, and device performance. **Safety Features:** 1. **Automatic Shutdown:** Device shutdown in case of overheating, power failure, or patient movement. 2. **Alarm Systems:** Audible and visual alarms for temperature deviations, patient discomfort, or system malfunctions. 3. **Electromagnetic Compatibility (EMC):** HTX is designed to meet international EMC standards for safe operation in a clinical environment. 4. **Patient Safety Interlocks:** Multiple safety interlocks to prevent accidental exposure to RF or microwave energy. **Clinical Benefits:** 1. **Improved Treatment Outcomes:** HTX provides precise and controlled heat delivery, enhancing treatment efficacy and reducing side effects. 2. **Increased Patient Comfort:** Comfortable treatment bed, adjustable applicators, and real-time temperature monitoring ensure a pleasant treatment experience. 3. **Flexibility and Versatility:** HTX can be used for various cancer types, treatment sites, and combination therapies, making it a valuable asset for oncology departments. 4. **Cost-Effective:** Reduced treatment times, improved patient outcomes, and minimized side effects contribute to cost savings and improved resource allocation. The Hyperthermia Triplex (HTX) is a cutting-edge, multi-modal hyperthermia device designed to provide superficial, deep, and capacitive hyperthermia treatment for various cancer types and medical conditions. Its advanced features, safety measures, and clinical benefits make it an ideal solution for oncology departments seeking to improve treatment outcomes and patient care. | Designing a perfect hyperthermia device for superficial, deep, and capacitive hyperthermia treatment involves several key considerations to ensure safety, efficacy, and versatility. Below is a conceptual design outline for such a device: ### Key Components and Features 1. **Energy Source:**  - **Microwave and Radiofrequency (RF) Generators:** These are commonly used for hyperthermia treatments. The device should include both microwave and RF capabilities to target different tissue depths effectively.  - **Adjustable Frequency and Power Settings:** Allow customization of energy delivery based on treatment depth and patient-specific needs. 2. **Applicators:**  - **Superficial Applicators:** Designed for treating surface-level tissues. These should be lightweight, ergonomic, and capable of delivering uniform energy distribution.  - **Deep Tissue Applicators:** These should be capable of penetrating deeper tissues, possibly using phased array technology to focus energy precisely.  - **Capacitive Coupling Applicators:** Utilize capacitive plates to create an electric field that heats tissues. These should be adjustable to conform to different body parts. 3. **Temperature Monitoring and Control:**  - **Real-time Temperature Sensors:** Embedded in applicators to provide continuous feedback and ensure precise temperature control.  - **Thermal Imaging:** Integrate infrared cameras or other imaging technologies to visualize temperature distribution in real-time.  - **Automated Feedback System:** Adjusts power output based on temperature readings to maintain optimal therapeutic temperatures (typically 40-45°C). 4. **User Interface:**  - **Touchscreen Display:** Intuitive interface for setting treatment parameters, monitoring progress, and accessing patient data.  - **Pre-set Protocols:** Include standard treatment protocols for common conditions, with the ability to customize for individual patients. 5. **Safety Features:**  - **Automatic Shut-off:** In case of overheating or device malfunction.  - **Patient Monitoring:** Integrate heart rate and oxygen saturation monitoring to ensure patient safety during treatment.  - **Emergency Stop Button:** Easily accessible for both operator and patient. 6. **Portability and Ergonomics:**  - **Compact and Mobile Design:** Ensure the device is easy to move and set up in different clinical settings.  - **Ergonomic Handles and Controls:** For ease of use by healthcare providers. 7. **Data Management:**  - **Patient Data Storage:** Securely store treatment data for future reference and analysis.  - **Connectivity:** Enable data transfer to electronic health records (EHR) systems and remote monitoring capabilities. 8. **Regulatory Compliance:**  - Ensure the device meets all relevant medical device regulations and standards, such as FDA approval in the United States or CE marking in Europe. ### Additional Considerations - **Research and Development:** Collaborate with medical professionals and researchers to continuously improve device efficacy and safety. - **Training and Support:** Provide comprehensive training for healthcare providers and ongoing technical support. - **Cost-effectiveness:** Design with cost in mind to make the device accessible to a wide range of healthcare facilities. By integrating these features, the hyperthermia device can offer versatile and effective treatment options for various medical conditions, enhancing patient outcomes and expanding the capabilities of healthcare providers. |

|  | Likert Scale rating | | |
| --- | --- | --- | --- |
|  | DeepSeek-V3 | Llama-3.3-70B-Instruct | GPT-4o |
|  | 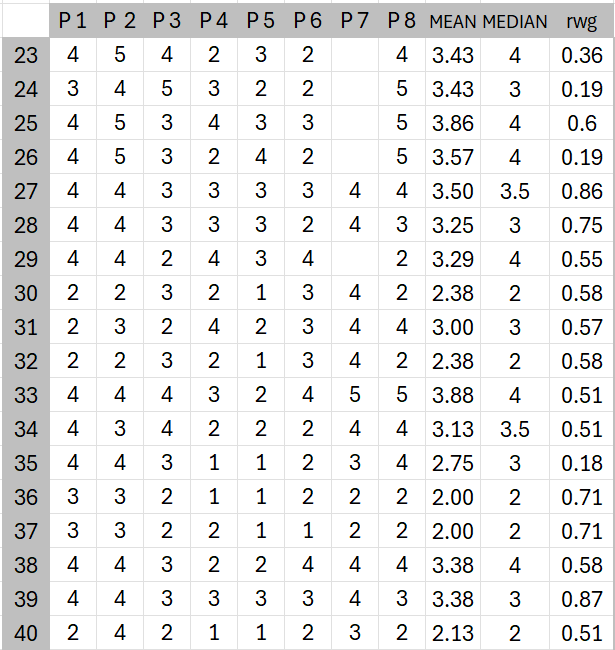 | 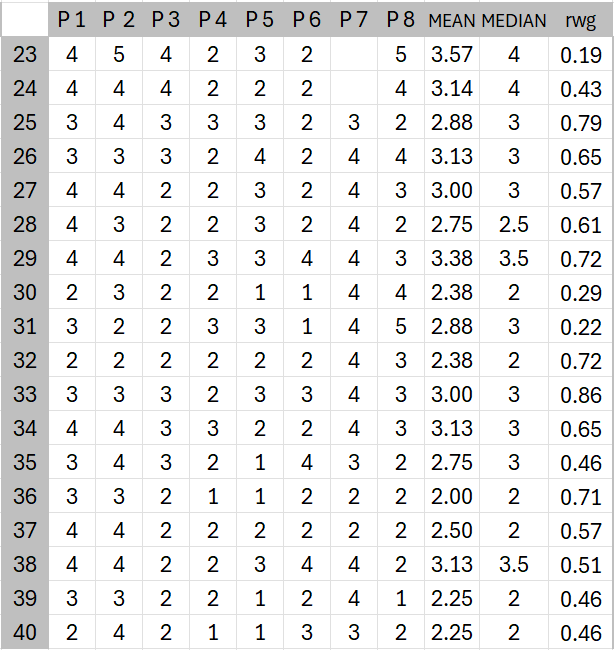 | 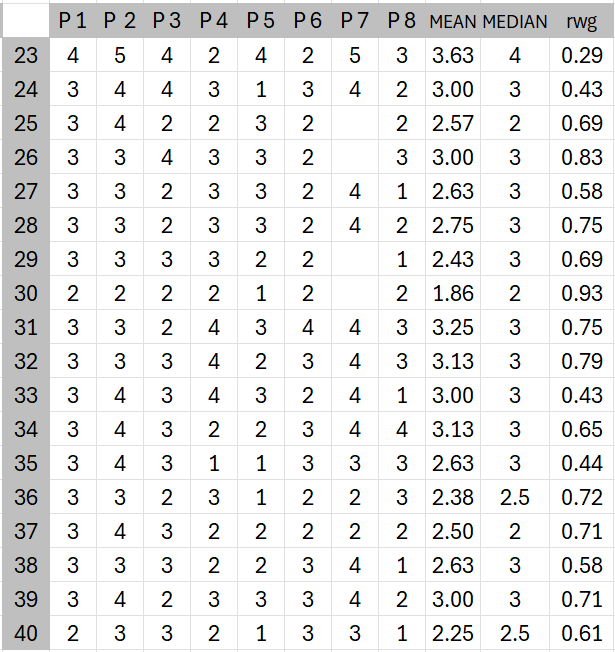 |
|  | Potentially harmful? | | |
|  | DeepSeek-V3 | Llama-3.3-70B-Instruct | GPT-4o |
|  | 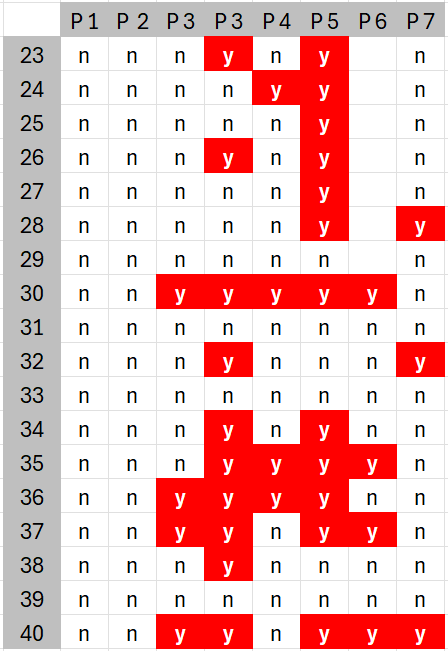 | 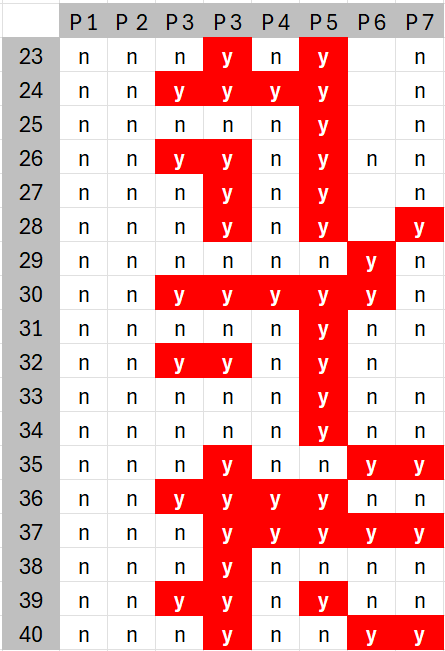 | 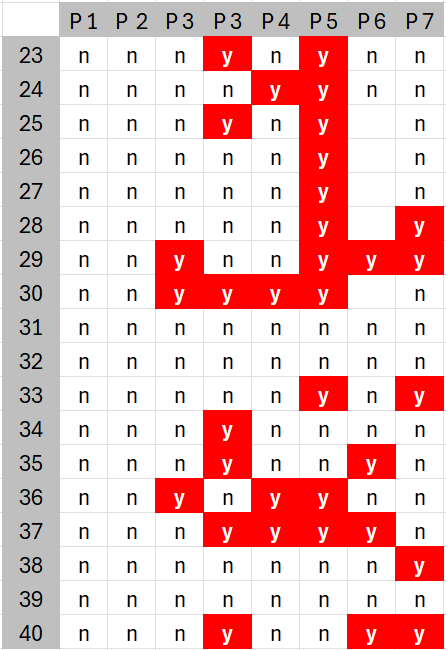 |
